# Baseline host inflammatory and transcriptional profiles associated with structural and functional recovery in drug-resistant tuberculosis

**DOI:** 10.64898/2026.03.07.25342291

**Authors:** Juan M García-Illarramendi, Chiara Sopegno, Kaori L. Fonseca, Lilibeth Arias, Ketevan Barbakadze, Iza Jikia, Malkhaz Tsotskhalashvili, Tamta Korinteli, Zaza Avaliani, Nestani Tukvadze, Sergo Vashakidze, Judith Farrés, Cristina Vilaplana

**Author notes:** Shared last co-authorship (Judith Farrés, Cristina Vilaplana). Corresponding author: Cristina Vilaplana, MD, PhD Head of the Experimental Tuberculosis Unit, Microbiology Dept. Germans Trias i Pujol Research Institute and Hospital (IGTP-HUGTIP) Can Ruti Campus Crtra. de Can Ruti, Camí de les Escoles, s/n 08916, Badalona (Barcelona) Catalonia. (Spain) Telf.

## Abstract

**Background:** In extensively drug-resistant and pre–extensively drug-resistant TB, bacteriology-based monitoring often fails to capture structural lung recovery and patient-reported functional health. We aimed to characterize multidomain treatment response and examine host inflammatory and transcriptional features associated with incomplete recovery.

**Methods:** We conducted an ancillary analysis of a prospective, open-label, pilot study evaluating adjunctive ibuprofen in XDR-TB (NCT02781909). Participants were assessed at baseline and during treatment using TBS, chest radiography, sputum culture, SGRQ, blood cell indices, plasma cytokines, and whole-blood transcriptomic profiling. Clinical and laboratory measures were compared across outcome groups, and blood transcriptional profiles were analyzed in relation to treatment outcomes.

**Results:** Here we show that microbiological and symptomatic improvement occurred earlier than radiological and functional recovery. Higher baseline systemic inflammation, including elevated NLR, SII, and IL-6, as well as increased expression of interferon-related genes such as *CD274* and *GBP5*, were associated with poorer radiological and SGRQ outcomes at 6 months. In contrast, transient elevations of IL-8 and IL-4 were associated with early bacteriological clearance. IL-8 was the only plasma biomarker consistently correlated with symptom severity, radiological findings, and functional health.

**Conclusions:** Treatment response in drug-resistant TB is asynchronous across biological domains. Integrated host profiling identifies inflammatory and transcriptional features associated with incomplete structural and functional recovery, supporting the use of multidimensional endpoints to better capture long-term outcomes and inform individualized patient management.

**Plain Language Summary:** People with highly drug-resistant tuberculosis can clear the infection but still experience lung damage and reduced quality of life after treatment. In this study, we examined recovery using several measures, including symptoms, chest X-rays, blood markers of inflammation, and gene activity, in addition to tests for tuberculosis bacteria. We analyzed data and stored samples from a small clinical trial to see how these measures changed over time. We found that lung structure and quality of life improved more slowly than bacterial clearance. People with higher levels of inflammation before treatment were more likely to have ongoing lung changes and poorer quality of life later. These results suggest that tuberculosis care should look beyond bacterial clearance and include monitoring inflammation to better support long-term recovery.

## Introduction

Tuberculosis (TB) remains one of the leading causes of morbidity and mortality worldwide. Extensively drug-resistant (XDR) TB accounted for over 4% of all TB cases diagnosed in 2022^1^. Despite a decline in the proportion of XDR-TB cases, and the appearance of newer and shorter regimens^2^, the treatment success rate for these patients remains far below the World Health Organization’s (WHO) target of 75%^3,4^.

Effective assessment of TB treatment outcomes is crucial for optimizing patient management. Monitoring treatment to identify those at risk of poor outcomes has the potential to improve overall success by individualizing treatment duration without increasing the risk of relapse.

According to the 2023 WHO Target Product Profiles for treatment monitoring, a ’good outcome’—or sustained treatment success—is defined as bacteriological and/or clinical improvement at the end of treatment without evidence of relapse within 6 months. Conversely, a ’poor outcome’ encompasses a lack of improvement, early relapse, the necessity to switch or terminate treatment prematurely, or TB-related death ^5^. This framework acknowledges that traditional programmatic definitions of ’cure,’ which rely heavily on sputum culture conversion, are limited in their ability to capture the complexities of tissue healing and full patient recovery. Positron Emission Tomography–Computed Tomography studies in drug-sensitive TB have shown new or expanding lung lesions at four weeks of treatment despite eventual culture conversion, illustrating this disconnect^6^. During TB treatment, sputum smear microscopy and mycobacterial culture show limited sensitivity and only moderate specificity in identifying patients at risk of failure or relapse^7^.

The use of host blood mRNA transcripts holds strong potential to enhance TB prognosis and treatment monitoring^8^. However, the clinical impact of treatment strategies guided by mRNA signatures still requires evaluation in prospective trials^9^. In addition, plasma cytokines and hematologic indices can provide complementary information on inflammatory activity during treatment^10^.

In resource-limited settings, symptom resolution often serves as a pragmatic surrogate when laboratory assays are inaccessible. Changes in cough frequency and intensity, febrile episodes, weight gain, and overall functional capacity deliver invaluable real-time insights into disease trajectory and therapeutic efficacy^11^. However, in XDR-TB, radiographic abnormalities (such as cavitation, fibrosis, and consolidation) may persist long after sputum cultures convert to negative, underscoring the need to track tissue-level healing, not just culture status, to guide extended or adjunctive interventions. TB treatment also significantly impacts MDR/XDR-TB survivors, who frequently report long-term physical health challenges and worse health-related quality of life (HQoL)^12^, highlighting the need for early intervention and holistic care to mitigate lasting impacts.

Systemic inflammation plays a dual role in TB treatment. While an effective immune response is necessary for bacterial clearance, excessive or prolonged inflammation can contribute to lung tissue damage and poor treatment outcomes. Elevated inflammatory markers have been associated with delayed bacterial clearance and an increased risk of post-TB lung disease (PTLD)^13^. Several studies have explored the host immune response to TB infection as a correlate of disease severity or a measure of treatment response, often through monitoring cytokine levels or transcriptomic profiling^8,14–18^ However, these immune signatures have largely been defined according to microbiological criteria. Transcriptomic profiling of TB granulomas has identified discrete immune-inflammatory and tissue-repair modules whose expression levels correlate with disease severity and treatment response in both drug-sensitive and MDR/XDR-TB^19^.

Additionally, modulation of the host immune response using existing drugs as adjuvants to standard of care (SoC) has been proposed to improve treatment response and reduce long-term effects without promoting drug resistance^20^. Within host-directed therapies (HDT), nonsteroidal anti-inflammatory drugs (NSAIDs) have been selected for their anti-inflammatory and immunomodulatory properties, relatively safe profile, and availability. Preclinical models have demonstrated clear benefits, although clinical evidence remains limited^21,22^. A recent pilot clinical study (NCT02781909) on the use of ibuprofen as adjunct to SoC in XDR-TB found the treatment to be safe and associated with a reduction in inflammatory markers. However, it did not show a significant advantage in the study’s primary outcomes^23^.

Quantifying clinical improvement is inherently complex: depending on whether one emphasizes physiological measures, functional assessments, patient-reported quality of life, or biomarker trajectories, markedly different conclusions may emerge. Microbiological monitoring remains the cornerstone of TB treatment evaluation, yet the disease’s heterogeneity and risk of treatment failure mandate a multiparametric approach. Integrating microbiological, radiological, clinical, and immunological data provides a more comprehensive framework for assessing treatment response, particularly in relation to long-term outcomes and PTLD, and it has been published that TB clinical trials should incorporate holistic endpoints, including lung function and quality-of-life measures^24^.

This study aimed to comprehensively characterize treatment response in individuals with pre-XDR and XDR-TB -independent of treatment received- by integrating clinical severity, radiological imaging, microbiological status, HQoL, plasma inflammatory biomarkers, and blood transcriptional profiles. The objective was to explore how baseline and longitudinal host characteristics relate to early and late treatment outcomes across domains, with relevance for understanding the heterogeneity of recovery and potential long-term sequelae.

## Materials & Methods

### Study design and participants

For this ancillary study we used the data and biobanked samples from individuals included in a phase-II prospective, interventional, open-label pilot trial to study the efficacy and safety of the use of adjunctive ibuprofen on pre-XDR and XDR TB patients (NSAIDS-XDRTB trial, NCT02781909). The methodology and main results of the pilot study have been reported previously^23^. This ancillary study builds upon the NSAIDS-XDR-TB clinical trial, expanding its scope to investigate host-derived determinants of treatment response.

### Data

We used the NSAIDS-XDR-TB trial database, previously extracted from the electronic case report form (CRF). Data collected included demographic information, medical history, and concomitant medications. Radiological efficacy was assessed using a scoring system that aggregates all X-ray findings (X-ray score) reported in the CRF. Parenchymal abnormalities, both primary and secondary, along with nodular and pleural abnormalities were considered^23^. TB disease severity (TBS) was evaluated at baseline using a score adapted from the Bandim TB score^25^ based on the data available^23^. The Saint George Respiratory Questionnaire (SGRQ) was used for measuring patients’ HQoL. The SGRQ measures overall health and well-being with an established reference value for healthy individuals^26^.

### Samples

We used biobanked frozen samples of the NSAIDS-XDRTB trial. Peripheral whole blood samples had been collected at baseline, months 2 and 6 of treatment. Plasma was isolated from whole blood collected in sodium citrate CPT tubes (BD Vacutainer® CPT^TM^). Whole blood collected in PAXgene® tubes (PreAnalytiX GmbH, Hombrechtikon, Switzerland) were used for the RNA-sequencing analysis. Samples were stored at -80⁰ until analysis.

### Inflammatory biomarkers

Composite general inflammatory indices were calculated using the available data. These indices integrate independent white blood cell subsets, including the neutrophil-to-lymphocyte ratio (NLR), the monocyte-to-lymphocyte ratio (MLR), the erythrocyte sedimentation rate (ESR) and the systemic immune-inflammation index (SII), which is computed as NLR multiplied by the platelet count. These indices were selected because they are derived from routinely collected full blood counts within the program setting and have been previously associated with TB severity and treatment response. CRP was not protocol-specified and was not routinely available at the study site during the study period.

Levels of circulating pro- and anti-inflammatory cytokines in plasma were assessed by immunoassay. Briefly, plasma samples isolated from whole blood using CPT tubes were processed within 2 hours and recovered by centrifugation. Levels of tumor necrosis factor (TNF), Interferon gamma (IFNG) and interleukins (IL) IL-1B, IL-2, IL-4, IL-6, IL-8, IL-10, IL-12, and IL-17A were measured using the Millipore’s MILLIPLEX MAP human high sensitivity T cell magnetic Bead panel (cat#HSTMAG-28SK, EMD Millipore corporation). The inflammatory markers were evaluated in a total of 74 plasma samples from the 28 enrolled participants at baseline, months 2 and 6. Assays were performed according to the manufacturer’s instructions and plates read using a Luminex® 200 System.

### RNA extraction and sequencing

Total RNA was extracted from 74 whole blood samples using the PAXgene Blood miRNA Kit (PreAnalitiX, Hombrechtikon, Switzerland) procedure, according to the manufacturer’s protocol. Total RNA quality was quantified using the Quant-iT Broad-Range dsDNA Assay Kit (Thermo Fisher Scientific), and RNA integrity was assessed using the Fragment Analyzer system HS RNA Kit (15NT) (Agilent). RNA-Seq libraries were prepared with the Illumina Stranded Total RNA Prep with Ribo-Zero Plus (Illumina), following the manufacturer’s recommendations, starting with 0·5 μg of total RNA as the input material. The final library was validated on an Agilent 2100 Bioanalyzer using the DNA 7500 assay. The libraries were sequenced on the NovaSeq 6000 (Illumina) with a read length of 2×101 bp following the manufacturer’s protocol for dual indexing. Image analysis, base calling, and quality scoring of the run were performed using the manufacturer’s software Real Time Analysis (RTA v3.4.4), followed by the generation of FASTQ sequence files. Expression of genes as raw counts was used in downstream differential expression analyses.

### Transcriptomics analysis

Differentially expressed genes (DEG) between different TB patient groups were identified with *DESeq2* package^28^ by applying Wald’s test and false discovery rate (FDR) of 0.05.

Overrepresentation analyses of the DEG against REACTOME pathways^29^ was performed on upregulated and downregulate DEGs separately, with the hypergeometric test, an FDR of 0.05 and a gene coverage of at least 10% of the input set.

Seven previously published blood RNA signatures associated with tuberculosis treatment outcomes (bacterial clearance) were extracted from the *TBSignatureProfiler* R package^30^ (Tables Table 1). These signatures were originally developed in independent cohorts; in our study, they were applied descriptively to explore their association with clinical, radiological, and microbiological outcomes. Single-sample gene set enrichment analysis (ssGSEA) scores^31^ were generated for each participant and time point for each of the seven published signatures and for the DEG sets identified.

**Table 1:** List of predictive gene signatures of TB treatment success, selected from *TBSignatureProfiler*.

| Signature name | Efficacy criteria |
| --- | --- |
| **Heycken_FAIL_22<sup>14</sup>** | Negative *M. tuberculosis* culture status at 6 months after treatment initiation without positive cultures thereafter and no disease recurrence during the follow-up period of 1 year after therapy end. |
| **Thompson_FAIL_13<sup>8</sup>** | Proven and maintained sputum culture negativity by month 6. |
| **Long_RES_10<sup>16</sup>** | Response criteria defined in the Thompson TB population |
| **Tabone_RES_25<sup>17</sup>** | Response criteria defined in the Thompson TB population |
| **Tabone_RES_27<sup>17</sup>** | Response criteria defined in the Thompson TB population |
| **Thompson_RES_5<sup>8</sup>** | Proven and maintained sputum culture negativity by month 6 |
| **Zimmer_RES_3<sup>18</sup>** | Negative microbiological test by month 6 or latest month 12 and improved or resolved symptoms |

### TB treatment outcomes

Outcome definitions and thresholds were specified for this ancillary analysis before transcriptomic and cytokine analyses were performed.

#### Microbiological perspective

MDR-TB and XDR-TB cases have been reported to achieve culture conversion after a median of 68.6 days (IQR 61.0–76.1)^27^. In this study, SCC within three months of treatment initiation was used to define fast responders (SCC outcome). The three-month threshold was chosen because it represents an earlier conversion than the cohort’s median time to SCC (four months) while maintaining a balanced distribution of participants between response groups (10 fast vs. 12 slow converters). This cutoff therefore provides sufficient contrast to explore biological differences associated with early versus delayed bacterial clearance.

#### Clinical perspective

Individuals who achieved a ≥50% reduction in the TBS score by month two (TBS outcome M2) or a ≥75% reduction by month six (TBS outcome M6) were considered good responders, as per decided by consensus by the SMA-TB Consortium^32^.

#### Radiological perspective

Individuals who achieved a ≥50% reduction in the X-ray score by month three (X-ray outcome M3) or a ≥75% reduction by month six (X-ray outcome M6) were considered good responders.

#### HQoL perspective

Study participants with SGRQ scores within the normal range (defined as below 7, based on the 95% confidence interval for individuals without a history of respiratory disease^26^) at months two (SGRQ outcome M2) and six (SGRQ outcome M6) after treatment initiation were considered good responders.

### Statistical analysis

Analyses were performed according to the predefined outcome classifications described above, and followed an intention-to-treat approach, including all allocated patients. Continuous data were reported as medians with first and third quartiles. The Mann-Whitney U-test was used to compare two groups for ordinal or continuous variables. The Kruskal-Wallis’s test followed by the Dunn’s post-hoc test was applied for comparisons of continuous variables of three or more groups. Associations between two numerical or ordinal variables were assessed using Spearman’s rank correlation. Categorical data were summarized by study group as counts and percentages. Absolute risk differences were calculated with 95% confidence intervals (CIs) using the Newcombe method for differences in proportions. All statistical tests were two-sided, with a significance level of 5% (α = 0.05). FDR values of the correlation tests were computed to mitigate the type I error rates and correlations with FDR < 0.05 were deemed as significant. Grader agreement between the different outcome classifications was assessed using Cohen’s kappa statistic. Statistical analyses were conducted using RStudio (v4.3.1) and Python (v3.11.6).

## Results

### Divergent response across domains during treatment

We assessed multidimensional TB treatment response across microbiological, clinical, radiological, and HQoL domains. Baseline characteristics of participants who did or did not achieve the outcomes defined as surrogates of good or poor response are summarized in Table 2, with no significant differences in demographics or medical history across comparisons.

**Table 2.**
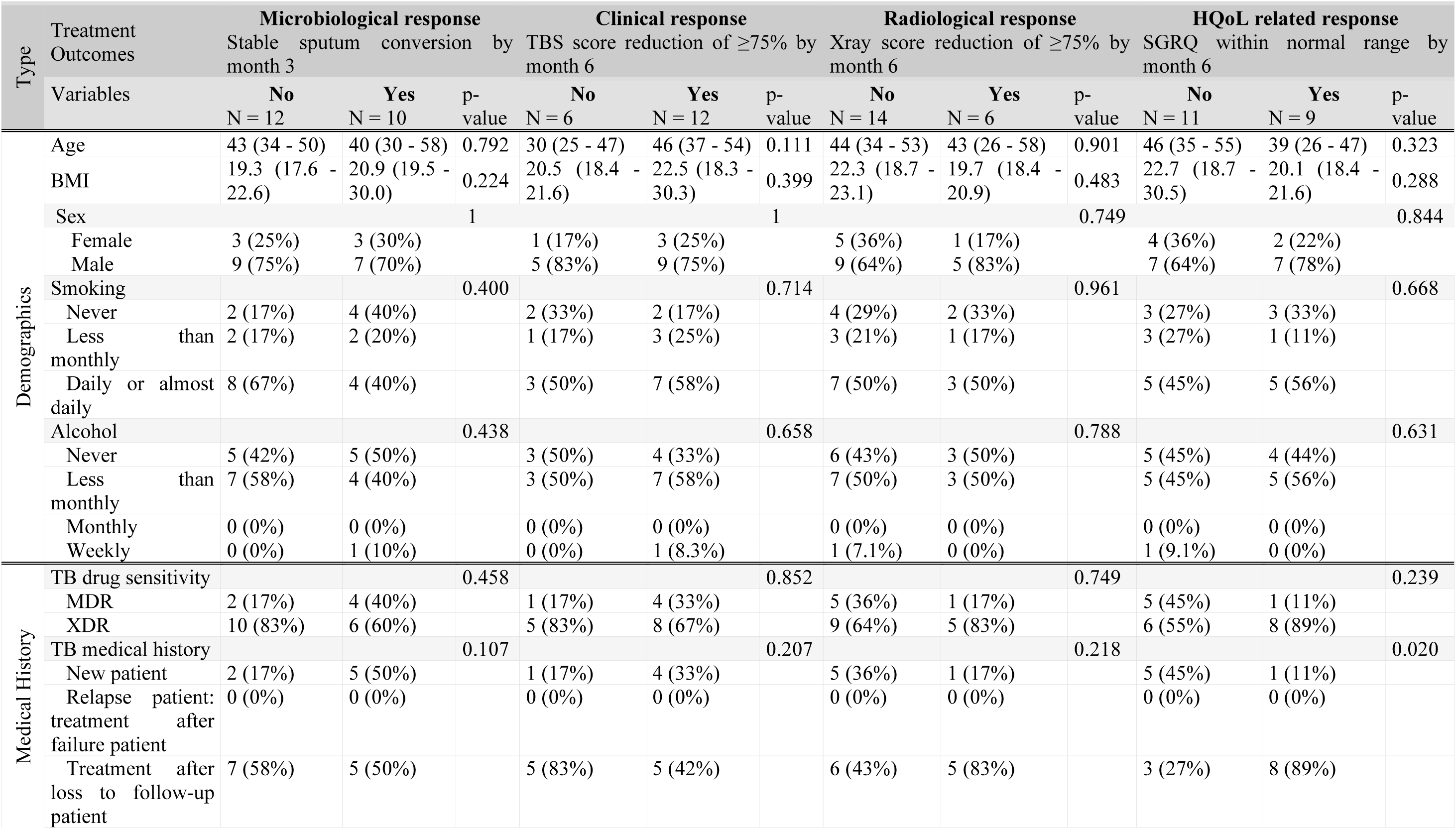
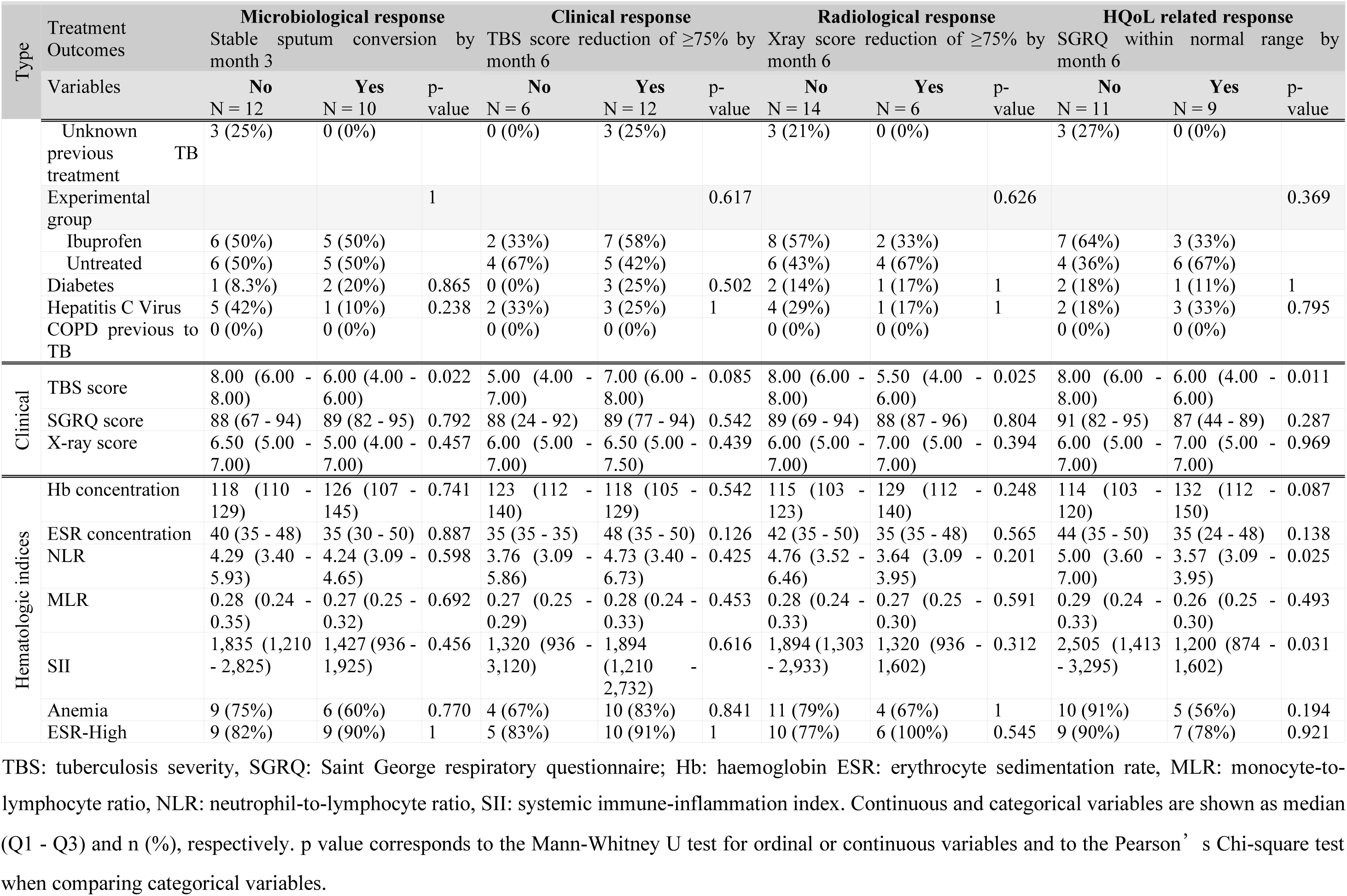
Baseline characteristics per outcome.

Early improvements were largely confined to microbiological and clinical severity scores, whereas radiological and HQoL recovery lagged. By month 2, ∼45 % of the participants achieved stable sputum culture conversion and ≥50 % reduction in TBS score was seen for over 50% of the participants, but less than 10 % of the participants met the thresholds for X-ray or SGRQ outcomes (Fig. 1a, Supplementary Table 1). At month 6, all participants achieved sputum culture conversion, 67 % of the participants achieved ≥75 % reduction in TBS score, 30 % of the participants achieved ≥75 % reduction of the X-ray score, and 45 % of the participants attained normal HQoL scores.

**Figure 1.**
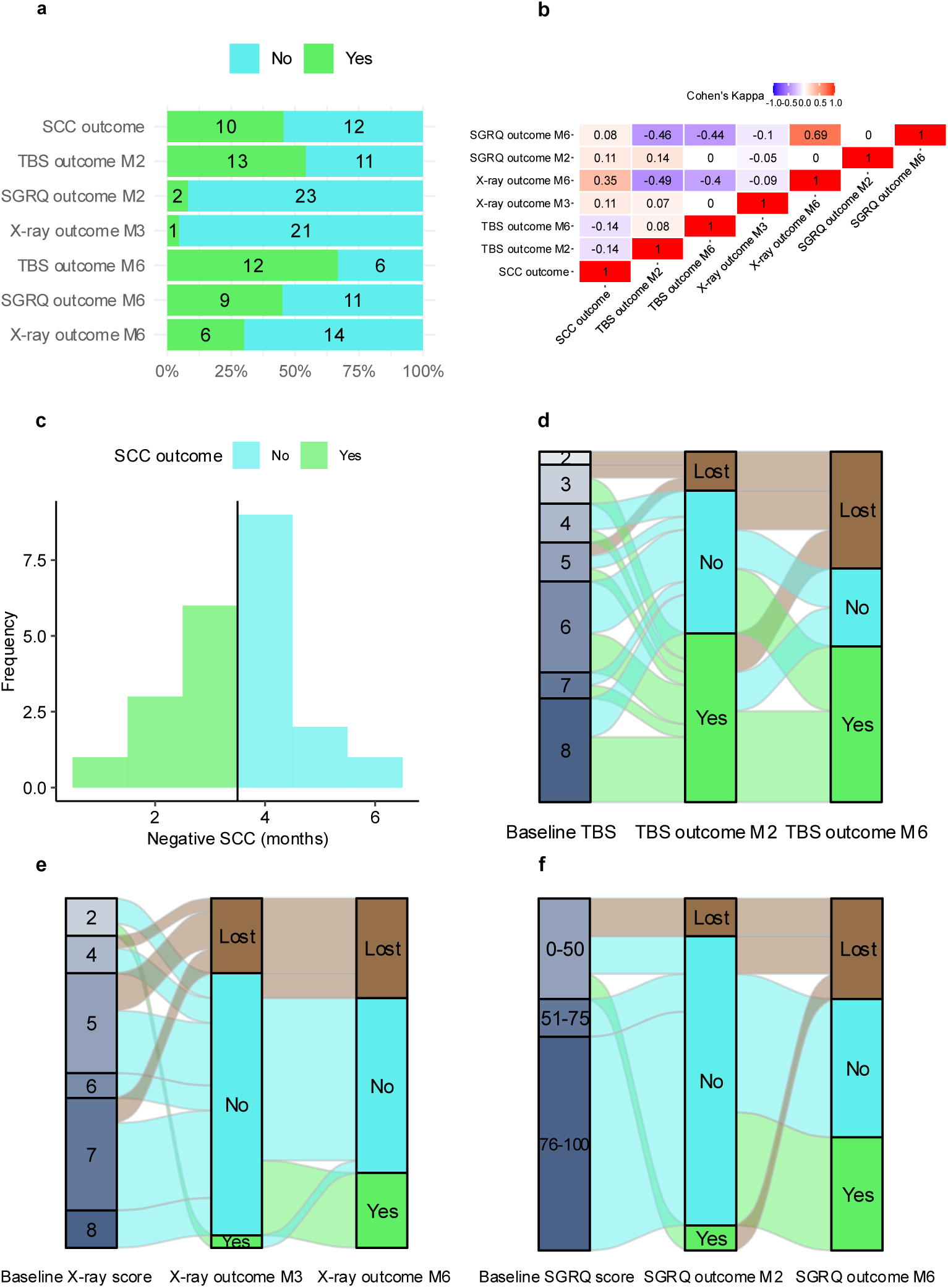
Treatment response across domains. **a)** Proportion of participants achieving (Yes) or not achieving (No) the different outcomes. **b)** Outcome concordance across domains and time, measured by Cohen’s kappa. **c)** Histogram of time to stable sputum culture conversion. Alluvial plots depicting individual patient evolution from baseline score to early (M2-M3) outcomes achievement and from early to late (M6) outcomes achievement; **d)** related to TBS score**, e)** related to X-ray score and **f)** related to SGRQ score.

Agreement among the different outcome domains, assessed using Cohen’s kappa, was generally limited (Fig. 1b). Substantial agreement was observed between the X-ray and SGRQ outcomes M6 (κ = 0.69). A fair level of agreement was found between the SCC outcome and the X-ray outcome M6 (κ = 0.35). In contrast, a moderate disagreement (indicating consistently opposite classifications) was observed between the X-ray and SGRQ outcomes M6 and the TBS outcomes M2 and M6, with κ values ranging from -0.40 to -0.49

Our data also indicate that recovery trajectories vary independently of initial disease severity. As shown in the alluvial plots (Fig. 1d–f), some participants with high baseline scores achieved good outcomes at month2, yet response at month 2 did not consistently translate into outcome achievement at month 6. This variability underscores that outcome achievement is not solely determined by baseline status or early response, but by asynchronous and multidimensional recovery processes.

Taken together, these findings show that bacterial and symptom resolution precede structural and functional recovery and early symptoms resolution does not consistently lead to structural and functional recovery at a later point, emphasizing the need for multidomain monitoring.

### Baseline features associated with later structural and HQoL improvement

We investigated whether baseline clinical and inflammatory characteristics were associated with later treatment outcomes. As shown in Table 2, baseline values of most clinical scores did not differ between participants who achieved or did not achieve their respective outcomes. However, baseline TBS scores were significantly higher in participants who failed to achieve the SCC at month 3 (p-value = 0.022), X-ray (p-value = 0.025), and SGRQ (p-value = 0.011) outcomes at month 6 (Fig. 2, Table 2). These differences were not maintained at later time points. Participants who achieved the TBS outcome M2 had significantly lower baseline SGRQ scores (p-value = 0.037), and this difference persisted throughout follow-up (p-value = 4.53e-3 and p-value = 0.013 at months 2 and 6, respectively; Fig. 2d).

**Figure 2.**
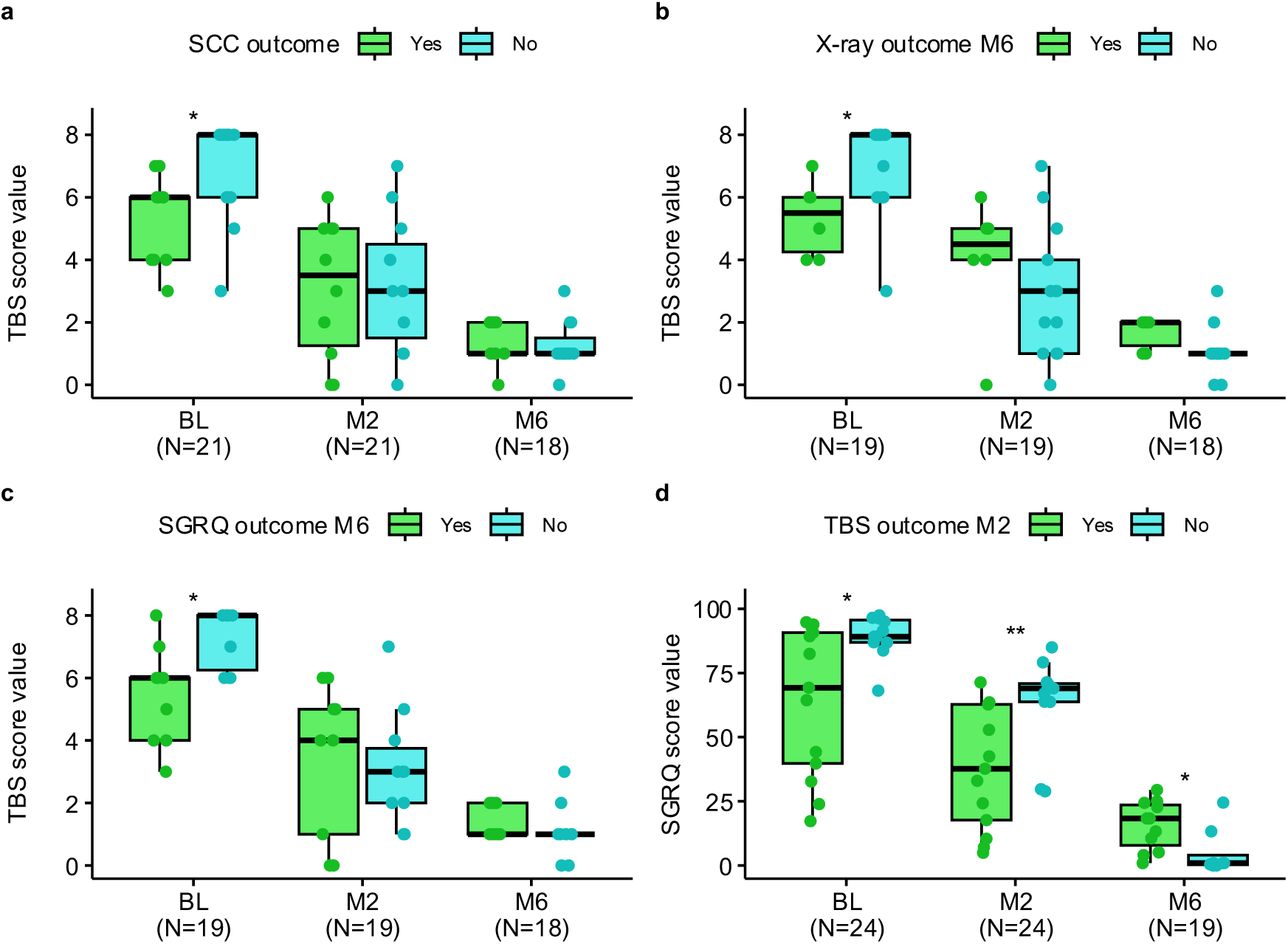
Clinical scores along treatment stratified by outcome achievement. Clinical scores values at baseline (BL), month 2 (M2) and month 6 (M6) for participants who achieved (Yes) or did not achieve (No) the specified outcomes. TBS score stratified by: **a**) SCC outcome, **b**) X-ray outcome M6 and **c**) SGRQ outcome M6. SGRQ score stratified by TBS outcome M2 **d**). Boxplots show the IQR; lines denote medians. The bounds of the box represent the 25th and 75th percentiles; whiskers span 1.5x IQR. Statistical significance was assessed using the Mann–Whitney U test at each time point. Asterisks denote significance levels: **: p-value < 0.01, *: p-value < 0.05

Regarding hematologic indices, good responders for the SGRQ outcome M6 had significantly lower NLR at baseline (p-value = 0.025) and significantly lower SII values at baseline (p-value = 0.031) and month 1 (p-value = 0.04; Fig. 3a–b, Table 2). Although baseline ESR values were not significantly different, they became significantly lower in good responders by month 1 (p-value = 7.02e-4) and remained so during treatment (p-value = 0.039, p-value = 2.79e-3, p-value = 1.29e-4, p-value = 0.040 and p-value = 0.032 for month 2 to 6 comparisons, respectively; Fig. 3c). Analyses based on ESR categories (normal vs high) also showed significant differences at months 1 and 6 (Supplementary Fig. 1a). Participants who achieved the SGRQ outcome M6 had higher hemoglobin levels at month 1 (p-value = 0.033), corresponding to a lower prevalence of anemia (Fig. 3d, Supplementary Fig. 1b), though this was not sustained later. For the X-ray outcome M6, the only blood parameter showing a significant difference was ESR at month 1(p-value = 0.038; Fig. 3e).

**Figure 3:**
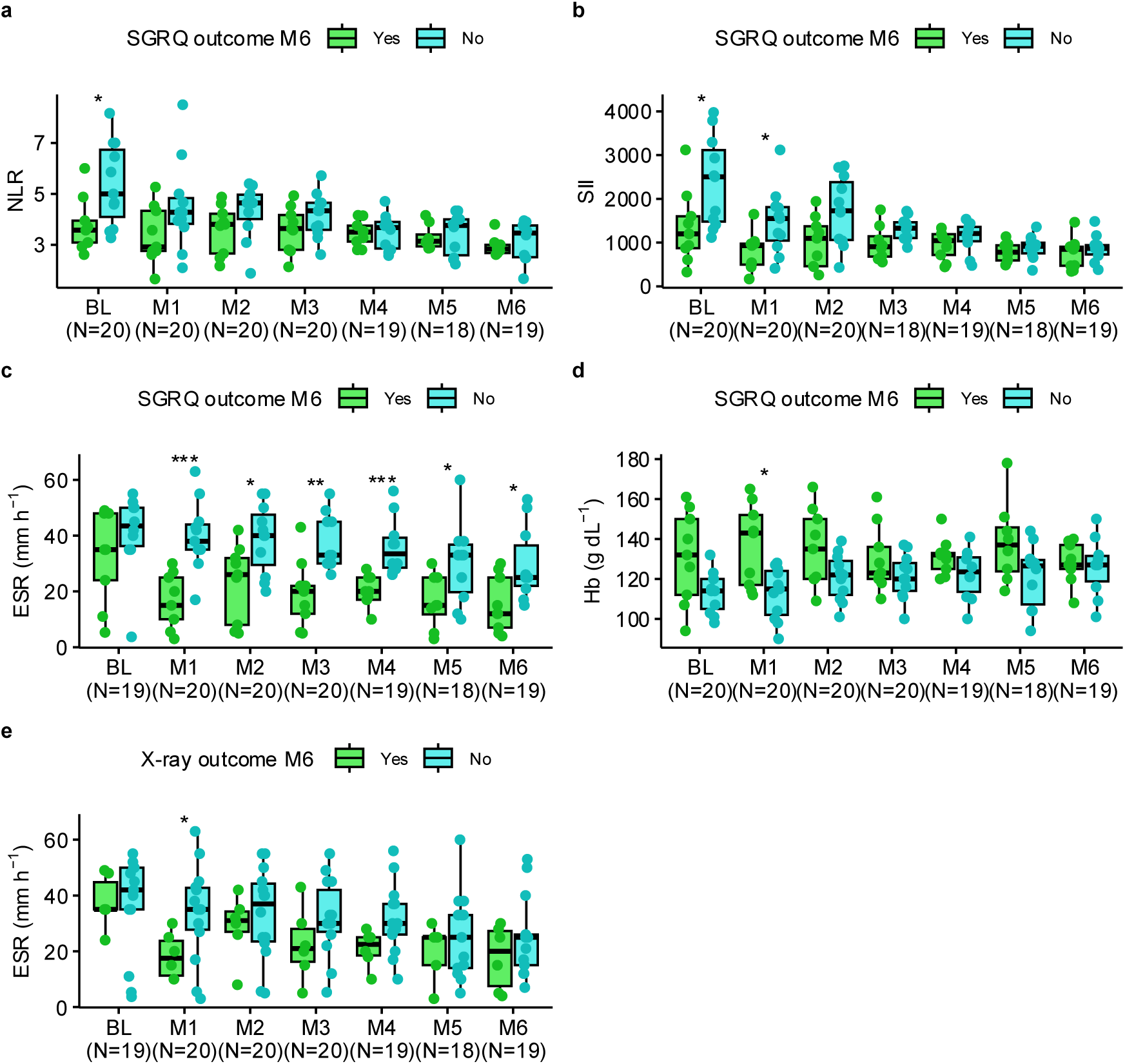
Hematological indices along treatment stratified by outcome achievement. Hematological indices at baseline (BL) and all follow-up months (M) for participants who achieved (Yes) or did not achieve (No) the specified outcomes. **a)** NLR (neutrophil-to-lymphocyte ratio), **b)** SII (systemic inflammatory index), **c)** ESR (erythrocyte sedimentation rate) and **d)** Hb (hemoglobin) values stratified by the SGRQ outcome M6. **e)** ESR values stratified by the X-ray Outcome M6. Boxplots show the IQR; lines denote medians. The bounds of the box represent the 25th and 75th percentiles; whiskers span 1.5x IQR. Statistical significance was assessed using the Mann–Whitney U test at each time point. Asterisks denote significance levels: ***: p-value < 0.001, **: p-value < 0.01, *: p-value < 0.05

At baseline, among the ten plasma biomarkers measured, IL-6 levels were significantly lower (p-value = 6.23e-3) in participants who later achieved the SGRQ outcome at month 6, but this difference was no longer evident at months 2 or 6 (Fig. 4a). By month 2, additional cytokine differences emerged, revealing two distinct inflammatory trajectories. Late outcomes showed a similar pattern to IL-6, with lower levels of IL-8 (p-value = 0.025) and IL-10 (p-value = 0.035) associated with achieving the SGRQ (Fig. 4b) and TBS (Fig. 4d) outcomes at M6, respectively. In contrast, participants who achieved early outcomes (SCC and the TBS M2) displayed significantly higher cytokine levels at the time of outcome achievement, specifically IL-8 (p-value = 0.031; Fig. 4c) and IL-4 (p-value = 0.028; Fig. 4e), respectively.

**Figure 4:**
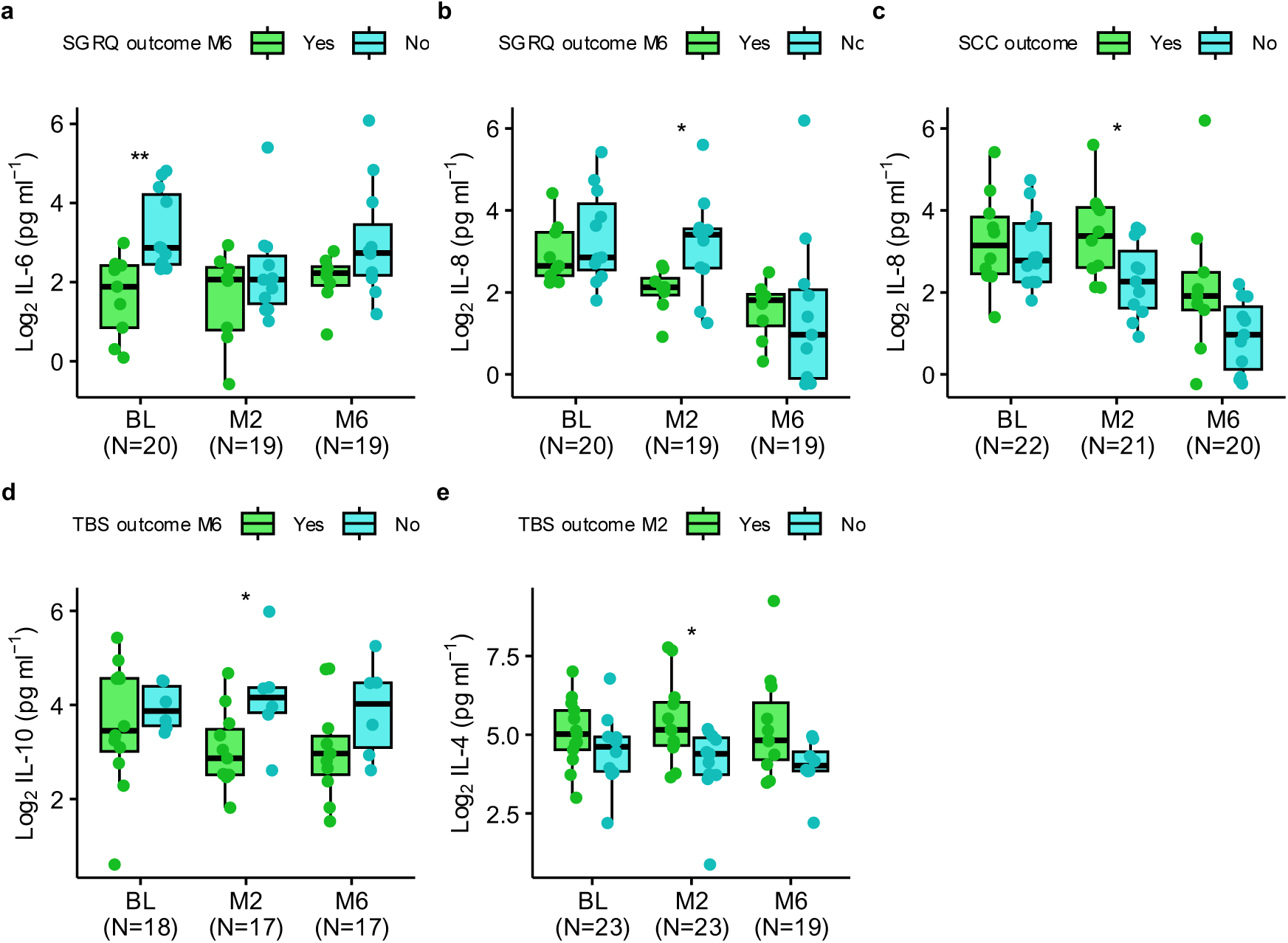
Plasma biomarkers along treatment stratified by outcome achievement. Plasma biomarkers concentration at baseline (BL), month 2 (M2) and month 6 (M6) for participants who achieved (Yes) or did not achieve (No) the specified outcomes. **a)** IL-6 stratified by SGRQ outcome M6. **b)** IL-8 stratified by SGRQ outcome M6 and **c)** SCC outcome. **d)** IL-10 stratified by TBS outcome M6. **e)** IL-4 expression stratified by TBS outcome M2. Boxplots show the IQR; lines denote medians. The bounds of the box represent the 25th and 75th percentiles; whiskers span 1.5x IQR. Statistical significance was assessed using the Mann–Whitney U test at each time point. Asterisks denote significance levels: ***: p-value < 0.001, **: p-value < 0.01, *: p-value < 0.05

### Baseline whole-blood transcriptional profiles associated with treatment outcomes

Differential gene expression at baseline was analyzed in relation to SCC and month 6 clinical outcomes, and the resulting differentially expressed genes (DEGs) were assessed for enrichment in Reactome pathways.

The comparison between fast and slow SCC converters yielded 894 DEGs (SCC DEG set, Supplementary Table 2). Genes upregulated in fast converters were primarily associated with the Immune System (21 % of the DEG set), with most (14 %) related to Innate Immunity, whereas downregulated genes were enriched in Gene expression (Transcription) (22 % of the DEG set) (Supplementary Table 3).

For TBS, X-ray, and SGRQ outcomes, differential expression was analyzed at baseline using the month 6 classifications of good versus poor responders. For the TBS outcome, only one DEG (H2AC19) was identified. For the X-ray M6 outcome, 130 DEGs distinguished good and poor responders (X-ray M6 DEG set, Supplementary Table 4). Downregulated genes in good responders were enriched in the Immune System (23 % of the DEG set) and Interferon Signalling (14 %), whereas upregulated genes showed no significant pathway enrichment (Supplementary Table 3).

For the SGRQ M6 outcome, 180 DEGs were identified (SGRQ M6 DEG set, Supplementary Table 5). Downregulated genes in good responders were enriched in the Immune System (28 % of the DEG set), with 9 % specifically involved in Interferon Signalling, while upregulated genes were enriched in Metabolism of amino acids and derivatives and Translation (10 % of the DEG set) (Supplementary Table 3).

The largest overlap between DEG sets was observed for the X-ray and SGRQ M6 outcomes, with 30 shared genes (Supplementary Fig. 2a, Supplementary Table 6), 11 of which belong to the Immune System and four (*GBP5*, *GBP4*, *TRIM22*, *JAK2*) specifically to Interferon-gamma signalling.

The SCC DEG set, despite being the largest, showed minimal overlap with the other outcome sets—only one gene (*KRT77*) shared with the SGRQ DEG set and seven genes (*AK1*, *MYMS1*, *RGPD2*, *SCAP*, *SCAPER*, *TIA1*, *TTC14*) shared with the X-ray DEG set (Supplementary Fig. 2).

Seven previously published blood RNA signatures predictive of TB treatment success—each originally defined by sputum culture conversion status—were selected for comparison (Table 1). Collectively, these signatures comprise 88 unique genes, with 14 genes shared by at least two signatures (Supplementary Table 7).

Overlap of existing signatures with our outcome-specific DEG sets varied (Supplementary Fig. 2b). Despite being the largest, the SCC DEG set showed the least overlap, sharing only one gene (KIAA0100) with the Thompson_FAIL_13 signature. The SGRQ DEG set overlapped with 22 genes from previously published signatures, while the X-ray DEG set shared three. Two genes, *CD274* (programmed cell death 1 ligand 1, PD-L1) and *GBP5*, showed the highest recurrence across external signatures and were also present in both the SGRQ and X-ray DEG sets. CD274, which modulates T-cell activation thresholds, appeared in four of the seven signatures, and *GBP5*, an interferon-inducible GTPase involved in innate immunity, appeared in three. Both genes were significantly lower at baseline in good responders for the SGRQ and X-ray outcomes at month 6 (Supplementary Fig. 2c-f).

To quantify the relationship between individual participants’ expression patterns and both published and study-derived DEG sets, single-sample gene set enrichment analysis (ssGSEA) scores were calculated. These ssGSEA-derived scores summarize the expression pattern of each signature in each participant. Baseline values of scores based on our outcome-specific DEG sets (SGRQ, SCC, and X-ray) were significantly different between good and poor responders for their corresponding outcomes as expected, with lower scores associated with a good outcome (Table 3, Supplementary Fig. 3).

**Table 3:**
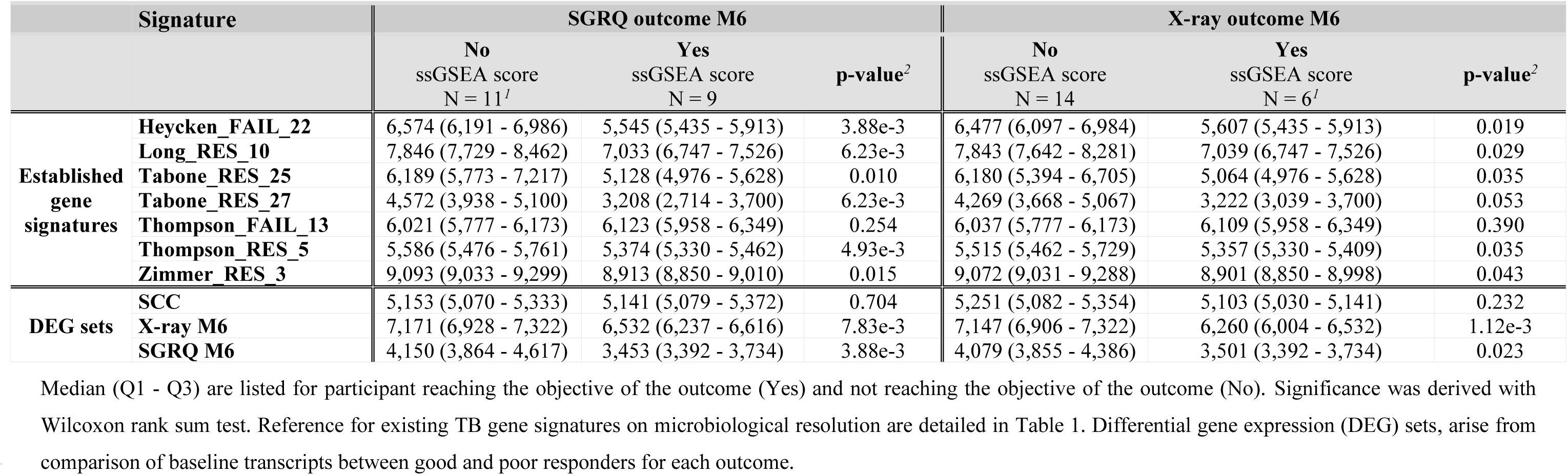
Descriptive statistics of baseline ssGSEA score of established transcriptomic signatures and DEG sets per SGRQ and X-ray outcomes at M6.

|  | Signature | SGRQ outcome M6 |  |  | X-ray outcome M6 |  |  |
| --- | --- | --- | --- | --- | --- | --- | --- |
|  |  | No<br>ssGSEA score<br>N = 11<sup>1</sup> | Yes<br>ssGSEA score<br>N = 9 | p-value<sup>2</sup> | No<br>ssGSEA score<br>N = 14 | Yes<br>ssGSEA score<br>N = 6<sup>1</sup> | p-value<sup>2</sup> |
| Established<br>gene<br>signatures | Heycken_FAIL_22 | 6,574 (6,191 - 6,986) | 5,545 (5,435 - 5,913) | 3.88e-3 | 6,477 (6,097 - 6,984) | 5,607 (5,435 - 5,913) | 0.019 |
|  | Long_RES_10 | 7,846 (7,729 - 8,462) | 7,033 (6,747 - 7,526) | 6.23e-3 | 7,843 (7,642 - 8,281) | 7,039 (6,747 - 7,526) | 0.029 |
|  | Tabone_RES_25 | 6,189 (5,773 - 7,217) | 5,128 (4,976 - 5,628) | 0.010 | 6,180 (5,394 - 6,705) | 5,064 (4,976 - 5,628) | 0.035 |
|  | Tabone_RES_27 | 4,572 (3,938 - 5,100) | 3,208 (2,714 - 3,700) | 6.23e-3 | 4,269 (3,668 - 5,067) | 3,222 (3,039 - 3,700) | 0.053 |
|  | Thompson_FAIL_13 | 6,021 (5,777 - 6,173) | 6,123 (5,958 - 6,349) | 0.254 | 6,037 (5,777 - 6,173) | 6,109 (5,958 - 6,349) | 0.390 |
|  | Thompson_RES_5 | 5,586 (5,476 - 5,761) | 5,374 (5,330 - 5,462) | 4.93e-3 | 5,515 (5,462 - 5,729) | 5,357 (5,330 - 5,409) | 0.035 |
|  | Zimmer_RES_3 | 9,093 (9,033 - 9,299) | 8,913 (8,850 - 9,010) | 0.015 | 9,072 (9,031 - 9,288) | 8,901 (8,850 - 8,998) | 0.043 |
| DEG sets | SCC | 5,153 (5,070 - 5,333) | 5,141 (5,079 - 5,372) | 0.704 | 5,251 (5,082 - 5,354) | 5,103 (5,030 - 5,141) | 0.232 |
|  | X-ray M6 | 7,171 (6,928 - 7,322) | 6,532 (6,237 - 6,616) | 7.83e-3 | 7,147 (6,906 - 7,322) | 6,260 (6,004 - 6,532) | 1.12e-3 |
|  | SGRQ M6 | 4,150 (3,864 - 4,617) | 3,453 (3,392 - 3,734) | 3.88e-3 | 4,079 (3,855 - 4,386) | 3,501 (3,392 - 3,734) | 0.023 |

When baseline ssGSEA scores of the published signatures were compared between good and poor responders for X-ray and SGRQ M6 outcomes, significant differences were observed, whereas no significant differences were found between fast and slow SCC converters (Table 3, Supplementary Fig. 4). For the X-ray outcome, five published signatures showed significant baseline differences, and for the SGRQ M6 outcome, all but one (Thompson_FAIL_13) showed significant differences in ssGSEA scores (Table 3, Supplementary Fig. 4).

Together, these analyses demonstrate that published blood transcriptomic signatures capture biological patterns associated with radiological and HQoL recovery, whereas they do not distinguish early bacteriological responders. The DEG sets follow a trajectory similar to that of the cytokines, revealing two distinct inflammatory patterns: under-expression of immune-related genes is associated with good responders for late structural and HQoL outcomes, whereas overexpression of immune-related genes characterizes participants achieving early SCC.

### On-treatment dynamics of hematologic indices, cytokines, and domain scores

During follow-up, all domain scores declined significantly over the course of treatment (Supplementary Fig. 5). The X-ray score showed a significant reduction from baseline to month 6 and from month 2 to month 6, but not between baseline and month 2, suggesting that most radiological improvement occurred later in treatment. TBS and SGRQ scores showed a significant decline between all timepoints.

Blood parameters also evolved during treatment (Supplementary Fig. 6). ESR and SII decreased significantly by month 1, with SII continuing to decline thereafter. NLR decreased later, reaching significance at month 4 and remaining low at subsequent time points. Hemoglobin concentration and MLR showed no significant longitudinal changes.

Among ten plasma cytokines analyzed, only IL-8 decreased significantly at month 6, relative to both baseline and month 2 (Supplementary Fig. 7).

Longitudinal analysis of whole-blood transcript gene signatures in terms of ssGSEA score revealed dynamic changes over treatment (Supplementary Fig. 8). Enrichment scores for previously established treatment-success signatures declined significantly from baseline to month 6, with the exception of Thompson_FAIL_13, a failure-associated signature that increased significantly at month 6. Similarly, enrichment scores for the SGRQ and X-ray DEG sets decreased over time, whereas SCC-related scores showed no significant longitudinal variation.

In summary, inflammatory indices (ESR, SII) improved rapidly within the first 1–2 months of treatment, whereas NLR and radiological (X-ray score) parameters improved later, reflecting asynchronous recovery kinetics across biological domains. Together, these findings indicate that immune declines during treatment, consistent with resolution of inflammation and tissue repair.

### Multidomain correlations

To gain an integrated view of treatment response, correlation analyses were performed across clinical, hematologic, cytokine, and transcriptomic parameters, pooling all available time points to identify consistent associations (Figure 5; Supplementary Table 8). As expected, strong within-domain correlations were observed, particularly among clinical scores (TBS, X-ray, and SGRQ). Although the domain scores exhibited different rates of recovery or normalization, they still showed significant correlations across timepoints.

**Figure 5.**
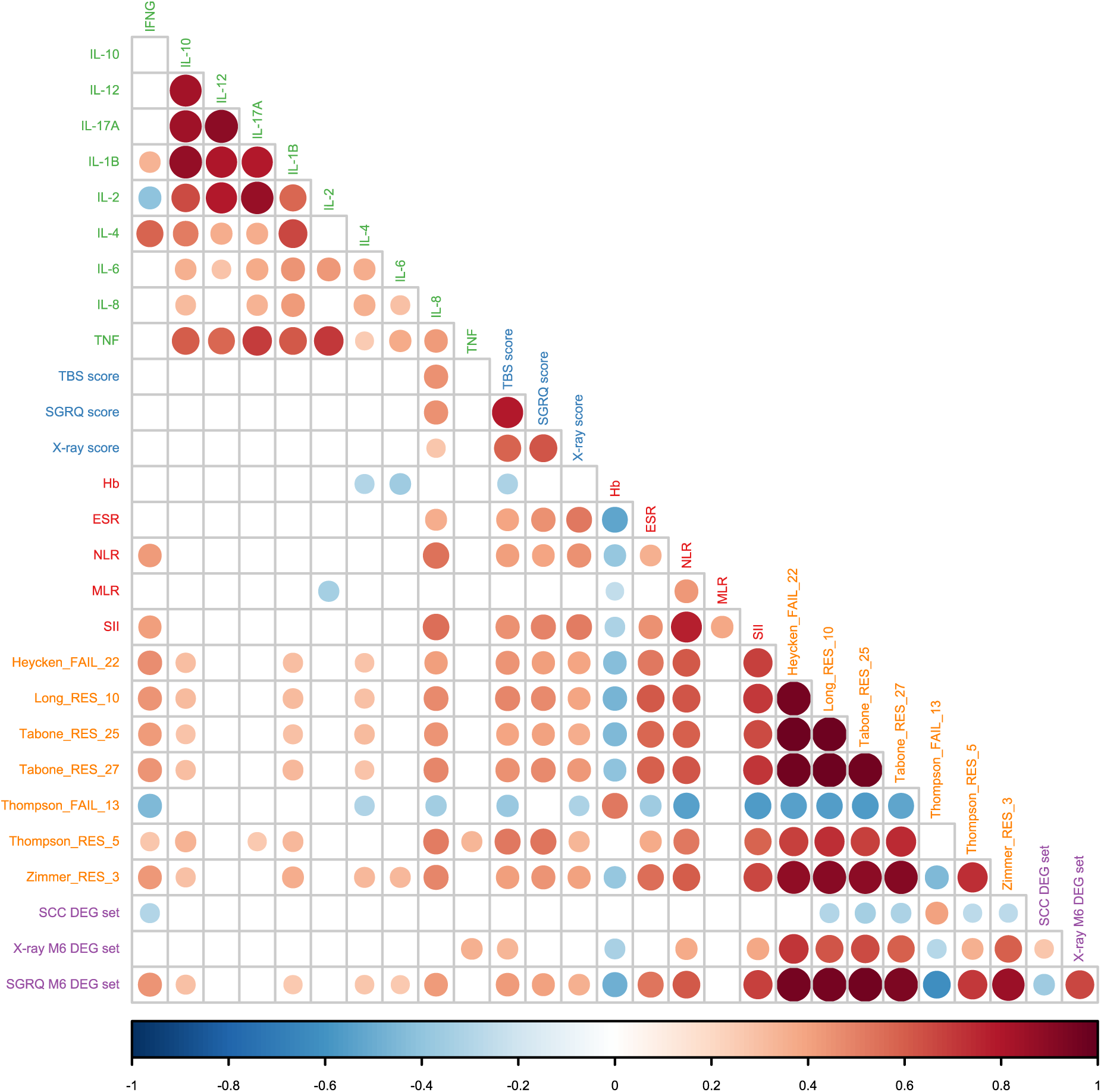
Correlation between the different follow-up parameters. Correlation between plasma biomarkers (green), domain scores (blue), hematological indices (red), ssGSEA scores of established gene signatures (orange) and ssGSEA scores of DEG sets (purple) was assessed using Spearman’s rank correlation. Only the significant correlations (adjusted p-value < 0.05) are shown in the heatmap. Dot size reflects the magnitude of the p-value, and dot color represents the Spearman’s rho coefficient.

Among plasma biomarkers, IL-8 was the only cytokine showing significant positive correlations with all three clinical scores (TBS, X-ray, and SGRQ), in line with being the only cytokine with significant decrease along treatment. Several other cytokines (IFNG, IL-2, IL-4, IL-6, and IL-8) correlated with some hematologic indices, and additional cytokines showed significant associations with gene-signature enrichment scores.

Established gene signatures exhibited highly correlated ssGSEA scores, which aligns with the comparable behavior we observed in prior analyses. A strong correlation was also observed between ssGSEA scores derived from our DEG sets and those from established gene signatures, with the SGRQ DEG set showing the highest overall correlation coefficients. This SGRQ DEG set also displayed the strongest cross-domain associations, correlating with plasma biomarkers, clinical scores, and hematologic parameters. Notably, SCC DEG set ssGSEA scores correlated inversely with several established treatment-success signatures.

Taken together, these integrative analyses highlight robust within-domain coherence (especially among clinical metrics) and reveal distinct cross-domain relationships, with IL-8 emerging as the only plasma biomarker consistently linked to clinical outcomes, and the SGRQ DEG set demonstrating the strongest integrative associations across molecular and clinical layers.

## Discussion

In this study, we leveraged data and samples from a pilot clinical trial in individuals with pre-XDR and XDR-TB to deliver a comprehensive evaluation of treatment response up to month 6 after starting treatment. Beyond conventional bacteriological clearance metrics, we incorporated complementary outcomes (symptom severity, radiological pathology, and HQoL) in line with recent recommendations on the use of more holistic outcome measures^24^. Clinical assessments, radiological findings, plasma biomarkers, and blood transcriptional profiles were analyzed correlating with these outcomes to provide a holistic view of response trajectory. Treatment-arm effects were not considered here, as the trial primary outcomes have been reported elsewhere and showed no statistically significant differences^23^. Longitudinal studies linking host cytokine or transcriptomic profiles to treatment outcomes in XDR-TB patients are scarce. Most existing research has focused on drug-susceptible TB or on one or few specific domains of responses^33,34^, leaving a major gap in understanding host dynamics during treatment for XDR-TB. Our study addresses this gap by integrating serial cytokine and blood transcriptional measurements with diverse clinical outcomes, including symptom severity, radiological findings, and quality of life. This comprehensive approach offers a novel framework for assessing treatment response in XDR-TB. This multidomain framework revealed distinct, asynchronous recovery trajectories and identified host inflammatory and transcriptional features at baseline associated with incomplete structural and functional recovery, providing insights highly relevant to post-TB sequelae.

Treatment was successful in 71 % of participants, according to WHO definitions. While symptom and bacterial clearance occurred early, radiological and HQoL recovery lagged, highlighting a disconnect between microbiological cure and full clinical recovery. This is consistent with prior studies showing that initial rapid symptom improvement is primarily driven by the early bactericidal activity of anti-TB therapy^10,35^, whereas structural and functional recovery is delayed due to residual inflammation or tissue damage. Among participants who completed treatment, all were bacteriologically cured by month 6; however, only 45% achieved a normal-range SGRQ and 30% attained a ≥75% reduction in X-ray score, highlighting the disconnect between microbial cure and full clinical recovery. These findings underscore the limitations of bacteriological outcomes alone for defining treatment success in advanced disease. Clinical, radiological, and functional responses showed limited concordance, indicating asynchronous recovery trajectories. This asynchrony is unlikely to reflect inconsistencies in scoring, as outcome measures were correlated over follow-up, and instead suggests differences in the biological processes underlying bacterial clearance and tissue repair. Baseline symptom severity captured aspects of disease associated with later structural and functional outcomes, whereas baseline radiological and quality-of-life measures alone were not consistently informative.

There is growing evidence that host factors driving lung injury in TB likely contribute to variable patterns of pulmonary impairment after TB^36^ and that systemic immunity in whole blood reflects local processes such as granuloma composition^37^. Accordingly, minimally invasive blood measures may have prognostic value. In our study, markers of systemic inflammation and transcriptional immune activation at baseline were strongly associated with distinct response trajectories.

Across multiple parameters, higher immune activation at baseline is associated with more rapid early bacterial (SCC within 3 months) and symptom resolution (50% TBS reduction at month 2) but poorer late outcomes (month 6). Poor SGRQ responders at month 6 had elevated NLR, SII, and IL-6 at baseline. Transcriptomics likewise showed that baseline immune overexpression correlated with faster sputum conversion but worse HQoL and radiological outcomes at month 6. The disconnect between culture conversion and radiological or HQoL recovery has been noted previously; for example, Rambaran et al. reported that elevated baseline inflammatory markers during active TB were linked both to faster bacterial clearance and to greater disease severity and tissue damage^38^. The correlation between HQoL measures such as SGRQ and lung function in TB has also been described^39^.

NLR, SII, and ESR declined over treatment and were positively correlated with TBS, X-ray, and SGRQ scores. Higher baseline NLR and SII were associated with poorer SGRQ outcome at month 6. Neither NLR nor SII distinguished fast versus slow SCC, consistent with studies suggesting MLR may predict delayed microbiological response^40^; in our cohort, even MLR showed no association. This suggests NLR/SII reflect broader systemic inflammation linked to tissue injury and functional recovery rather than bacteriological response per se. Neutrophils contribute to TB immunopathology, promoting tissue damage and dysregulated immunity^41^; platelets also drive pro-inflammatory phenotypes associated with tissue degradation^42^. Although ESR was not different at baseline, by month 1 future good X-ray and SGRQ responders already showed significantly lower ESR, and this persisted; while nonspecific, ESR appears useful for monitoring^43^. IL-6, a key proinflammatory cytokine, has been linked to TB severity^44^ and poor microbiologic outcomes^45^. In our study, baseline IL-6 did not differ between fast and slow SCC but was associated with poor SGRQ at month 6, consistent with a hyperinflammatory milieu favoring early bacterial killing yet predisposing to chronic symptoms and fibrotic remodeling. At month 2, good early responders (SCC and TBS outcomes) showed higher IL-8 and IL-4, whereas good late responders (SGRQ and TBS outcomes M6) had lower IL-8 and IL-10. IL-8 declined significantly by month 6; while it may aid pathogen clearance^46^, sustained elevations are linked to persistent symptoms and tissue injury.

Blood transcriptomic differences before treatment that predict bacterial-clearance success have been reported^47^. In our comparison of seven published sputum-based gene signatures (88 unique biomarkers; 14 shared across signatures), DEG sets tied to radiological (X-ray) and HQoL (SGRQ) outcomes overlapped more with these signatures than did the large SCC DEG set. Two IFNG-inducible genes — *CD274* and *GBP5*— were present in both our SGRQ and X-ray DEG lists and are among the most recurrent markers across published signatures; both have been linked to TB progression^37,48^. Baseline ssGSEA scores for these published signatures differed between good and poor responders for X-ray and SGRQ month 6 outcomes, but not between fast and slow SCC converters. The discrepancy between our SCC DEG set and the established gene signatures (despite both being defined by bacterial clearance) may reflect differences in the timing of that clearance. Our SCC outcome captures early converters, whereas the published signatures were developed to predict end-of-treatment culture conversion. Altogether these results highlight distinct biology underlying early bacteriological clearance versus later structural/functional recovery.

Together, these findings have two major implications. First, they support multiparametric treatment monitoring beyond bacteriological conversion to include clinical, radiological, and immunological domains, particularly relevant in pre-XDR/XDR-TB where structural disease and inflammatory sequelae are more likely to persist after microbiological cure. Second, they identify baseline inflammatory and transcriptional markers (e.g., NLR, SII, ESR, IL-6, *CD274*, *GBP5*) as candidates for exploratory risk stratification of incomplete recovery and to inform host-directed interventions targeting persistent inflammation, a concept supported by the HDT literature^22,49^.

The main limitations of this study are the small sample size, treatment heterogeneity inherent to an ancillary analysis of a pilot clinical trial, and follow-up limited to six months after treatment initiation. Potential sources of variability include the influence of HDT, differences in resistance severity, and variation in standard-of-care regimens. In addition, outcome thresholds were defined pragmatically to capture clinically meaningful change across domains and should be interpreted as exploratory rather than externally validated endpoints. Despite this heterogeneity, the consistency of associations observed across multiple clinical, inflammatory, and transcriptional domains suggests that our findings capture relevant aspects of TB pathophysiology and treatment response.

Importantly, our findings suggest potential avenues for more individualized treatment strategies in XDR-TB that warrant further validation. Elevated baseline levels of systemic inflammatory markers, including ESR, NLR, and IL-6, as well as immune-related genes such as *CD274* and *GBP5*, may represent candidate markers associated with incomplete recovery in pre-XDR/XDR-TB. Identifying patients at risk of poor structural or functional recovery despite bacteriological cure highlights an opportunity for further investigation of tailored follow-up and monitoring strategies. This perspective aligns with emerging themes in TB research, including the move toward precision medicine in drug-resistant TB. Future studies should aim to validate these findings in larger, prospectively designed cohorts, assess their relevance in patients receiving standardized regimens such as BPaL or BPaLM, and determine whether targeted modulation of inflammatory pathways can improve long-term structural and functional outcomes.

## Data availability

All data supporting the findings of this study are available within the paper and its supplementary Information. RNAseq data is deposited in GEO database under accession number GSE298762.

## Acknowledgments

We would like sincerely to thank the study participants who agreed to be part of the NSAIDS-XDR-TB study and the staff from the NCTLD who conducted the pilot clinical trial. Funding for this project was provided by Catalan Government (2021 SGR 00920), Spanish Government-FEDER Funds (CPII18/00031, PI20/01424 and CB06/06/0031), the CIBER Enfermedades Respiratorias (CB06/06/0031)), and the European Union’s Horizon 2020 research and innovation program under grant agreement No. 847762 (SMA-TB project). JMGI and CS acknowledge funding from the European Union’s Horizon 2020 research and innovation programme under the Marie Skłodowska-Curie grant agreement No 859962 and No 956148, respectively.

## Author Contributions

J.F. and C.V. conceptualized the study and C.V. acquired the funding, which was managed by her and L-A. N.T., K.B., I.J., M.T., N.B., T.K., Z.A., N.T. and S.V. enrolled participants and oversaw clinical follow-up and samples and data collection at the clinical site. Immune responses data was generated by K.F, L.A, J.M. G-I., and C.S. J.F., J.M. G-I. and C.S. performed the secondary analysis of the data presented here. J.F. and C.V. supervised the project, J.M. G-I., J.F. and C.V. drafted the manuscript. C.V. was responsible for the decision to submit the manuscript. All authors contributed to data interpretation, critical review, and revision of the manuscript, and approved the decision to submit for publication. All authors provided written comments and feedback during manuscript development and were directly involved in the execution of the study.

