## Supplementary Figures for "Baseline host inflammatory and transcriptional profiles associated with structural and functional recovery in drug-resistant tuberculosis"

### Supplementary figures

#### Supplementary Figure 1. ESR and anemia status by outcome.

Risk differences for **a**) high ESR levels and **b**) anemia between participants who achieved or did not achieve each outcome. Confidence intervals were calculated using the Newcombe method.

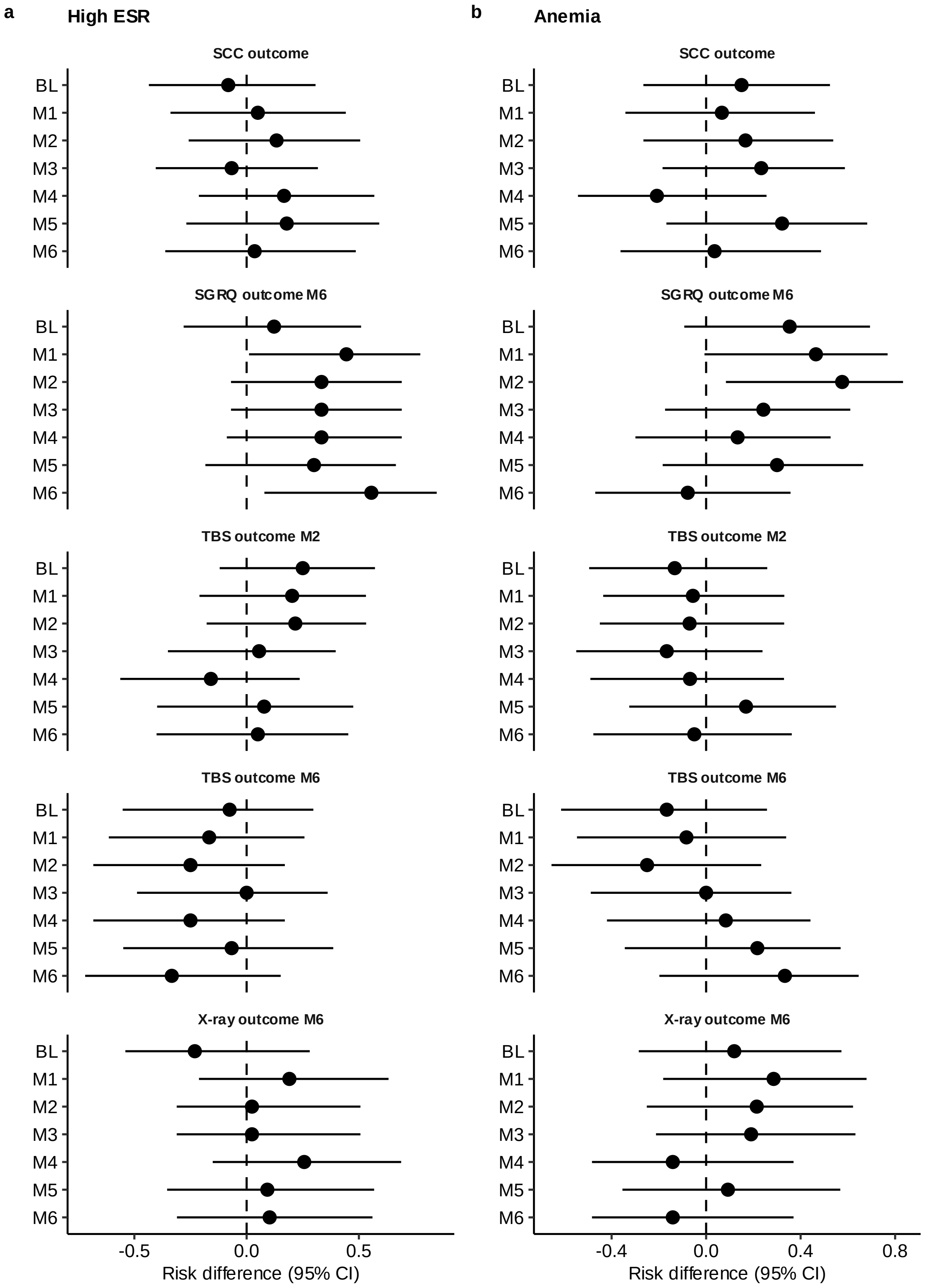

#### Supplementary Figure 2. Baseline differential gene expression between achievers across outcome domains.

**a**) Overlap between baseline differentially expressed gene (DEG) sets. The SGRQ M6 DEG set includes DEGs between achievers and non-achievers of the SGRQ outcome at month 6; the SCC DEG set includes DEGs by SCC outcome; and the X-ray M6 DEG set includes DEGs by X-ray outcome at month 6. **b**) UpSet illustrating the number of overlapping genes among the SGRQ M6, SCC, and X-ray M6 DEG sets, alongside previously published TB signatures (references provided in the main manuscript). **c**-**f**) Longitudinal expression of CD274 and GBP5 between achievers and not achievers of **c, d**) SGRQ outcome M6 and **e, f**) X-ray outcome M6**.** Boxplots show the IQR; lines denote medians. Boxplots show the interquartile range (IQR) with median lines; whiskers extend to 1.5× IQR. Statistical significance at each time point was assessed using DESeq2 (Wald test). Asterisks denote significance levels: ***: adjusted p-value < 0.001, **: adjusted p-value < 0.01 and *: adjusted p-value < 0.05.

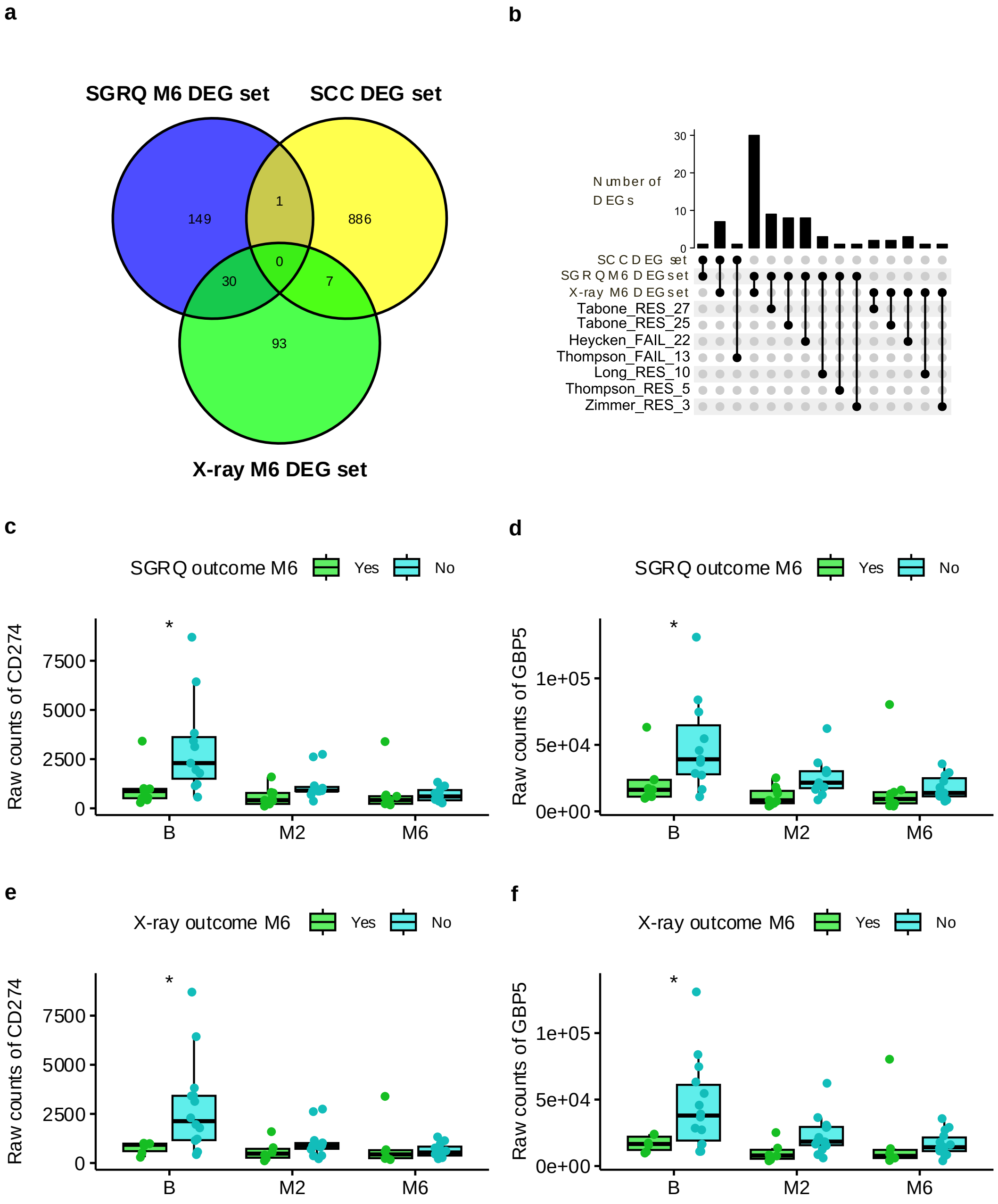

#### Supplementary Figure 3. Baseline ssGSEA scores of DEG sets stratified by outcomes.

ssGSEA scores of SCC DEG set (**a**, **d**, **g**), X-ray M6 DEG set (**b**, **e**, **h**) and SGRQ M6 DEG set (**c**, **f**, **i**) re shown stratified by achievement vs. non-achievement of the following outcomes: SSC outcome (**a**, **b**, **c**), X-ray outcome M6 (**d**, **e**, **f**) and SGRQ outcome M6 (**g**, **h**, **i**). Boxplots show the interquartile range (IQR) with median lines; whiskers extend to 1.5× IQR. Statistical significance was assessed using the Mann–Whitney U test. Asterisks denote significance levels: ***: p-value < 0.001, **: p-value < 0.01, *: p-value < 0.05

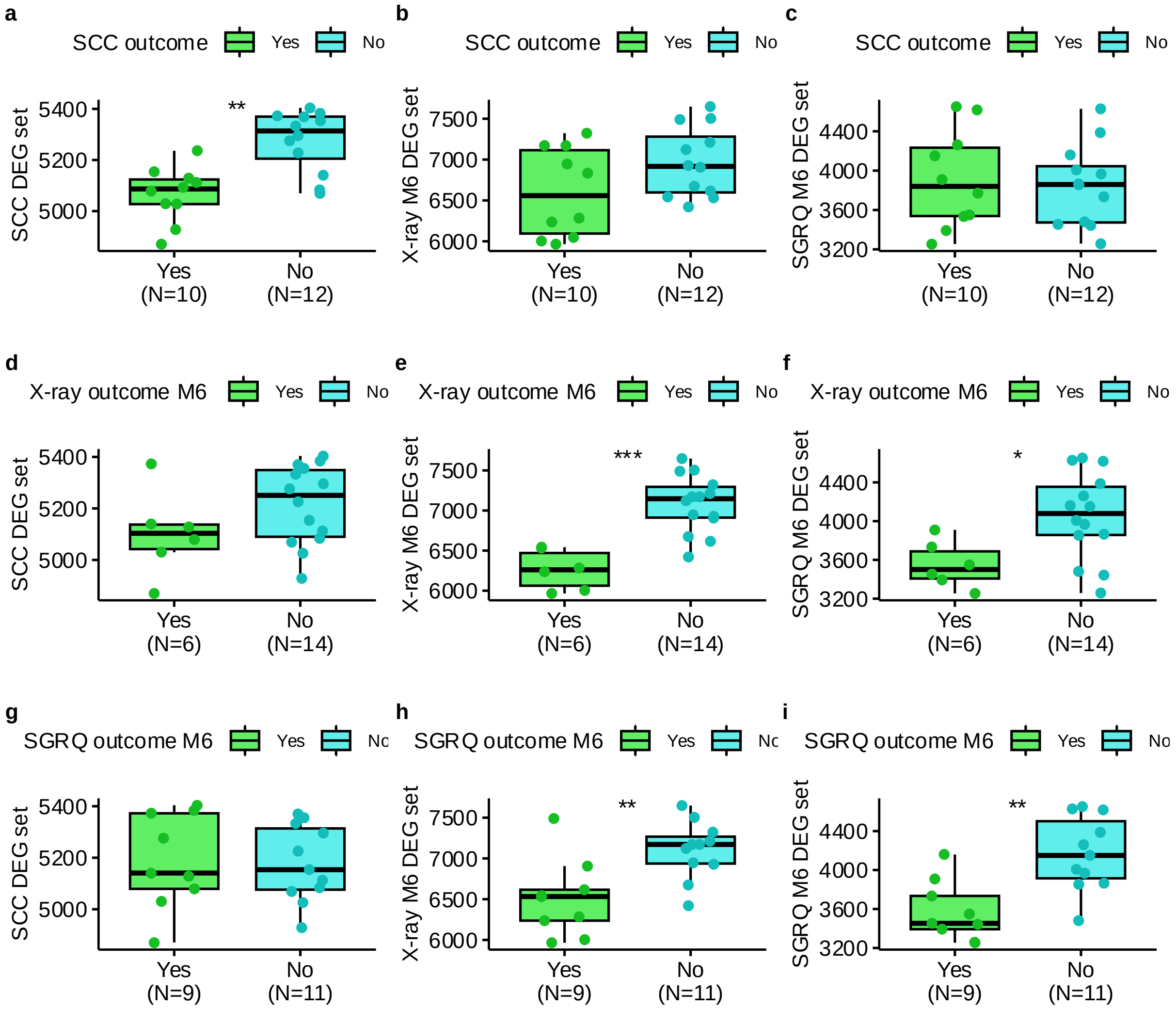

#### Supplementary Figure 4. Baseline ssGSEA scores of established gene signatures stratified by outcome.

Boxplots show the ssGSEA scores of seven previously published blood RNA signatures predictive of TB treatment success (each originally defined by sputum culture conversion status) stratified by **a**) X-ray outcome M6 and **b**) SGRQ outcome M6. Reference for stablished signatures are listed in Table1 of the manuscript. Boxes represent the interquartile range (IQR) with median lines; whiskers extend to 1.5× IQR. Statistical significance was assessed using the Mann–Whitney U test. Asterisks denote significance levels: ***: p-value < 0.001, **: p-value < 0.01, *: p-value < 0.05

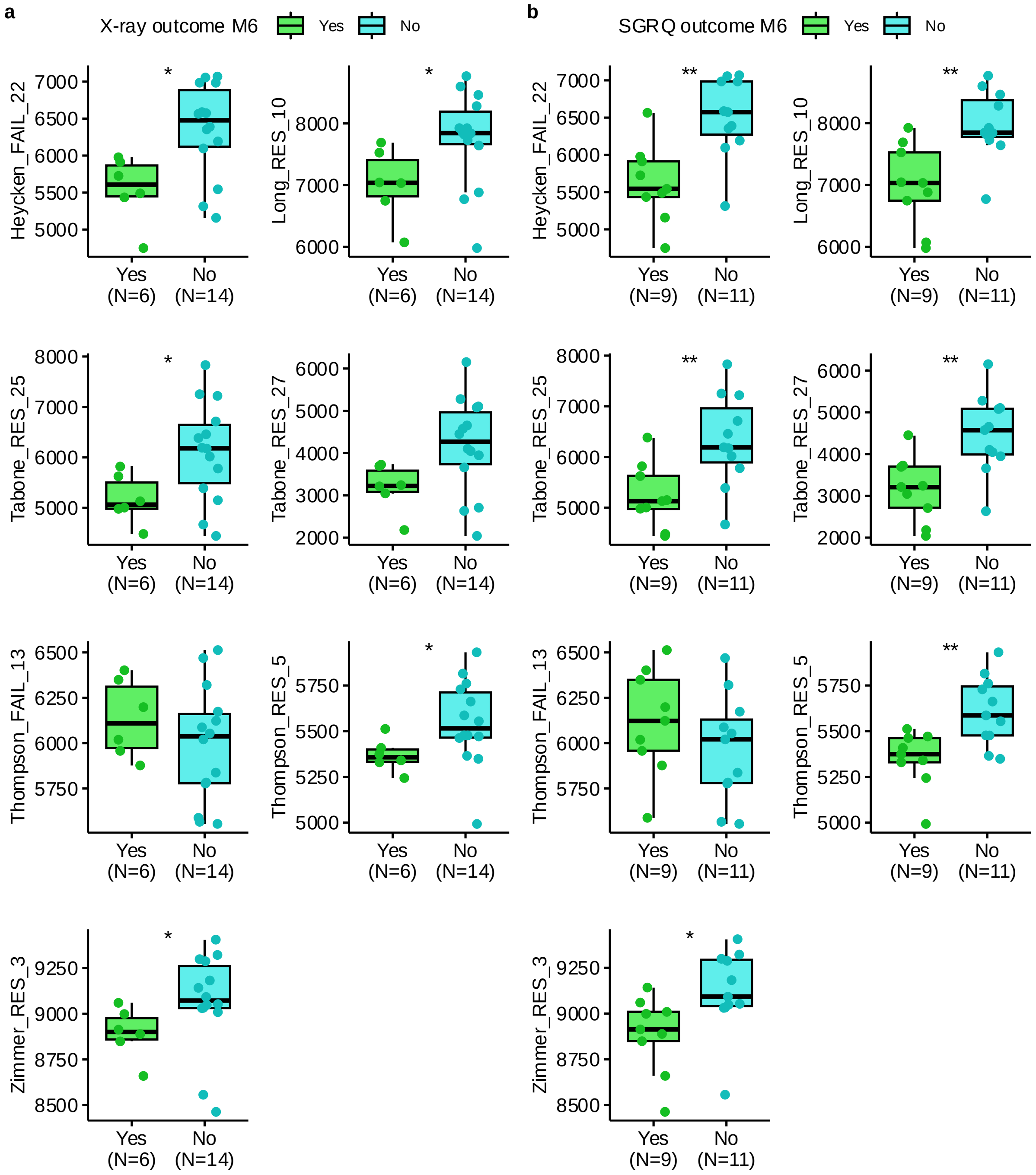

#### Supplementary Figure 5. Evolution of domain scores over the course of treatment

Boxplots show the scores for **a**) SGRQ, **b**) TBS and **c**) X-ray at baseline (BL), month 2 (M2), and month 6 (M6). Boxes represent the interquartile range (IQR) with median lines; whiskers extend to 1.5× IQR. Statistical significance of pairwise comparisons was assessed using Dunn's post-hoc test. Adjusted p-values of the significant pairwise post-hoc tests are shown.

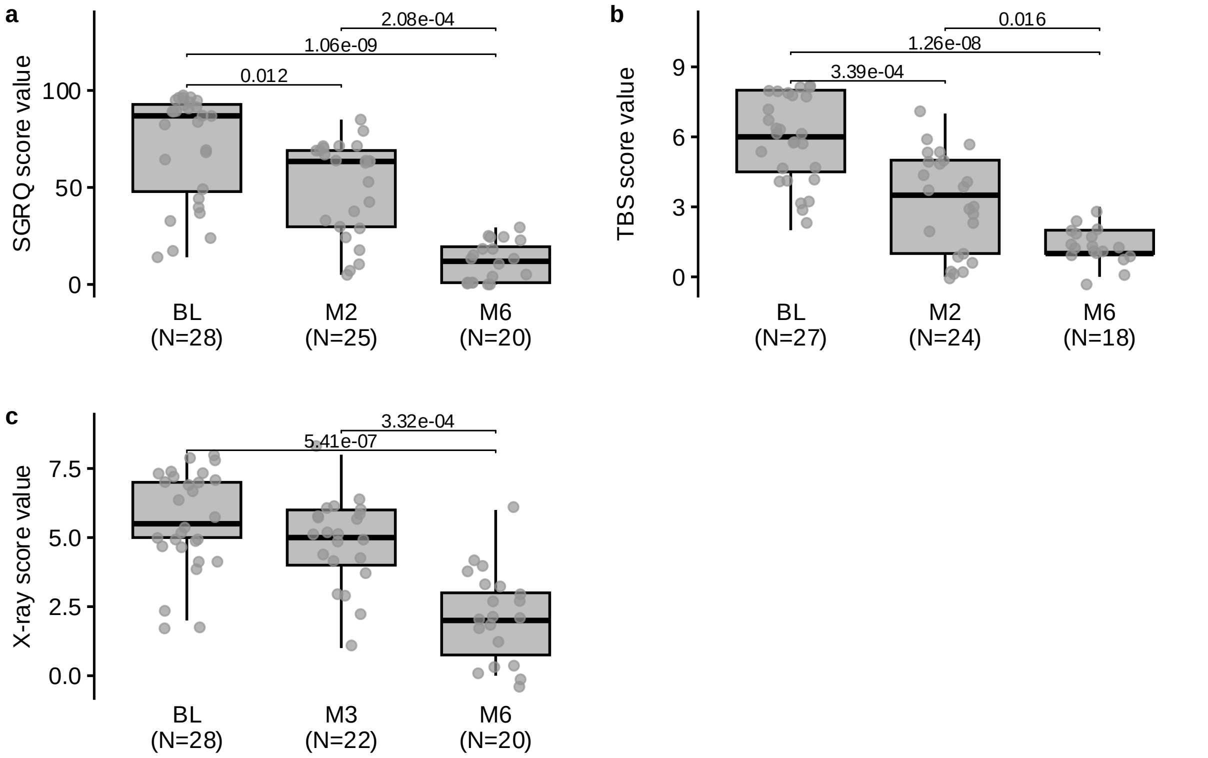

#### Supplementary Figure 6. Evolution of hematological indices over the course of treatment

Boxplots of **a)** hemoglobin (Hb) concentration, **b)** erythrocyte sedimentation rate (ESR), **c)** monocyte-to-lymphocyte ratio (MLR), **d)** neutrophil-to-lymphocyte ratio (NLR), and **e)** systemic inflammation index (SII) at baseline (BL), month 2 (M2), and month 6 (M6). Boxes represent the interquartile range (IQR) with median lines; whiskers extend to 1.5× IQR. Statistical significance of pairwise comparisons was assessed using Dunn's post-hoc test. Adjusted p-values of the significant pairwise post-hoc tests are shown.

**f)** Bar plots showing the incidence of anemia and high ESR across all six measured timepoints; statistical significance was assessed using the chi-square test.

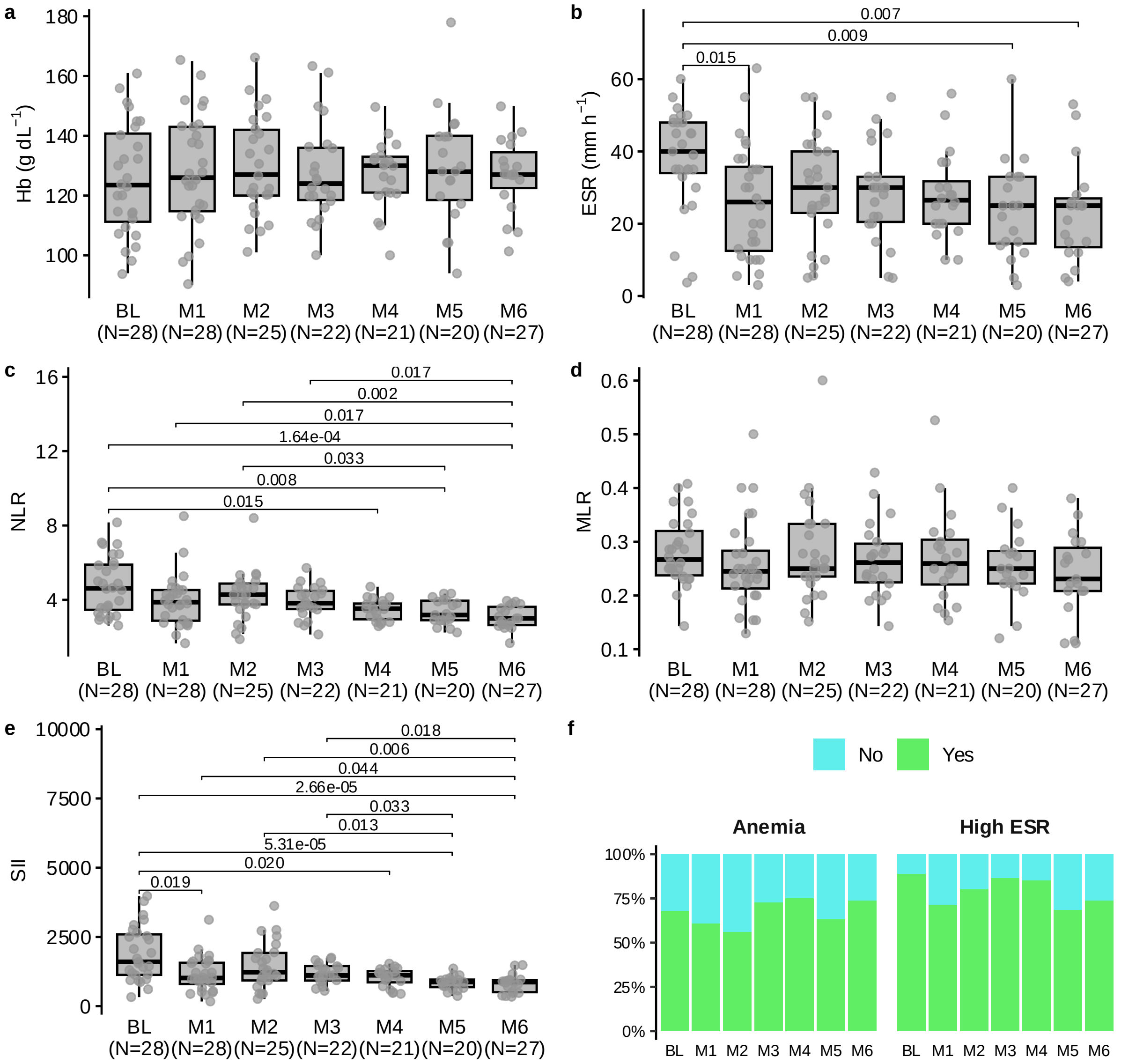

#### Supplementary Figure 7. Evolution of plasma biomarkers over the course of treatment

Boxplots show the concentration of **a)** interferon gamma (INFG), **b)** interleukin 10 (IL-10), **c)** interleukin 12 (IL-12), **d)** interleukin 17A (IL-17A), **e)** interleukin 1B (IL-1B), **f)** interleukin 2 (IL-2), **g**) interleukin 4 (IL-4), **h**) interleukin 6 (IL-6), **i**) interleukin 8 (IL-8) and **j**) tumor necrosis factor (TNF) at baseline (BL), month 2 (M2), and month 6 (M6). Boxes represent the interquartile range (IQR) with median lines; whiskers extend to 1.5× IQR. Statistical significance of pairwise comparisons was assessed using Dunn's post-hoc test. Adjusted p-values of the significant pairwise post-hoc tests are shown.

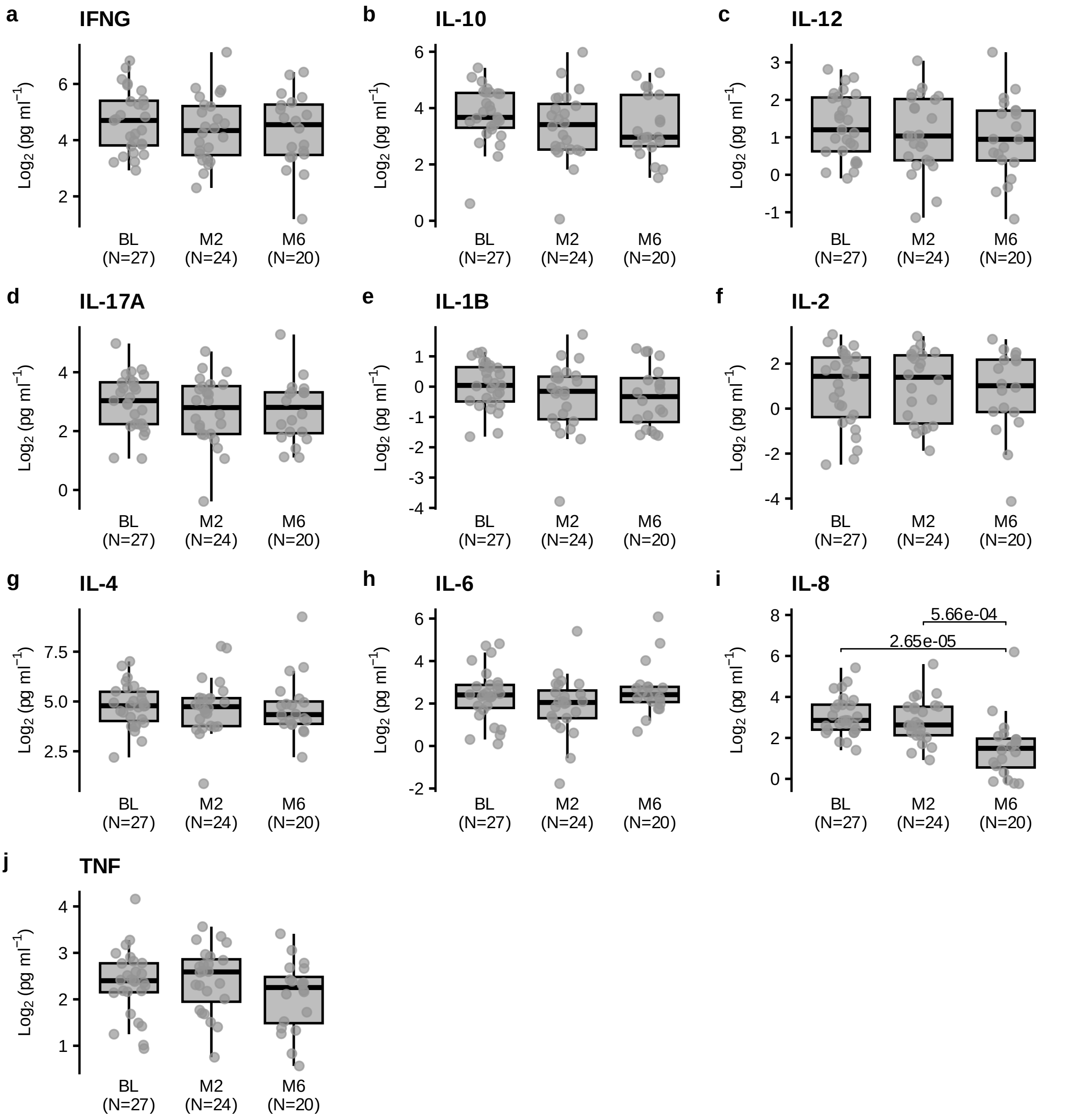

#### Supplementary Figure 8. Evolution of existing gene signatures and DEG sets over the course of treatment

Boxplots show the ssGSEA scores of existing gene signatures **a)** Heycken_FAIL_22, **b)** Long_RES_10, **c)** Tabone_RES_25, **d)** Tabone_RES_27, **e)** Thompson_FAIL_13, **f)** Thompson_RES_5, **g)** Zimmer_RES_3, and DEG sets **h**) SSC DEG set, **i**) X-ray M6 DEG set and **j**) SGRQ DEG set at baseline (BL), month 2 (M2), and month 6 (M6). Boxes represent the interquartile range (IQR) with median lines; whiskers extend to 1.5× IQR. Statistical significance of pairwise comparisons was assessed using Dunn's post-hoc test. Adjusted p-values of the significant pairwise post-hoc tests are shown.

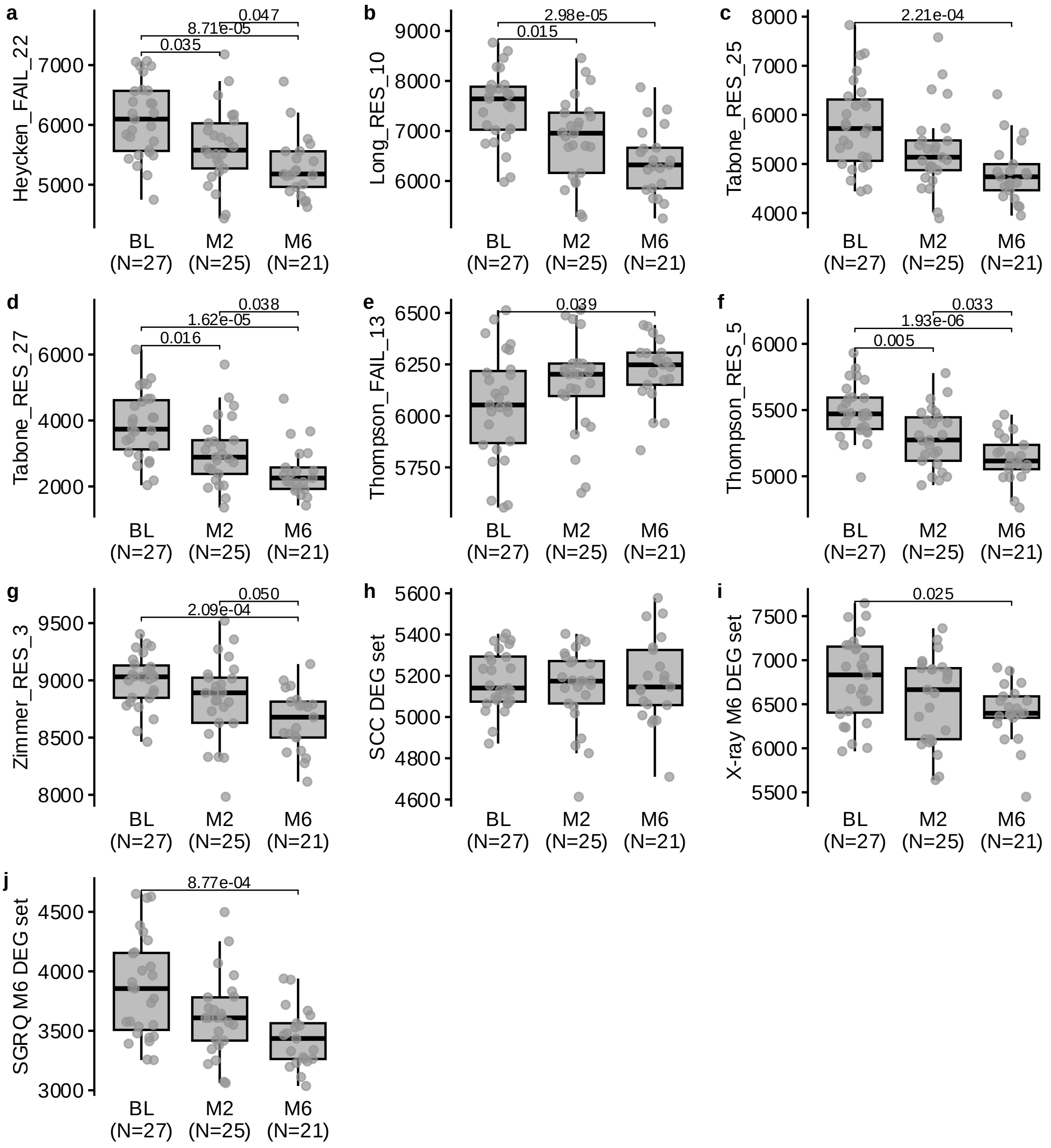
