## Supplementary Tables for "Baseline host inflammatory and transcriptional profiles associated with structural and functional recovery in drug-resistant tuberculosis"

García-Illarramendi et al.

| Table S1 | Outcome achievement per participant |
|---|---|

| Table S2 | SCC DEG set |
|---|---|
| | List of differentially expressed genes (DEGs) between fast converters (conversion within 3 months)and slow converters |

| Table S3 | DEG Sets REATCOME Enrichments |
|---|---|
| **DEG set Name** | **Description** |
| SCC DEG set_Down | Comparison Fast vs Slow converter - Down regulated DEGs in fast converters |
| SCC DEG set_Up | Comparison Fast vs Slow converter - Up regulated DEGs in fast converters |
| X-ray DEG set_Down | Comparison achivement of 75% reduction of X-Ray score at M6 vs not achievemet - Down regulated DEGs in good Xray progressors at M6 |
| SGRQ DEG set_Down | Comparison reaching a SGRQ score in the normal range at M6 or not - Down regulated DEGs in good SGRQ progressors at M6 |
| SGRQ DEG set_Up | Comparison reaching a SGRQ score in the normal range at M6 or not - Up regulated DEGs in good SGRQ progressors at M6 |
| Reactome Set Size | Total number of genes in the REACTOME pathway |
| InSet Intersection Size | Number of genes from the DEG set present in the pathway |
| FDR (adj p-value) | Corrected p value |
| Gene InSet Intersection | List of DEG Set genes that belong to the pathway |

| Table S4 | X-ray DEG set |
|---|---|
| | List of differentially expressed genes (DEGs) between between good Xray progressors at M6 (Reduced X-Ray score 75% respect to baseline) and poor progressors |

| Table S5 | SGRQ DEG set |
|---|---|
| | List of differentially expressed genes (DEGs) between good SGRQ progressors at M6 (attained a normal score) and poor progressors |

| Table S6 | Overlap between X-ray DEG set and SGRQ DEG set |
|---|---|
| Log FC SGRQ outcome M6 | Log fold change between good responders and bad responders for SGRQ outcome M6 |
| Log FC X-ray outome M6 | Log fold change between good responders and bad responders for X-ray outcome M6 |
| Reactome - Immune System | Tick on those belonging to the pathway |
| Reactome - Interferon gamma signaling | Tick on those belonging to the pathway |

| Table S7 | List of genes overlaping between existing gene sigantures of TB response and DEG sets. |
|---|---|
| Sum -Public signatures | Counts the number of times the gene is present in the Existing TB signatures of response |
| Sum- Public sign + DEG Sets | Counts the number of times the gene is present in the Existing TB signatures of response and in the DEG Sets |

| Table S8 | Correlation values observed between the clinical, hematological, plasma biomarker and ssGSEA score values |
|---|---|
| Variable Name (diagonal) | Name of the variable displayed diagonally in the heatmap |
| Variable Name (vertical) | Name of the variable displayed vertically in the heatmap |
| Spearman's rank correlation | Spearman's rank correlation value |
| Adjusted p-value | Adjusted p-value by FDR |

| Participant ID | SCC ≤M3 | TBS 50% reduction M2 | TBS 75% reduction M6 | XRay 50% reduction M3 | XRay 75% reduction M6 | SGRQ Normal Range M2 | SGRQ Normal Range M6 |
|---|---|---|---|---|---|---|---|
| NSAIDS-XDR-TB-CT-01 | Yes | Yes | No | No | Yes | Poor | Normal |
| NSAIDS-XDR-TB-CT-02 | No | Yes | No | No | No | Poor | Poor |
| NSAIDS-XDR-TB-CT-03 | No | No | Yes | No | Yes | Poor | Normal |
| NSAIDS-XDR-TB-CT-04 | No | Yes | Yes | No | No | Poor | Normal |
| NSAIDS-XDR-TB-CT-05 | Yes | Yes | Yes | No | No | Poor | Poor |
| NSAIDS-XDR-TB-CT-06 | Yes | Yes | | No | | Normal | |
| NSAIDS-XDR-TB-CT-07 | | | | | | | |
| NSAIDS-XDR-TB-CT-08 | No | Yes | Yes | No | No | Poor | Poor |
| NSAIDS-XDR-TB-CT-09 | Yes | Yes | | Yes | No | Poor | Poor |
| NSAIDS-XDR-TB-CT-10 | | Yes | | | | Normal | |
| NSAIDS-XDR-TB-CT-11 | | | | | | | |
| NSAIDS-XDR-TB-CT-12 | Yes | No | Yes | No | Yes | Poor | Normal |
| NSAIDS-XDR-TB-CT-13 | No | No | No | No | Yes | Poor | Normal |
| NSAIDS-XDR-TB-CT-14 | No | Yes | No | No | No | Poor | Normal |
| NSAIDS-XDR-TB-IBU-01 | No | No | Yes | No | No | Poor | Normal |
| NSAIDS-XDR-TB-IBU-02 | No | Yes | Yes | No | No | Poor | Poor |
| NSAIDS-XDR-TB-IBU-03 | Yes | No | Yes | No | No | Poor | Poor |
| NSAIDS-XDR-TB-IBU-04 | Yes | Yes | Yes | No | No | Poor | Poor |
| NSAIDS-XDR-TB-IBU-05 | | No | | | | Poor | |
| NSAIDS-XDR-TB-IBU-06 | No | Yes | Yes | No | No | Poor | Poor |
| NSAIDS-XDR-TB-IBU-07 | | | | | | | |
| NSAIDS-XDR-TB-IBU-08 | | No | | | | Poor | |
| NSAIDS-XDR-TB-IBU-09 | Yes | No | | No | | Poor | |
| NSAIDS-XDR-TB-IBU-10 | No | No | Yes | No | No | Poor | Poor |
| NSAIDS-XDR-TB-IBU-11 | No | | | No | No | Poor | Poor |
| NSAIDS-XDR-TB-IBU-12 | Yes | No | No | No | Yes | Poor | Normal |
| NSAIDS-XDR-TB-IBU-13 | Yes | No | No | No | Yes | Poor | Normal |
| NSAIDS-XDR-TB-IBU-14 | No | Yes | Yes | No | No | Poor | Poor |

| SCC DEG set | | | | | | | |
|---|---|---|---|---|---|---|---|
| Ensembl ID | Gene name | baseMean | Log FC | Log FC SE | Wald statistic value | P-value | Adjusted p-value |
| ENSG00000157426.14 | AASDH | 202.07 | -0.27 | 0.08 | -3.25 | 0.00 | 0.03 |
| ENSG00000008311.16 | AASS | 99.93 | -0.59 | 0.20 | -2.97 | 0.00 | 0.05 |
| ENSG00000183044.12 | ABAT | 1014.37 | 0.40 | 0.13 | 3.13 | 0.00 | 0.04 |
| ENSG00000154263.18 | ABCA10 | 25.43 | -0.51 | 0.17 | -3.01 | 0.00 | 0.04 |
| ENSG00000154265.16 | ABCA5 | 669.45 | -0.45 | 0.12 | -3.75 | 0.00 | 0.01 |
| ENSG00000140526.18 | ABHD2 | 15379.95 | 0.44 | 0.14 | 3.14 | 0.00 | 0.04 |
| ENSG00000163322.14 | ABRAXAS1 | 298.61 | -0.43 | 0.14 | -3.01 | 0.00 | 0.04 |
| ENSG00000114626.18 | ABTB1 | 13618.48 | 0.56 | 0.15 | 3.73 | 0.00 | 0.01 |
| ENSG00000117054.15 | ACADM | 615.57 | -0.42 | 0.10 | -4.08 | 0.00 | 0.01 |
| ENSG00000130402.13 | ACTN4 | 8379.97 | 0.38 | 0.11 | 3.48 | 0.00 | 0.02 |
| ENSG00000075089.10 | ACTR6 | 88.95 | -0.47 | 0.15 | -3.04 | 0.00 | 0.04 |
| ENSG00000093072.18 | ADA2 | 7249.13 | 0.30 | 0.10 | 2.92 | 0.00 | 0.05 |
| ENSG00000143537.14 | ADAM15 | 852.32 | 0.33 | 0.10 | 3.43 | 0.00 | 0.02 |
| ENSG00000151651.16 | ADAM8 | 12757.04 | 0.55 | 0.17 | 3.21 | 0.00 | 0.03 |
| ENSG00000143382.16 | ADAMTSL4 | 1165.53 | 0.44 | 0.14 | 3.19 | 0.00 | 0.03 |
| ENSG00000131355.15 | ADGRE3 | 5019.55 | 0.50 | 0.17 | 2.94 | 0.00 | 0.05 |
| ENSG00000123146.20 | ADGRE5 | 17394.45 | 0.49 | 0.16 | 3.13 | 0.00 | 0.04 |
| ENSG00000170222.12 | ADPRM | 156.98 | -0.48 | 0.12 | -4.12 | 0.00 | 0.01 |
| ENSG00000204305.16 | AGER | 587.57 | 0.50 | 0.16 | 3.08 | 0.00 | 0.04 |
| ENSG00000123908.12 | AGO2 | 6713.82 | 0.38 | 0.12 | 3.24 | 0.00 | 0.03 |
| ENSG00000135541.22 | AHI1 | 261.43 | -0.81 | 0.25 | -3.26 | 0.00 | 0.03 |
| ENSG00000106992.19 | AK1 | 439.74 | 0.89 | 0.23 | 3.84 | 0.00 | 0.01 |
| ENSG00000155085.16 | AK9 | 235.98 | -0.42 | 0.13 | -3.16 | 0.00 | 0.03 |
| ENSG00000023516.9 | AKAP11 | 1991.72 | -0.30 | 0.10 | -2.94 | 0.00 | 0.05 |
| ENSG00000197976.12 | AKAP17A | 2239.22 | 0.30 | 0.09 | 3.43 | 0.00 | 0.02 |
| ENSG00000179841.8 | AKAP5 | 48.98 | -1.13 | 0.32 | -3.54 | 0.00 | 0.02 |
| ENSG00000127914.19 | AKAP9 | 3126.36 | -0.45 | 0.09 | -4.80 | 0.00 | 0.00 |
| ENSG00000149925.22 | ALDOA | 13521.90 | 0.41 | 0.13 | 3.11 | 0.00 | 0.04 |
| ENSG00000175548.9 | ALG10B | 301.61 | -0.46 | 0.15 | -3.10 | 0.00 | 0.04 |
| ENSG00000088035.18 | ALG6 | 345.68 | -0.22 | 0.07 | -3.08 | 0.00 | 0.04 |
| ENSG00000137760.15 | ALKBH8 | 173.90 | -0.53 | 0.15 | -3.59 | 0.00 | 0.01 |
| ENSG00000164162.14 | ANAPC10 | 56.62 | -0.48 | 0.15 | -3.15 | 0.00 | 0.03 |
| ENSG00000053900.11 | ANAPC4 | 513.05 | -0.25 | 0.08 | -2.97 | 0.00 | 0.05 |
| ENSG00000174606.14 | ANGEL2 | 931.15 | -0.30 | 0.09 | -3.22 | 0.00 | 0.03 |
| ENSG00000151687.15 | ANKAR | 124.43 | -0.50 | 0.17 | -2.99 | 0.00 | 0.04 |
| ENSG00000106524.9 | ANKMY2 | 144.93 | -0.34 | 0.10 | -3.59 | 0.00 | 0.01 |
| ENSG00000172932.16 | ANKRD13D | 4082.34 | 0.45 | 0.12 | 3.79 | 0.00 | 0.01 |
| ENSG00000135976.21 | ANKRD36 | 1037.16 | -0.63 | 0.18 | -3.49 | 0.00 | 0.02 |
| ENSG00000174501.15 | ANKRD36C | 864.50 | -0.74 | 0.19 | -3.96 | 0.00 | 0.01 |
| ENSG00000168876.9 | ANKRD49 | 390.67 | -0.37 | 0.12 | -3.18 | 0.00 | 0.03 |
| ENSG00000166825.15 | ANPEP | 12993.89 | 0.82 | 0.19 | 4.21 | 0.00 | 0.00 |
| ENSG00000122359.18 | ANXA11 | 9815.83 | 0.30 | 0.10 | 2.93 | 0.00 | 0.05 |
| ENSG00000131480.9 | AOC2 | 472.58 | 0.63 | 0.16 | 3.84 | 0.00 | 0.01 |
| ENSG00000131471.7 | AOC3 | 285.70 | 0.75 | 0.22 | 3.35 | 0.00 | 0.02 |
| ENSG00000183020.15 | AP2A2 | 1337.08 | 0.20 | 0.05 | 4.02 | 0.00 | 0.01 |
| ENSG00000065000.19 | AP3D1 | 2740.03 | 0.22 | 0.06 | 3.57 | 0.00 | 0.02 |
| ENSG00000185009.13 | AP3M1 | 1169.86 | -0.18 | 0.05 | -3.30 | 0.00 | 0.03 |
| ENSG00000157500.12 | APPL1 | 1027.87 | -0.26 | 0.08 | -3.11 | 0.00 | 0.04 |
| ENSG00000021776.11 | AQR | 1803.95 | -0.18 | 0.06 | -3.04 | 0.00 | 0.04 |
| ENSG00000186635.15 | ARAP1 | 18017.27 | 0.47 | 0.13 | 3.70 | 0.00 | 0.01 |
| ENSG00000120318.16 | ARAP3 | 3015.22 | 0.67 | 0.17 | 4.05 | 0.00 | 0.01 |
| ENSG00000004059.11 | ARF5 | 2080.50 | 0.32 | 0.11 | 2.96 | 0.00 | 0.05 |
| ENSG00000134884.15 | ARGLU1 | 4271.46 | -0.28 | 0.09 | -3.23 | 0.00 | 0.03 |
| ENSG00000175220.12 | ARHGAP1 | 3491.15 | 0.38 | 0.09 | 4.10 | 0.00 | 0.01 |

| SCC DEG set | | | | | | | |
|---|---|---|---|---|---|---|---|
| Ensembl ID | Gene name | baseMean | Log FC | Log FC SE | Wald statistic value | P-value | Adjusted p-value |
| ENSG00000165322.18 | ARHGAP12 | 545.69 | -0.45 | 0.12 | -3.76 | 0.00 | 0.01 |
| ENSG00000163219.12 | ARHGAP25 | 9485.87 | 0.38 | 0.12 | 3.18 | 0.00 | 0.03 |
| ENSG00000159314.12 | ARHGAP27 | 9047.06 | 0.39 | 0.13 | 3.15 | 0.00 | 0.03 |
| ENSG00000089820.16 | ARHGAP4 | 5697.58 | 0.29 | 0.09 | 3.27 | 0.00 | 0.03 |
| ENSG00000100852.14 | ARHGAP5 | 940.23 | -0.38 | 0.10 | -3.99 | 0.00 | 0.01 |
| ENSG00000076928.18 | ARHGEF1 | 11977.41 | 0.34 | 0.10 | 3.52 | 0.00 | 0.02 |
| ENSG00000116584.21 | ARHGEF2 | 8206.98 | 0.30 | 0.09 | 3.48 | 0.00 | 0.02 |
| ENSG00000117713.21 | ARID1A | 9296.85 | 0.32 | 0.10 | 3.18 | 0.00 | 0.03 |
| ENSG00000162980.17 | ARL5A | 1059.59 | -0.35 | 0.11 | -3.16 | 0.00 | 0.03 |
| ENSG00000113966.10 | ARL6 | 44.37 | -0.74 | 0.23 | -3.25 | 0.00 | 0.03 |
| ENSG00000197070.14 | ARRDC1 | 1479.73 | 0.31 | 0.10 | 3.28 | 0.00 | 0.03 |
| ENSG00000100299.18 | ARSA | 1658.92 | 0.39 | 0.10 | 3.79 | 0.00 | 0.01 |
| ENSG00000100325.15 | ASCC2 | 8255.35 | 0.89 | 0.27 | 3.34 | 0.00 | 0.02 |
| ENSG00000213676.13 | ATF6B | 2484.30 | 0.26 | 0.07 | 3.57 | 0.00 | 0.02 |
| ENSG00000168010.11 | ATG16L2 | 17397.69 | 0.47 | 0.14 | 3.37 | 0.00 | 0.02 |
| ENSG00000168397.17 | ATG4B | 971.98 | 0.25 | 0.08 | 3.31 | 0.00 | 0.03 |
| ENSG00000057663.16 | ATG5 | 337.78 | -0.41 | 0.13 | -3.15 | 0.00 | 0.03 |
| ENSG00000175054.16 | ATR | 2038.54 | -0.31 | 0.08 | -3.79 | 0.00 | 0.01 |
| ENSG00000175711.9 | B3GNTL1 | 895.73 | 0.62 | 0.13 | 4.71 | 0.00 | 0.00 |
| ENSG00000112208.11 | BAG2 | 120.34 | -0.50 | 0.16 | -3.09 | 0.00 | 0.04 |
| ENSG00000076108.12 | BAZ2A | 9992.08 | 0.37 | 0.12 | 3.00 | 0.00 | 0.04 |
| ENSG00000107949.17 | BCCIP | 228.85 | -0.41 | 0.12 | -3.53 | 0.00 | 0.02 |
| ENSG00000103507.14 | BCKDK | 1116.16 | 0.34 | 0.10 | 3.42 | 0.00 | 0.02 |
| ENSG00000085185.16 | BCORL1 | 1404.58 | 0.48 | 0.14 | 3.42 | 0.00 | 0.02 |
| ENSG00000145734.20 | BDP1 | 1479.95 | -0.45 | 0.11 | -3.98 | 0.00 | 0.01 |
| ENSG00000023445.16 | BIRC3 | 2133.31 | -0.55 | 0.17 | -3.30 | 0.00 | 0.03 |
| ENSG00000090013.11 | BLVRB | 3234.78 | 1.04 | 0.33 | 3.12 | 0.00 | 0.04 |
| ENSG00000168283.14 | BMI1 | 550.37 | -0.34 | 0.11 | -3.04 | 0.00 | 0.04 |
| ENSG00000104331.9 | BPNT2 | 1230.53 | -0.26 | 0.09 | -2.95 | 0.00 | 0.05 |
| ENSG00000141867.19 | BRD4 | 9712.00 | 0.48 | 0.13 | 3.66 | 0.00 | 0.01 |
| ENSG00000185658.14 | BRWD1 | 3419.85 | -0.29 | 0.07 | -4.15 | 0.00 | 0.01 |
| ENSG00000159388.6 | BTG2 | 5567.93 | 0.32 | 0.10 | 3.12 | 0.00 | 0.04 |
| ENSG00000179476.8 | C14orf28 | 83.76 | -0.53 | 0.13 | -3.98 | 0.00 | 0.01 |
| ENSG00000258315.5 | C17orf49 | 588.26 | 0.28 | 0.08 | 3.49 | 0.00 | 0.02 |
| ENSG00000141219.16 | C17orf80 | 318.82 | -0.36 | 0.08 | -4.34 | 0.00 | 0.00 |
| ENSG00000135974.10 | C2orf49 | 581.02 | -0.27 | 0.07 | -4.04 | 0.00 | 0.01 |
| ENSG00000134830.6 | C5AR2 | 3940.67 | 0.48 | 0.12 | 4.05 | 0.00 | 0.01 |
| ENSG00000164241.14 | C5orf63 | 85.01 | -0.75 | 0.21 | -3.51 | 0.00 | 0.02 |
| ENSG00000145349.18 | CAMK2D | 1003.70 | -0.46 | 0.15 | -3.12 | 0.00 | 0.04 |
| ENSG00000108509.21 | CAMTA2 | 2567.29 | 0.37 | 0.11 | 3.48 | 0.00 | 0.02 |
| ENSG00000149260.18 | CAPN5 | 446.75 | 0.91 | 0.27 | 3.35 | 0.00 | 0.02 |
| ENSG00000131375.10 | CAPN7 | 957.63 | -0.37 | 0.09 | -3.95 | 0.00 | 0.01 |
| ENSG00000138380.18 | CARF | 325.90 | -0.47 | 0.15 | -3.10 | 0.00 | 0.04 |
| ENSG00000118412.13 | CASP8AP2 | 849.50 | -0.49 | 0.11 | -4.26 | 0.00 | 0.00 |
| ENSG00000087589.17 | CASS4 | 2132.66 | 0.52 | 0.14 | 3.76 | 0.00 | 0.01 |
| ENSG00000110395.7 | CBL | 11607.94 | 0.55 | 0.15 | 3.76 | 0.00 | 0.01 |
| ENSG00000132024.18 | CC2D1A | 731.51 | 0.22 | 0.07 | 3.17 | 0.00 | 0.03 |
| ENSG00000164221.13 | CCDC112 | 91.87 | -0.56 | 0.17 | -3.32 | 0.00 | 0.03 |
| ENSG00000152492.15 | CCDC50 | 652.31 | -0.45 | 0.14 | -3.29 | 0.00 | 0.03 |
| ENSG00000133773.12 | CCDC59 | 178.83 | -0.44 | 0.12 | -3.51 | 0.00 | 0.02 |
| ENSG00000180376.17 | CCDC66 | 375.26 | -0.45 | 0.14 | -3.26 | 0.00 | 0.03 |
| ENSG00000149231.16 | CCDC82 | 1348.96 | -0.43 | 0.09 | -4.69 | 0.00 | 0.00 |
| ENSG00000168071.22 | CCDC88B | 6120.82 | 0.47 | 0.13 | 3.64 | 0.00 | 0.01 |
| ENSG00000137500.10 | CCDC90B | 441.54 | -0.30 | 0.09 | -3.57 | 0.00 | 0.02 |

| SCC DEG set | | | | | | | |
|---|---|---|---|---|---|---|---|
| Ensembl ID | Gene name | baseMean | Log FC | Log FC SE | Wald statistic value | P-value | Adjusted p-value |
| ENSG00000123106.11 | CCDC91 | 356.05 | -0.33 | 0.10 | -3.30 | 0.00 | 0.03 |
| ENSG00000107771.17 | CCSER2 | 1813.53 | -0.31 | 0.10 | -3.16 | 0.00 | 0.03 |
| ENSG00000178789.9 | CD300LB | 1392.18 | 0.42 | 0.13 | 3.30 | 0.00 | 0.03 |
| ENSG00000004468.13 | CD38 | 834.90 | -0.62 | 0.18 | -3.49 | 0.00 | 0.02 |
| ENSG00000196776.17 | CD47 | 6235.60 | -0.20 | 0.07 | -2.96 | 0.00 | 0.05 |
| ENSG00000106993.12 | CDC37L1 | 153.57 | -0.44 | 0.13 | -3.29 | 0.00 | 0.03 |
| ENSG00000149798.5 | CDC42EP2 | 1892.69 | 0.56 | 0.19 | 3.04 | 0.00 | 0.04 |
| ENSG00000145996.11 | CDKAL1 | 419.85 | -0.39 | 0.10 | -3.94 | 0.00 | 0.01 |
| ENSG00000168564.6 | CDKN2AIP | 431.28 | -0.25 | 0.08 | -3.13 | 0.00 | 0.04 |
| ENSG00000151849.16 | CENPJ | 108.32 | -0.45 | 0.13 | -3.39 | 0.00 | 0.02 |
| ENSG00000168944.17 | CEP120 | 1189.05 | -0.34 | 0.08 | -4.31 | 0.00 | 0.00 |
| ENSG00000133393.13 | CEP20 | 282.86 | -0.27 | 0.08 | -3.25 | 0.00 | 0.03 |
| ENSG00000198707.17 | CEP290 | 448.70 | -0.56 | 0.16 | -3.59 | 0.00 | 0.01 |
| ENSG00000166004.15 | CEP295 | 932.51 | -0.23 | 0.06 | -3.60 | 0.00 | 0.01 |
| ENSG00000166037.11 | CEP57 | 597.36 | -0.51 | 0.13 | -3.91 | 0.00 | 0.01 |
| ENSG00000148019.14 | CEP78 | 882.82 | -0.62 | 0.21 | -3.00 | 0.00 | 0.04 |
| ENSG00000173588.16 | CEP83 | 132.67 | -0.58 | 0.14 | -4.26 | 0.00 | 0.00 |
| ENSG00000165410.15 | CFL2 | 108.67 | -0.53 | 0.17 | -3.06 | 0.00 | 0.04 |
| ENSG00000177200.18 | CHD9 | 2326.08 | -0.24 | 0.06 | -3.97 | 0.00 | 0.01 |
| ENSG00000188419.14 | CHM | 437.56 | -0.38 | 0.12 | -3.23 | 0.00 | 0.03 |
| ENSG00000203668.3 | CHML | 447.49 | -0.52 | 0.18 | -2.93 | 0.00 | 0.05 |
| ENSG00000171310.11 | CHST11 | 5184.32 | 0.36 | 0.10 | 3.55 | 0.00 | 0.02 |
| ENSG00000079432.9 | CIC | 5293.10 | 0.36 | 0.10 | 3.66 | 0.00 | 0.01 |
| ENSG00000106404.14 | CLDN15 | 422.28 | 0.31 | 0.10 | 3.00 | 0.00 | 0.04 |
| ENSG00000038532.16 | CLEC16A | 2273.78 | 0.26 | 0.09 | 2.94 | 0.00 | 0.05 |
| ENSG00000069493.15 | CLEC2D | 2541.08 | -0.51 | 0.15 | -3.34 | 0.00 | 0.02 |
| ENSG00000102805.16 | CLN5 | 584.33 | -0.29 | 0.09 | -3.18 | 0.00 | 0.03 |
| ENSG00000162129.14 | CLPB | 651.51 | 0.27 | 0.09 | 3.20 | 0.00 | 0.03 |
| ENSG00000104853.16 | CLPTM1 | 1902.60 | 0.28 | 0.08 | 3.40 | 0.00 | 0.02 |
| ENSG00000198791.12 | CNOT7 | 1344.90 | -0.27 | 0.07 | -3.86 | 0.00 | 0.01 |
| ENSG00000068120.15 | COASY | 947.83 | 0.27 | 0.09 | 3.01 | 0.00 | 0.04 |
| ENSG00000164597.15 | COG5 | 1268.07 | -0.26 | 0.09 | -3.00 | 0.00 | 0.04 |
| ENSG00000133103.17 | COG6 | 276.89 | -0.43 | 0.11 | -3.76 | 0.00 | 0.01 |
| ENSG00000105669.14 | COPE | 2295.65 | 0.29 | 0.09 | 3.22 | 0.00 | 0.03 |
| ENSG00000198612.11 | COPS8 | 284.74 | -0.36 | 0.12 | -3.02 | 0.00 | 0.04 |
| ENSG00000102879.16 | CORO1A | 26523.65 | 0.36 | 0.12 | 3.03 | 0.00 | 0.04 |
| ENSG00000103187.8 | COTL1 | 23887.03 | 0.34 | 0.11 | 2.96 | 0.00 | 0.05 |
| ENSG00000197603.16 | CPLANE1 | 593.79 | -0.35 | 0.10 | -3.48 | 0.00 | 0.02 |
| ENSG00000005339.15 | CREBBP | 13929.66 | 0.41 | 0.14 | 3.02 | 0.00 | 0.04 |
| ENSG00000160741.17 | CRTC2 | 2208.11 | 0.37 | 0.11 | 3.46 | 0.00 | 0.02 |
| ENSG00000205758.12 | CRYZL1 | 321.66 | -0.31 | 0.06 | -4.91 | 0.00 | 0.00 |
| ENSG00000141551.15 | CSNK1D | 6489.93 | 0.32 | 0.11 | 3.01 | 0.00 | 0.04 |
| ENSG00000133275.16 | CSNK1G2 | 3711.30 | 0.28 | 0.09 | 3.26 | 0.00 | 0.03 |
| ENSG00000175215.11 | CTDSP2 | 10863.79 | 0.43 | 0.12 | 3.62 | 0.00 | 0.01 |
| ENSG00000196188.12 | CTSE | 49.21 | 1.32 | 0.41 | 3.22 | 0.00 | 0.03 |
| ENSG00000166266.14 | CUL5 | 753.07 | -0.38 | 0.12 | -3.18 | 0.00 | 0.03 |
| ENSG00000153015.17 | CWC27 | 988.31 | -0.36 | 0.11 | -3.24 | 0.00 | 0.03 |
| ENSG00000152404.16 | CWF19L2 | 362.88 | -0.59 | 0.14 | -4.21 | 0.00 | 0.00 |
| ENSG00000154832.16 | CXXC1 | 812.32 | 0.24 | 0.08 | 3.09 | 0.00 | 0.04 |
| ENSG00000186204.15 | CYP4F12 | 143.99 | 0.67 | 0.21 | 3.13 | 0.00 | 0.04 |
| ENSG00000145476.16 | CYP4V2 | 667.77 | -0.49 | 0.15 | -3.29 | 0.00 | 0.03 |
| ENSG00000100592.16 | DAAM1 | 314.56 | -0.32 | 0.11 | -2.94 | 0.00 | 0.05 |
| ENSG00000164535.15 | DAGLB | 699.19 | 0.23 | 0.06 | 4.18 | 0.00 | 0.00 |
| ENSG00000164934.14 | DCAF13 | 326.34 | -0.46 | 0.12 | -3.93 | 0.00 | 0.01 |

| SCC DEG set | | | | | | | |
|---|---|---|---|---|---|---|---|
| Ensembl ID | Gene name | baseMean | Log FC | Log FC SE | Wald statistic value | P-value | Adjusted p-value |
| ENSG00000164465.19 | DCBLD1 | 101.22 | -0.49 | 0.16 | -3.10 | 0.00 | 0.04 |
| ENSG00000109184.15 | DCUN1D4 | 310.31 | -0.42 | 0.10 | -4.36 | 0.00 | 0.00 |
| ENSG00000100523.16 | DDHD1 | 2307.67 | -0.41 | 0.09 | -4.62 | 0.00 | 0.00 |
| ENSG00000079785.16 | DDX1 | 539.94 | -0.30 | 0.10 | -3.10 | 0.00 | 0.04 |
| ENSG00000178105.12 | DDX10 | 342.76 | -0.46 | 0.13 | -3.42 | 0.00 | 0.02 |
| ENSG00000064703.13 | DDX20 | 353.41 | -0.31 | 0.09 | -3.46 | 0.00 | 0.02 |
| ENSG00000107625.14 | DDX50 | 292.49 | -0.37 | 0.13 | -2.92 | 0.00 | 0.05 |
| ENSG00000278053.5 | DDX52 | 569.83 | -0.25 | 0.07 | -3.60 | 0.00 | 0.01 |
| ENSG00000140995.17 | DEF8 | 3488.24 | 0.45 | 0.14 | 3.24 | 0.00 | 0.03 |
| ENSG00000105339.11 | DENND3 | 18292.76 | 0.44 | 0.14 | 3.06 | 0.00 | 0.04 |
| ENSG00000198837.10 | DENND4B | 4611.37 | 0.33 | 0.09 | 3.67 | 0.00 | 0.01 |
| ENSG00000102780.17 | DGKH | 648.68 | -0.33 | 0.10 | -3.33 | 0.00 | 0.02 |
| ENSG00000149091.15 | DGKZ | 5691.28 | 0.31 | 0.10 | 3.19 | 0.00 | 0.03 |
| ENSG00000174953.14 | DHX36 | 1316.84 | -0.33 | 0.07 | -4.84 | 0.00 | 0.00 |
| ENSG00000083520.15 | DIS3 | 1145.13 | -0.32 | 0.08 | -4.11 | 0.00 | 0.01 |
| ENSG00000075711.21 | DLG1 | 1136.02 | -0.30 | 0.08 | -3.81 | 0.00 | 0.01 |
| ENSG00000135164.19 | DMTF1 | 2356.36 | -0.20 | 0.07 | -2.94 | 0.00 | 0.05 |
| ENSG00000164031.17 | DNAJB14 | 2338.89 | -0.37 | 0.12 | -3.01 | 0.00 | 0.04 |
| ENSG00000105821.15 | DNAJC2 | 225.33 | -0.37 | 0.12 | -3.15 | 0.00 | 0.03 |
| ENSG00000168724.17 | DNAJC21 | 454.68 | -0.35 | 0.12 | -2.98 | 0.00 | 0.05 |
| ENSG00000167968.13 | DNASE1L2 | 36.59 | 0.66 | 0.18 | 3.65 | 0.00 | 0.01 |
| ENSG00000136908.18 | DPM2 | 2331.02 | 0.93 | 0.31 | 3.03 | 0.00 | 0.04 |
| ENSG00000156162.16 | DPY19L4 | 217.39 | -0.42 | 0.14 | -3.06 | 0.00 | 0.04 |
| ENSG00000104047.15 | DTWD1 | 297.00 | -0.31 | 0.10 | -3.15 | 0.00 | 0.03 |
| ENSG00000184786.6 | DYNLT2 | 38.31 | -0.72 | 0.18 | -3.96 | 0.00 | 0.01 |
| ENSG00000205250.9 | E2F4 | 1945.12 | 0.43 | 0.11 | 3.79 | 0.00 | 0.01 |
| ENSG00000133740.11 | E2F5 | 72.27 | -0.75 | 0.24 | -3.08 | 0.00 | 0.04 |
| ENSG00000038358.15 | EDC4 | 1413.30 | 0.25 | 0.08 | 3.06 | 0.00 | 0.04 |
| ENSG00000102189.17 | EEA1 | 783.35 | -0.35 | 0.10 | -3.62 | 0.00 | 0.01 |
| ENSG00000203791.15 | EEF1AKMT2 | 206.97 | -0.39 | 0.09 | -4.23 | 0.00 | 0.00 |
| ENSG00000104529.19 | EEF1D | 4103.32 | 0.27 | 0.08 | 3.60 | 0.00 | 0.01 |
| ENSG00000172638.13 | EFEMP2 | 96.48 | 0.53 | 0.18 | 2.94 | 0.00 | 0.05 |
| ENSG00000115504.15 | EHBP1 | 354.45 | -0.30 | 0.10 | -2.99 | 0.00 | 0.04 |
| ENSG00000173442.13 | EHBP1L1 | 8779.27 | 0.47 | 0.13 | 3.61 | 0.00 | 0.01 |
| ENSG00000148730.7 | EIF4EBP2 | 7881.78 | 0.25 | 0.08 | 3.18 | 0.00 | 0.03 |
| ENSG00000163577.8 | EIF5A2 | 111.35 | -0.50 | 0.13 | -3.79 | 0.00 | 0.01 |
| ENSG00000109911.19 | ELP4 | 670.67 | -0.44 | 0.13 | -3.33 | 0.00 | 0.02 |
| ENSG00000149499.12 | EML3 | 1394.80 | 0.36 | 0.10 | 3.59 | 0.00 | 0.01 |
| ENSG00000254469.9 | ENSG0000025 | 233.19 | -0.57 | 0.18 | -3.18 | 0.00 | 0.03 |
| ENSG00000257767.3 | ENSG0000025 | 95.93 | 0.75 | 0.25 | 2.95 | 0.00 | 0.05 |
| ENSG00000260861.6 | ENSG0000026 | 66.61 | 0.97 | 0.31 | 3.08 | 0.00 | 0.04 |
| ENSG00000263620.1 | ENSG0000026 | 94.75 | 0.62 | 0.15 | 4.18 | 0.00 | 0.00 |
| ENSG00000283782.2 | ENSG0000028 | 158.29 | -1.61 | 0.33 | -4.89 | 0.00 | 0.00 |
| ENSG00000286185.1 | ENSG0000028 | 168.41 | 0.56 | 0.15 | 3.77 | 0.00 | 0.01 |
| ENSG00000286190.2 | ENSG0000028 | 390.20 | 0.36 | 0.11 | 3.21 | 0.00 | 0.03 |
| ENSG00000289027.1 | ENSG0000028 | 256.33 | -0.50 | 0.12 | -4.09 | 0.00 | 0.01 |
| ENSG00000175595.16 | ERCC4 | 471.89 | -0.27 | 0.09 | -3.18 | 0.00 | 0.03 |
| ENSG00000182150.19 | ERCC6L2 | 1843.18 | -0.30 | 0.10 | -3.10 | 0.00 | 0.04 |
| ENSG00000105722.10 | ERF | 1760.11 | 0.33 | 0.11 | 2.99 | 0.00 | 0.04 |
| ENSG00000068912.15 | ERLEC1 | 669.39 | -0.30 | 0.09 | -3.48 | 0.00 | 0.02 |
| ENSG00000136541.15 | ERMN | 163.44 | -0.44 | 0.15 | -2.95 | 0.00 | 0.05 |
| ENSG00000141446.11 | ESCO1 | 724.30 | -0.54 | 0.15 | -3.70 | 0.00 | 0.01 |
| ENSG00000089048.15 | ESF1 | 91.37 | -0.83 | 0.20 | -4.08 | 0.00 | 0.01 |
| ENSG00000143971.9 | ETAA1 | 172.02 | -0.73 | 0.19 | -3.84 | 0.00 | 0.01 |

| SCC DEG set | | | | | | | |
|---|---|---|---|---|---|---|---|
| Ensembl ID | Gene name | baseMean | Log FC | Log FC SE | Wald statistic value | P-value | Adjusted p-value |
| ENSG00000107371.14 | EXOSC3 | 514.49 | -0.31 | 0.08 | -4.07 | 0.00 | 0.01 |
| ENSG00000125898.13 | FAM110A | 992.88 | 0.26 | 0.09 | 3.06 | 0.00 | 0.04 |
| ENSG00000048828.17 | FAM120A | 6964.30 | 0.26 | 0.08 | 3.04 | 0.00 | 0.04 |
| ENSG00000122591.13 | FAM126A | 1125.54 | -0.35 | 0.09 | -3.74 | 0.00 | 0.01 |
| ENSG00000234545.8 | FAM133B | 462.70 | -0.32 | 0.11 | -3.02 | 0.00 | 0.04 |
| ENSG00000221909.3 | FAM200A | 95.07 | -0.43 | 0.13 | -3.42 | 0.00 | 0.02 |
| ENSG00000165669.14 | FAM204A | 815.22 | -0.24 | 0.08 | -2.92 | 0.00 | 0.05 |
| ENSG00000047346.13 | FAM214A | 1785.74 | -0.22 | 0.07 | -3.10 | 0.00 | 0.04 |
| ENSG00000005238.20 | FAM214B | 3321.73 | 0.53 | 0.17 | 3.17 | 0.00 | 0.03 |
| ENSG00000196937.11 | FAM3C | 235.36 | -0.56 | 0.16 | -3.63 | 0.00 | 0.01 |
| ENSG00000120709.11 | FAM53C | 5173.74 | 0.53 | 0.16 | 3.36 | 0.00 | 0.02 |
| ENSG00000171262.11 | FAM98B | 365.96 | -0.48 | 0.12 | -3.95 | 0.00 | 0.01 |
| ENSG00000187790.11 | FANCM | 201.19 | -0.42 | 0.14 | -2.95 | 0.00 | 0.05 |
| ENSG00000156860.16 | FBRS | 5600.01 | 0.38 | 0.11 | 3.37 | 0.00 | 0.02 |
| ENSG00000155034.20 | FBXL18 | 697.35 | 0.25 | 0.08 | 3.05 | 0.00 | 0.04 |
| ENSG00000167196.14 | FBXO22 | 579.25 | -0.51 | 0.11 | -4.48 | 0.00 | 0.00 |
| ENSG00000104870.13 | FCGRT | 6062.41 | 0.50 | 0.12 | 4.26 | 0.00 | 0.00 |
| ENSG00000143297.19 | FCRL5 | 515.52 | -1.29 | 0.43 | -3.01 | 0.00 | 0.04 |
| ENSG00000169018.6 | FEM1B | 1631.16 | -0.27 | 0.09 | -3.12 | 0.00 | 0.04 |
| ENSG00000174721.10 | FGFBP3 | 47.93 | -0.58 | 0.20 | -2.99 | 0.00 | 0.05 |
| ENSG00000183386.10 | FHL3 | 1239.01 | 0.94 | 0.20 | 4.62 | 0.00 | 0.00 |
| ENSG00000135723.14 | FHOD1 | 1201.75 | 0.27 | 0.08 | 3.24 | 0.00 | 0.03 |
| ENSG00000100442.11 | FKBP3 | 164.03 | -0.38 | 0.12 | -3.12 | 0.00 | 0.04 |
| ENSG00000177731.16 | FLII | 5582.63 | 0.36 | 0.10 | 3.42 | 0.00 | 0.02 |
| ENSG00000065970.9 | FOXJ2 | 2032.22 | 0.34 | 0.11 | 3.14 | 0.00 | 0.04 |
| ENSG00000259030.8 | FPGT-TNNI3K | 23.15 | -1.00 | 0.29 | -3.43 | 0.00 | 0.02 |
| ENSG00000148690.12 | FRA10AC1 | 409.09 | -0.49 | 0.17 | -2.98 | 0.00 | 0.05 |
| ENSG00000126391.14 | FRMD8 | 1477.95 | 0.36 | 0.10 | 3.79 | 0.00 | 0.01 |
| ENSG00000075618.18 | FSCN1 | 128.53 | 0.52 | 0.18 | 2.93 | 0.00 | 0.05 |
| ENSG00000162613.17 | FUBP1 | 2450.13 | -0.28 | 0.08 | -3.42 | 0.00 | 0.02 |
| ENSG00000178950.18 | GAK | 4036.01 | 0.30 | 0.06 | 4.94 | 0.00 | 0.00 |
| ENSG00000156958.15 | GALK2 | 466.64 | -0.22 | 0.07 | -3.03 | 0.00 | 0.04 |
| ENSG00000102145.15 | GATA1 | 841.24 | 0.91 | 0.30 | 3.02 | 0.00 | 0.04 |
| ENSG00000135968.22 | GCC2 | 1943.96 | -0.50 | 0.11 | -4.38 | 0.00 | 0.00 |
| ENSG00000140905.11 | GCSH | 22.78 | -0.71 | 0.23 | -3.13 | 0.00 | 0.04 |
| ENSG00000158555.15 | GDPD5 | 820.25 | 0.48 | 0.13 | 3.71 | 0.00 | 0.01 |
| ENSG00000178295.15 | GEN1 | 327.02 | -0.51 | 0.17 | -3.07 | 0.00 | 0.04 |
| ENSG00000125447.18 | GGA3 | 2315.52 | 0.25 | 0.08 | 2.98 | 0.00 | 0.05 |
| ENSG00000146830.11 | GIGYF1 | 2817.28 | 0.25 | 0.08 | 3.16 | 0.00 | 0.03 |
| ENSG00000088256.9 | GNA11 | 106.42 | 0.49 | 0.15 | 3.28 | 0.00 | 0.03 |
| ENSG00000114353.17 | GNAI2 | 20197.67 | 0.34 | 0.11 | 2.98 | 0.00 | 0.05 |
| ENSG00000141404.17 | GNAL | 29.72 | -1.47 | 0.37 | -3.97 | 0.00 | 0.01 |
| ENSG00000130119.17 | GNL3L | 3288.11 | 0.35 | 0.10 | 3.37 | 0.00 | 0.02 |
| ENSG00000100522.10 | GNPNAT1 | 145.21 | -0.34 | 0.12 | -2.98 | 0.00 | 0.05 |
| ENSG00000144674.17 | GOLGA4 | 1538.61 | -0.40 | 0.11 | -3.56 | 0.00 | 0.02 |
| ENSG00000047932.14 | GOPC | 684.96 | -0.32 | 0.11 | -2.93 | 0.00 | 0.05 |
| ENSG00000120370.14 | GORAB | 305.47 | -0.38 | 0.12 | -3.11 | 0.00 | 0.04 |
| ENSG00000092978.11 | GPATCH2 | 239.89 | -0.39 | 0.12 | -3.12 | 0.00 | 0.04 |
| ENSG00000188394.8 | GPR21 | 83.63 | 0.35 | 0.11 | 3.16 | 0.00 | 0.03 |
| ENSG00000188092.15 | GPR89B | 327.47 | -0.38 | 0.11 | -3.48 | 0.00 | 0.02 |
| ENSG00000178719.17 | GRINA | 10020.47 | 0.75 | 0.25 | 3.04 | 0.00 | 0.04 |
| ENSG00000173020.11 | GRK2 | 13627.11 | 0.33 | 0.10 | 3.21 | 0.00 | 0.03 |
| ENSG00000148180.21 | GSN | 4547.81 | 0.64 | 0.19 | 3.37 | 0.00 | 0.02 |
| ENSG00000119041.11 | GTF3C3 | 370.78 | -0.31 | 0.09 | -3.35 | 0.00 | 0.02 |

| SCC DEG set | | | | | | | |
|---|---|---|---|---|---|---|---|
| Ensembl ID | Gene name | baseMean | Log FC | Log FC SE | Wald statistic value | P-value | Adjusted p-value |
| ENSG00000105793.16 | GTPBP10 | 473.92 | -0.36 | 0.08 | -4.55 | 0.00 | 0.00 |
| ENSG00000163607.18 | GTPBP8 | 143.60 | -0.53 | 0.13 | -3.99 | 0.00 | 0.01 |
| ENSG00000138867.17 | GUCD1 | 8873.41 | 0.73 | 0.25 | 2.97 | 0.00 | 0.05 |
| ENSG00000147874.11 | HAUS6 | 518.83 | -0.26 | 0.08 | -3.09 | 0.00 | 0.04 |
| ENSG00000112339.15 | HBS1L | 899.64 | -0.33 | 0.10 | -3.40 | 0.00 | 0.02 |
| ENSG00000100429.18 | HDAC10 | 758.30 | 0.24 | 0.08 | 2.92 | 0.00 | 0.05 |
| ENSG00000115677.18 | HDLBP | 5083.85 | 0.33 | 0.11 | 2.93 | 0.00 | 0.05 |
| ENSG00000165259.14 | HDX | 63.58 | -0.61 | 0.19 | -3.16 | 0.00 | 0.03 |
| ENSG00000173064.14 | HECTD4 | 4605.79 | 0.39 | 0.12 | 3.15 | 0.00 | 0.03 |
| ENSG00000163312.11 | HELQ | 286.13 | -0.29 | 0.08 | -3.68 | 0.00 | 0.01 |
| ENSG00000136929.13 | HEMGN | 1813.39 | 1.02 | 0.34 | 2.99 | 0.00 | 0.04 |
| ENSG00000185359.14 | HGS | 2277.28 | 0.36 | 0.10 | 3.68 | 0.00 | 0.01 |
| ENSG00000127946.17 | HIP1 | 3230.65 | 0.89 | 0.25 | 3.52 | 0.00 | 0.02 |
| ENSG00000237541.4 | HLA-DQA2 | 116.62 | -2.86 | 0.81 | -3.54 | 0.00 | 0.02 |
| ENSG00000152795.18 | HNRNPDL | 2645.47 | -0.39 | 0.11 | -3.50 | 0.00 | 0.02 |
| ENSG00000204389.10 | HSPA1A | 7258.79 | 0.72 | 0.22 | 3.19 | 0.00 | 0.03 |
| ENSG00000204388.7 | HSPA1B | 1288.65 | 1.12 | 0.34 | 3.33 | 0.00 | 0.02 |
| ENSG00000081870.12 | HSPB11 | 146.18 | -0.32 | 0.10 | -3.23 | 0.00 | 0.03 |
| ENSG00000163596.17 | ICA1L | 47.36 | -0.54 | 0.17 | -3.11 | 0.00 | 0.04 |
| ENSG00000076662.10 | ICAM3 | 13102.84 | 0.50 | 0.14 | 3.67 | 0.00 | 0.01 |
| ENSG00000105371.10 | ICAM4 | 78.23 | 0.72 | 0.22 | 3.30 | 0.00 | 0.03 |
| ENSG00000096872.17 | IFT74 | 91.18 | -0.61 | 0.16 | -3.72 | 0.00 | 0.01 |
| ENSG00000197081.16 | IGF2R | 40531.17 | 0.60 | 0.19 | 3.15 | 0.00 | 0.03 |
| ENSG00000240864.3 | IGKV1-16 | 53.49 | -1.09 | 0.31 | -3.55 | 0.00 | 0.02 |
| ENSG00000239855.1 | IGKV1-6 | 31.48 | -1.26 | 0.33 | -3.80 | 0.00 | 0.01 |
| ENSG00000241351.3 | IGKV3-11 | 263.60 | -1.31 | 0.35 | -3.78 | 0.00 | 0.01 |
| ENSG00000211651.3 | IGLV1-44 | 156.60 | -1.20 | 0.33 | -3.68 | 0.00 | 0.01 |
| ENSG00000211669.3 | IGLV3-10 | 24.90 | -1.38 | 0.43 | -3.24 | 0.00 | 0.03 |
| ENSG00000211663.2 | IGLV3-19 | 70.92 | -1.06 | 0.36 | -2.97 | 0.00 | 0.05 |
| ENSG00000095574.12 | IKZF5 | 803.44 | -0.23 | 0.08 | -2.93 | 0.00 | 0.05 |
| ENSG00000196083.10 | IL1RAP | 2598.02 | 0.60 | 0.20 | 2.92 | 0.00 | 0.05 |
| ENSG00000198825.15 | INPP5F | 253.45 | -0.36 | 0.11 | -3.17 | 0.00 | 0.03 |
| ENSG00000164880.16 | INTS1 | 2197.45 | 0.27 | 0.09 | 3.03 | 0.00 | 0.04 |
| ENSG00000064102.15 | INTS13 | 176.43 | -0.30 | 0.09 | -3.47 | 0.00 | 0.02 |
| ENSG00000143624.14 | INTS3 | 2653.38 | 0.37 | 0.11 | 3.46 | 0.00 | 0.02 |
| ENSG00000086200.17 | IPO11 | 303.24 | -0.39 | 0.11 | -3.64 | 0.00 | 0.01 |
| ENSG00000173226.17 | IQCB1 | 237.00 | -0.43 | 0.14 | -2.94 | 0.00 | 0.05 |
| ENSG00000114473.14 | IQCG | 172.59 | -0.45 | 0.14 | -3.20 | 0.00 | 0.03 |
| ENSG00000144711.16 | IQSEC1 | 15680.92 | 0.42 | 0.13 | 3.08 | 0.00 | 0.04 |
| ENSG00000161638.11 | ITGA5 | 5577.75 | 0.39 | 0.11 | 3.53 | 0.00 | 0.02 |
| ENSG00000005844.19 | ITGAL | 11781.25 | 0.29 | 0.10 | 3.06 | 0.00 | 0.04 |
| ENSG00000140678.17 | ITGAX | 25954.15 | 0.55 | 0.17 | 3.35 | 0.00 | 0.02 |
| ENSG00000160593.19 | JAML | 28958.93 | 0.44 | 0.14 | 3.07 | 0.00 | 0.04 |
| ENSG00000092051.18 | JPH4 | 34.35 | 0.63 | 0.17 | 3.64 | 0.00 | 0.01 |
| ENSG00000103510.20 | KAT8 | 1216.86 | 0.19 | 0.05 | 3.73 | 0.00 | 0.01 |
| ENSG00000099337.5 | KCNK6 | 961.42 | 0.24 | 0.08 | 3.11 | 0.00 | 0.04 |
| ENSG00000127663.15 | KDM4B | 7206.46 | 0.38 | 0.12 | 3.20 | 0.00 | 0.03 |
| ENSG00000132510.11 | KDM6B | 19928.05 | 0.56 | 0.17 | 3.18 | 0.00 | 0.03 |
| ENSG00000007202.15 | KIAA0100 | 4107.40 | 0.24 | 0.07 | 3.35 | 0.00 | 0.02 |
| ENSG00000075945.13 | KIFAP3 | 386.44 | -0.35 | 0.10 | -3.62 | 0.00 | 0.01 |
| ENSG00000169926.11 | KLF13 | 10342.50 | 0.24 | 0.08 | 3.17 | 0.00 | 0.03 |
| ENSG00000272333.8 | KMT2B | 4466.83 | 0.41 | 0.10 | 4.27 | 0.00 | 0.00 |
| ENSG00000167548.18 | KMT2D | 21680.88 | 0.41 | 0.11 | 3.75 | 0.00 | 0.01 |
| ENSG00000196911.11 | KPNA5 | 341.03 | -0.61 | 0.14 | -4.26 | 0.00 | 0.00 |

| SCC DEG set | | | | | | | |
|---|---|---|---|---|---|---|---|
| Ensembl ID | Gene name | baseMean | Log FC | Log FC SE | Wald statistic value | P-value | Adjusted p-value |
| ENSG00000001631.17 | KRIT1 | 1826.59 | -0.27 | 0.07 | -3.94 | 0.00 | 0.01 |
| ENSG00000111615.14 | KRR1 | 532.43 | -0.51 | 0.12 | -4.30 | 0.00 | 0.00 |
| ENSG00000167768.4 | KRT1 | 1398.25 | 1.63 | 0.51 | 3.20 | 0.00 | 0.03 |
| ENSG00000108244.17 | KRT23 | 1523.27 | 0.80 | 0.25 | 3.23 | 0.00 | 0.03 |
| ENSG00000189182.10 | KRT77 | 20.56 | 9.27 | 2.62 | 3.54 | 0.00 | 0.02 |
| ENSG00000126777.18 | KTN1 | 1286.85 | -0.34 | 0.11 | -3.23 | 0.00 | 0.03 |
| ENSG00000213398.8 | LCAT | 93.60 | 0.43 | 0.12 | 3.52 | 0.00 | 0.02 |
| ENSG00000166816.15 | LDHD | 24.05 | 0.91 | 0.31 | 2.93 | 0.00 | 0.05 |
| ENSG00000133317.16 | LGALS12 | 236.61 | 0.53 | 0.16 | 3.37 | 0.00 | 0.02 |
| ENSG00000131042.15 | LILRB2 | 7510.47 | 0.48 | 0.12 | 4.02 | 0.00 | 0.01 |
| ENSG00000148943.12 | LIN7C | 979.11 | -0.30 | 0.10 | -2.94 | 0.00 | 0.05 |
| ENSG00000077454.17 | LRCH4 | 5554.30 | 0.42 | 0.11 | 3.94 | 0.00 | 0.01 |
| ENSG00000123384.14 | LRP1 | 8897.14 | 0.84 | 0.21 | 4.01 | 0.00 | 0.01 |
| ENSG00000130881.14 | LRP3 | 162.91 | 0.53 | 0.17 | 3.18 | 0.00 | 0.03 |
| ENSG00000161036.13 | LRWD1 | 708.56 | 0.58 | 0.17 | 3.37 | 0.00 | 0.02 |
| ENSG00000128534.8 | LSM8 | 1459.21 | -0.45 | 0.13 | -3.47 | 0.00 | 0.02 |
| ENSG00000130592.17 | LSP1 | 27706.50 | 0.45 | 0.15 | 2.97 | 0.00 | 0.05 |
| ENSG00000204482.11 | LST1 | 3767.41 | 0.39 | 0.12 | 3.30 | 0.00 | 0.03 |
| ENSG00000119681.12 | LTBP2 | 229.14 | 0.76 | 0.24 | 3.12 | 0.00 | 0.04 |
| ENSG00000108848.16 | LUC7L3 | 2126.08 | -0.37 | 0.08 | -4.36 | 0.00 | 0.00 |
| ENSG00000061337.15 | LZTS1 | 22.64 | 1.20 | 0.34 | 3.57 | 0.00 | 0.02 |
| ENSG00000116670.15 | MAD2L2 | 197.41 | 0.38 | 0.12 | 3.10 | 0.00 | 0.04 |
| ENSG00000111196.10 | MAGOHB | 145.79 | -0.46 | 0.16 | -2.95 | 0.00 | 0.05 |
| ENSG00000196782.13 | MAML3 | 2002.69 | 0.41 | 0.13 | 3.25 | 0.00 | 0.03 |
| ENSG00000104774.13 | MAN2B1 | 4661.52 | 0.28 | 0.08 | 3.38 | 0.00 | 0.02 |
| ENSG00000130479.11 | MAP1S | 1405.33 | 0.31 | 0.10 | 3.07 | 0.00 | 0.04 |
| ENSG00000173327.8 | MAP3K11 | 7200.61 | 0.37 | 0.13 | 2.92 | 0.00 | 0.05 |
| ENSG00000142733.17 | MAP3K6 | 169.97 | 0.48 | 0.13 | 3.61 | 0.00 | 0.01 |
| ENSG00000011566.15 | MAP4K3 | 199.15 | -0.39 | 0.11 | -3.55 | 0.00 | 0.02 |
| ENSG00000116871.16 | MAP7D1 | 5434.74 | 0.37 | 0.11 | 3.34 | 0.00 | 0.02 |
| ENSG00000069956.14 | MAPK6 | 370.22 | -0.35 | 0.12 | -3.02 | 0.00 | 0.04 |
| ENSG00000107643.17 | MAPK8 | 582.09 | -0.32 | 0.09 | -3.60 | 0.00 | 0.01 |
| ENSG00000099308.11 | MAST3 | 7420.75 | 0.51 | 0.14 | 3.78 | 0.00 | 0.01 |
| ENSG00000015479.20 | MATR3 | 3512.90 | -0.32 | 0.10 | -3.06 | 0.00 | 0.04 |
| ENSG00000088888.18 | MAVS | 5507.04 | 0.19 | 0.06 | 3.21 | 0.00 | 0.03 |
| ENSG00000166987.15 | MBD6 | 5660.78 | 0.41 | 0.13 | 3.07 | 0.00 | 0.04 |
| ENSG00000125505.17 | MBOAT7 | 15711.69 | 0.48 | 0.15 | 3.13 | 0.00 | 0.04 |
| ENSG00000197971.16 | MBP | 11590.81 | 0.30 | 0.10 | 2.98 | 0.00 | 0.05 |
| ENSG00000012174.12 | MBTPS2 | 221.01 | -0.37 | 0.11 | -3.54 | 0.00 | 0.02 |
| ENSG00000232119.8 | MCTS1 | 703.33 | -0.26 | 0.07 | -3.55 | 0.00 | 0.02 |
| ENSG00000111554.15 | MDM1 | 348.36 | -0.32 | 0.10 | -3.22 | 0.00 | 0.03 |
| ENSG00000184634.17 | MED12 | 3335.64 | 0.37 | 0.12 | 3.23 | 0.00 | 0.03 |
| ENSG00000175221.16 | MED16 | 1734.69 | 0.45 | 0.15 | 2.95 | 0.00 | 0.05 |
| ENSG00000152944.9 | MED21 | 519.93 | -0.31 | 0.10 | -3.19 | 0.00 | 0.03 |
| ENSG00000104973.19 | MED25 | 3898.57 | 0.60 | 0.17 | 3.55 | 0.00 | 0.02 |
| ENSG00000116604.19 | MEF2D | 3391.28 | 0.29 | 0.08 | 3.48 | 0.00 | 0.02 |
| ENSG00000145388.15 | METTL14 | 527.50 | -0.26 | 0.08 | -3.39 | 0.00 | 0.02 |
| ENSG00000109736.15 | MFSD10 | 612.53 | 0.31 | 0.10 | 3.00 | 0.00 | 0.04 |
| ENSG00000164073.11 | MFSD8 | 575.41 | -0.23 | 0.07 | -3.06 | 0.00 | 0.04 |
| ENSG00000197530.13 | MIB2 | 629.52 | 0.28 | 0.10 | 2.98 | 0.00 | 0.05 |
| ENSG00000164877.19 | MICALL2 | 76.95 | 0.69 | 0.20 | 3.46 | 0.00 | 0.02 |
| ENSG00000165487.14 | MICU2 | 628.96 | -0.30 | 0.10 | -3.09 | 0.00 | 0.04 |
| ENSG00000155970.12 | MICU3 | 26.66 | -1.16 | 0.33 | -3.47 | 0.00 | 0.02 |
| ENSG00000165175.17 | MID1IP1 | 1800.85 | 0.37 | 0.12 | 3.08 | 0.00 | 0.04 |

| SCC DEG set | | | | | | | |
|---|---|---|---|---|---|---|---|
| Ensembl ID | Gene name | baseMean | Log FC | Log FC SE | Wald statistic value | P-value | Adjusted p-value |
| ENSG00000141503.17 | MINK1 | 3007.63 | 0.28 | 0.09 | 3.16 | 0.00 | 0.03 |
| ENSG00000175727.14 | MLXIP | 4343.61 | 0.36 | 0.10 | 3.71 | 0.00 | 0.01 |
| ENSG00000198598.7 | MMP17 | 206.46 | 0.42 | 0.13 | 3.17 | 0.00 | 0.03 |
| ENSG00000020426.11 | MNAT1 | 94.01 | -0.60 | 0.16 | -3.78 | 0.00 | 0.01 |
| ENSG00000182208.15 | MOB2 | 825.72 | 0.23 | 0.08 | 2.92 | 0.00 | 0.05 |
| ENSG00000185716.12 | MOSMO | 307.24 | -0.29 | 0.10 | -3.00 | 0.00 | 0.04 |
| ENSG00000124383.9 | MPHOSPH10 | 240.60 | -0.46 | 0.15 | -2.97 | 0.00 | 0.05 |
| ENSG00000051825.15 | MPHOSPH9 | 678.12 | -0.44 | 0.12 | -3.61 | 0.00 | 0.01 |
| ENSG00000117400.18 | MPL | 230.80 | 0.99 | 0.31 | 3.17 | 0.00 | 0.03 |
| ENSG00000149573.9 | MPZL2 | 283.57 | 0.63 | 0.21 | 3.07 | 0.00 | 0.04 |
| ENSG00000011028.14 | MRC2 | 521.12 | 1.72 | 0.33 | 5.23 | 0.00 | 0.00 |
| ENSG00000020922.13 | MRE11 | 760.43 | -0.26 | 0.08 | -3.29 | 0.00 | 0.03 |
| ENSG00000204839.9 | MROH6 | 290.28 | 0.80 | 0.26 | 3.07 | 0.00 | 0.04 |
| ENSG00000136897.8 | MRPL50 | 168.58 | -0.48 | 0.16 | -2.97 | 0.00 | 0.05 |
| ENSG00000120333.4 | MRPS14 | 222.27 | -0.44 | 0.12 | -3.80 | 0.00 | 0.01 |
| ENSG00000175110.14 | MRPS22 | 391.46 | -0.28 | 0.09 | -3.04 | 0.00 | 0.04 |
| ENSG00000196588.21 | MRTFA | 4148.43 | 0.35 | 0.09 | 3.78 | 0.00 | 0.01 |
| ENSG00000095002.15 | MSH2 | 260.94 | -0.48 | 0.16 | -3.06 | 0.00 | 0.04 |
| ENSG00000147065.17 | MSN | 33057.69 | 0.33 | 0.11 | 3.11 | 0.00 | 0.04 |
| ENSG00000137409.20 | MTCH1 | 1784.93 | 0.25 | 0.08 | 3.30 | 0.00 | 0.03 |
| ENSG00000127989.14 | MTERF1 | 312.54 | -0.54 | 0.11 | -4.77 | 0.00 | 0.00 |
| ENSG00000156469.9 | MTERF3 | 62.21 | -0.44 | 0.14 | -3.04 | 0.00 | 0.04 |
| ENSG00000143033.18 | MTF2 | 631.03 | -0.33 | 0.10 | -3.44 | 0.00 | 0.02 |
| ENSG00000177000.13 | MTHFR | 2104.25 | 0.22 | 0.07 | 3.10 | 0.00 | 0.04 |
| ENSG00000085760.15 | MTIF2 | 245.43 | -0.29 | 0.09 | -3.03 | 0.00 | 0.04 |
| ENSG00000087053.20 | MTMR2 | 339.35 | -0.35 | 0.10 | -3.39 | 0.00 | 0.02 |
| ENSG00000039123.16 | MTREX | 853.87 | -0.43 | 0.11 | -3.78 | 0.00 | 0.01 |
| ENSG00000128654.14 | MTX2 | 44.25 | -0.54 | 0.18 | -3.00 | 0.00 | 0.04 |
| ENSG00000116990.11 | MYCL | 712.96 | 0.37 | 0.11 | 3.45 | 0.00 | 0.02 |
| ENSG00000100345.23 | MYH9 | 45563.92 | 0.49 | 0.15 | 3.26 | 0.00 | 0.03 |
| ENSG00000197879.17 | MYO1C | 378.82 | 0.30 | 0.10 | 3.01 | 0.00 | 0.04 |
| ENSG00000142347.19 | MYO1F | 34736.88 | 0.49 | 0.14 | 3.42 | 0.00 | 0.02 |
| ENSG00000099331.14 | MYO9B | 12371.37 | 0.37 | 0.11 | 3.26 | 0.00 | 0.03 |
| ENSG00000162601.11 | MYSM1 | 2005.47 | -0.28 | 0.08 | -3.54 | 0.00 | 0.02 |
| ENSG00000078177.14 | N4BP2 | 790.35 | -0.48 | 0.16 | -2.93 | 0.00 | 0.05 |
| ENSG00000164134.13 | NAA15 | 809.87 | -0.41 | 0.13 | -3.22 | 0.00 | 0.03 |
| ENSG00000111300.10 | NAA25 | 574.20 | -0.34 | 0.12 | -2.93 | 0.00 | 0.05 |
| ENSG00000139977.14 | NAA30 | 541.98 | -0.33 | 0.11 | -3.05 | 0.00 | 0.04 |
| ENSG00000152620.13 | NADK2 | 178.22 | -0.36 | 0.10 | -3.53 | 0.00 | 0.02 |
| ENSG00000198951.12 | NAGA | 1687.95 | 0.28 | 0.08 | 3.28 | 0.00 | 0.03 |
| ENSG00000187109.15 | NAP1L1 | 3191.85 | -0.32 | 0.09 | -3.68 | 0.00 | 0.01 |
| ENSG00000134369.16 | NAV1 | 335.32 | 0.46 | 0.15 | 3.08 | 0.00 | 0.04 |
| ENSG00000160796.18 | NBEAL2 | 21652.10 | 0.59 | 0.16 | 3.74 | 0.00 | 0.01 |
| ENSG00000100365.16 | NCF4 | 6674.69 | 0.54 | 0.18 | 3.03 | 0.00 | 0.04 |
| ENSG00000167566.17 | NCKAP5L | 1472.25 | 0.32 | 0.09 | 3.52 | 0.00 | 0.02 |
| ENSG00000196498.13 | NCOR2 | 4449.08 | 0.25 | 0.09 | 2.94 | 0.00 | 0.05 |
| ENSG00000110400.11 | NECTIN1 | 522.20 | 0.59 | 0.15 | 4.03 | 0.00 | 0.01 |
| ENSG00000165525.18 | NEMF | 1135.49 | -0.36 | 0.09 | -3.92 | 0.00 | 0.01 |
| ENSG00000196865.4 | NHLRC2 | 1319.23 | -0.26 | 0.08 | -3.22 | 0.00 | 0.03 |
| ENSG00000136830.12 | NIBAN2 | 1051.11 | 0.48 | 0.15 | 3.19 | 0.00 | 0.03 |
| ENSG00000171840.13 | NINJ2 | 542.42 | 0.79 | 0.22 | 3.50 | 0.00 | 0.02 |
| ENSG00000114857.19 | NKTR | 6137.74 | -0.24 | 0.08 | -2.96 | 0.00 | 0.05 |
| ENSG00000091592.16 | NLRP1 | 9660.05 | 0.32 | 0.10 | 3.04 | 0.00 | 0.04 |
| ENSG00000022556.16 | NLRP2 | 67.40 | -1.10 | 0.37 | -2.95 | 0.00 | 0.05 |

| SCC DEG set | | | | | | | |
|---|---|---|---|---|---|---|---|
| Ensembl ID | Gene name | baseMean | Log FC | Log FC SE | Wald statistic value | P-value | Adjusted p-value |
| ENSG00000162711.18 | NLRP3 | 1057.77 | 0.45 | 0.12 | 3.70 | 0.00 | 0.01 |
| ENSG00000169251.13 | NMD3 | 246.34 | -0.48 | 0.16 | -2.98 | 0.00 | 0.05 |
| ENSG00000143156.14 | NME7 | 85.86 | -0.50 | 0.15 | -3.46 | 0.00 | 0.02 |
| ENSG00000173145.11 | NOC3L | 196.26 | -0.55 | 0.16 | -3.53 | 0.00 | 0.02 |
| ENSG00000198000.12 | NOL8 | 693.30 | -0.37 | 0.09 | -4.17 | 0.00 | 0.00 |
| ENSG00000148400.13 | NOTCH1 | 15717.40 | 0.60 | 0.16 | 3.80 | 0.00 | 0.01 |
| ENSG00000134250.21 | NOTCH2 | 19383.98 | 0.49 | 0.14 | 3.41 | 0.00 | 0.02 |
| ENSG00000120798.17 | NR2C1 | 264.96 | -0.45 | 0.14 | -3.16 | 0.00 | 0.03 |
| ENSG00000177463.16 | NR2C2 | 3496.35 | 0.19 | 0.07 | 2.93 | 0.00 | 0.05 |
| ENSG00000148200.17 | NR6A1 | 519.93 | 0.53 | 0.16 | 3.26 | 0.00 | 0.03 |
| ENSG00000149761.9 | NUDT22 | 471.75 | 0.29 | 0.08 | 3.72 | 0.00 | 0.01 |
| ENSG00000170917.14 | NUDT6 | 52.27 | -0.75 | 0.20 | -3.78 | 0.00 | 0.01 |
| ENSG00000093000.19 | NUP50 | 5252.11 | 0.27 | 0.09 | 3.02 | 0.00 | 0.04 |
| ENSG00000152193.8 | OBI1 | 259.24 | -0.35 | 0.10 | -3.44 | 0.00 | 0.02 |
| ENSG00000122417.15 | ODF2L | 615.95 | -0.55 | 0.12 | -4.61 | 0.00 | 0.00 |
| ENSG00000105953.16 | OGDH | 2719.49 | 0.31 | 0.09 | 3.59 | 0.00 | 0.01 |
| ENSG00000160991.17 | ORAI2 | 6578.38 | 0.44 | 0.14 | 3.08 | 0.00 | 0.04 |
| ENSG00000155100.11 | OTUD6B | 225.19 | -0.53 | 0.13 | -3.96 | 0.00 | 0.01 |
| ENSG00000175115.13 | PACS1 | 6755.89 | 0.37 | 0.11 | 3.31 | 0.00 | 0.03 |
| ENSG00000149269.10 | PAK1 | 4848.98 | 0.46 | 0.15 | 3.12 | 0.00 | 0.04 |
| ENSG00000198682.13 | PAPSS2 | 118.36 | 0.49 | 0.15 | 3.28 | 0.00 | 0.03 |
| ENSG00000163291.14 | PAQR3 | 183.40 | -0.38 | 0.12 | -3.09 | 0.00 | 0.04 |
| ENSG00000140694.17 | PARN | 890.84 | -0.24 | 0.05 | -4.39 | 0.00 | 0.00 |
| ENSG00000173200.13 | PARP15 | 1199.65 | -0.56 | 0.19 | -3.00 | 0.00 | 0.04 |
| ENSG00000138964.17 | PARVG | 6723.58 | 0.26 | 0.09 | 3.08 | 0.00 | 0.04 |
| ENSG00000159086.15 | PAXBP1 | 685.33 | -0.39 | 0.10 | -3.93 | 0.00 | 0.01 |
| ENSG00000184226.15 | PCDH9 | 464.07 | -1.32 | 0.33 | -4.05 | 0.00 | 0.01 |
| ENSG00000253731.3 | PCDHGA6 | 38.21 | -0.85 | 0.28 | -2.99 | 0.00 | 0.04 |
| ENSG00000078674.19 | PCM1 | 3170.61 | -0.32 | 0.07 | -4.47 | 0.00 | 0.00 |
| ENSG00000197136.5 | PCNX3 | 2282.19 | 0.25 | 0.08 | 3.09 | 0.00 | 0.04 |
| ENSG00000126773.13 | PCNX4 | 1656.86 | -0.32 | 0.09 | -3.62 | 0.00 | 0.01 |
| ENSG00000071994.11 | PDCD2 | 652.72 | -0.26 | 0.08 | -3.48 | 0.00 | 0.02 |
| ENSG00000175087.10 | PDIK1L | 207.01 | -0.40 | 0.12 | -3.26 | 0.00 | 0.03 |
| ENSG00000160209.19 | PDXK | 2649.10 | 0.34 | 0.11 | 3.07 | 0.00 | 0.04 |
| ENSG00000162366.8 | PDZK1IP1 | 1696.92 | 1.14 | 0.30 | 3.76 | 0.00 | 0.01 |
| ENSG00000127980.16 | PEX1 | 407.29 | -0.26 | 0.09 | -2.97 | 0.00 | 0.05 |
| ENSG00000108518.8 | PFN1 | 16750.72 | 0.25 | 0.08 | 3.03 | 0.00 | 0.04 |
| ENSG00000185220.12 | PGBD2 | 262.07 | -0.25 | 0.06 | -3.95 | 0.00 | 0.01 |
| ENSG00000013375.16 | PGM3 | 225.25 | -0.39 | 0.12 | -3.28 | 0.00 | 0.03 |
| ENSG00000106443.17 | PHF14 | 695.60 | -0.53 | 0.12 | -4.42 | 0.00 | 0.00 |
| ENSG00000070047.13 | PHRF1 | 1707.70 | 0.20 | 0.07 | 3.02 | 0.00 | 0.04 |
| ENSG00000006576.16 | PHTF2 | 899.76 | -0.42 | 0.11 | -3.72 | 0.00 | 0.01 |
| ENSG00000078043.16 | PIAS2 | 988.95 | -0.42 | 0.10 | -4.09 | 0.00 | 0.01 |
| ENSG00000083535.16 | PIBF1 | 322.39 | -0.57 | 0.15 | -3.80 | 0.00 | 0.01 |
| ENSG00000165195.16 | PIGA | 240.43 | -0.30 | 0.09 | -3.37 | 0.00 | 0.02 |
| ENSG00000119227.8 | PIGZ | 28.69 | 0.56 | 0.19 | 2.98 | 0.00 | 0.05 |
| ENSG00000011405.14 | PIK3C2A | 1336.31 | -0.23 | 0.08 | -3.03 | 0.00 | 0.04 |
| ENSG00000171608.17 | PIK3CD | 13543.82 | 0.45 | 0.15 | 3.02 | 0.00 | 0.04 |
| ENSG00000141506.15 | PIK3R5 | 7485.51 | 0.36 | 0.10 | 3.62 | 0.00 | 0.01 |
| ENSG00000186111.11 | PIP5K1C | 1491.03 | 0.27 | 0.07 | 3.98 | 0.00 | 0.01 |
| ENSG00000174238.16 | PITPNA | 2471.56 | 0.32 | 0.10 | 3.39 | 0.00 | 0.02 |
| ENSG00000110697.13 | PITPNM1 | 2577.64 | 0.31 | 0.09 | 3.53 | 0.00 | 0.02 |
| ENSG00000137055.15 | PLAA | 577.83 | -0.22 | 0.07 | -3.17 | 0.00 | 0.03 |
| ENSG00000181690.8 | PLAG1 | 157.32 | -0.49 | 0.17 | -2.92 | 0.00 | 0.05 |

| SCC DEG set | | | | | | | |
|---|---|---|---|---|---|---|---|
| Ensembl ID | Gene name | baseMean | Log FC | Log FC SE | Wald statistic value | P-value | Adjusted p-value |
| ENSG00000129219.14 | PLD2 | 353.52 | 0.30 | 0.09 | 3.28 | 0.00 | 0.03 |
| ENSG00000023902.14 | PLEKHO1 | 3722.66 | 0.25 | 0.08 | 3.02 | 0.00 | 0.04 |
| ENSG00000106397.12 | PLOD3 | 402.79 | 0.31 | 0.09 | 3.58 | 0.00 | 0.02 |
| ENSG00000171566.12 | PLRG1 | 313.16 | -0.35 | 0.11 | -3.13 | 0.00 | 0.04 |
| ENSG00000196576.16 | PLXNB2 | 3868.79 | 0.42 | 0.12 | 3.65 | 0.00 | 0.01 |
| ENSG00000004399.13 | PLXND1 | 2206.46 | 0.53 | 0.11 | 4.89 | 0.00 | 0.00 |
| ENSG00000105819.14 | PMPCB | 722.04 | -0.28 | 0.07 | -4.04 | 0.00 | 0.01 |
| ENSG00000100941.9 | PNN | 872.38 | -0.30 | 0.10 | -2.92 | 0.00 | 0.05 |
| ENSG00000255529.9 | POLR2M | 549.79 | -0.37 | 0.11 | -3.29 | 0.00 | 0.03 |
| ENSG00000172531.16 | PPP1CA | 3195.39 | 0.25 | 0.08 | 3.13 | 0.00 | 0.04 |
| ENSG00000125503.13 | PPP1R12C | 2957.05 | 0.24 | 0.07 | 3.25 | 0.00 | 0.03 |
| ENSG00000108819.11 | PPP1R9B | 9365.99 | 0.31 | 0.09 | 3.45 | 0.00 | 0.02 |
| ENSG00000105063.19 | PPP6R1 | 7901.53 | 0.33 | 0.11 | 3.00 | 0.00 | 0.04 |
| ENSG00000100239.16 | PPP6R2 | 1574.81 | 0.20 | 0.07 | 2.98 | 0.00 | 0.05 |
| ENSG00000113593.12 | PPWD1 | 499.65 | -0.46 | 0.09 | -5.10 | 0.00 | 0.00 |
| ENSG00000117592.9 | PRDX6 | 4305.53 | 0.83 | 0.22 | 3.80 | 0.00 | 0.01 |
| ENSG00000138078.16 | PREPL | 748.14 | -0.39 | 0.13 | -2.93 | 0.00 | 0.05 |
| ENSG00000124126.14 | PREX1 | 25678.28 | 0.44 | 0.14 | 3.01 | 0.00 | 0.04 |
| ENSG00000164306.11 | PRIMPOL | 126.13 | -0.38 | 0.11 | -3.34 | 0.00 | 0.02 |
| ENSG00000142875.20 | PRKACB | 1587.08 | -0.45 | 0.15 | -3.02 | 0.00 | 0.04 |
| ENSG00000185238.13 | PRMT3 | 322.50 | -0.41 | 0.08 | -5.15 | 0.00 | 0.00 |
| ENSG00000164169.13 | PRMT9 | 171.11 | -0.32 | 0.10 | -3.18 | 0.00 | 0.03 |
| ENSG00000185246.18 | PRPF39 | 356.66 | -0.37 | 0.10 | -3.61 | 0.00 | 0.01 |
| ENSG00000068489.13 | PRR11 | 314.58 | 0.54 | 0.16 | 3.33 | 0.00 | 0.02 |
| ENSG00000204469.13 | PRRC2A | 9368.94 | 0.34 | 0.09 | 3.65 | 0.00 | 0.01 |
| ENSG00000125637.16 | PSD4 | 10148.81 | 0.32 | 0.09 | 3.50 | 0.00 | 0.02 |
| ENSG00000164985.15 | PSIP1 | 1273.34 | -0.36 | 0.07 | -5.02 | 0.00 | 0.00 |
| ENSG00000115233.12 | PSMD14 | 221.16 | -0.45 | 0.12 | -3.84 | 0.00 | 0.01 |
| ENSG00000095261.14 | PSMD5 | 588.71 | -0.25 | 0.07 | -3.37 | 0.00 | 0.02 |
| ENSG00000131467.12 | PSME3 | 2507.66 | 0.31 | 0.09 | 3.58 | 0.00 | 0.02 |
| ENSG00000125818.18 | PSMF1 | 8832.67 | 0.74 | 0.25 | 2.97 | 0.00 | 0.05 |
| ENSG00000140368.13 | PSTPIP1 | 3470.22 | 0.32 | 0.11 | 2.93 | 0.00 | 0.05 |
| ENSG00000169403.12 | PTAFR | 10619.62 | 0.56 | 0.11 | 5.23 | 0.00 | 0.00 |
| ENSG00000104960.15 | PTOV1 | 972.04 | 0.33 | 0.09 | 3.69 | 0.00 | 0.01 |
| ENSG00000076201.16 | PTPN23 | 1146.35 | 0.28 | 0.09 | 3.18 | 0.00 | 0.03 |
| ENSG00000129317.15 | PUS7L | 898.57 | -0.39 | 0.10 | -3.77 | 0.00 | 0.01 |
| ENSG00000089159.17 | PXN | 18920.43 | 0.43 | 0.14 | 3.08 | 0.00 | 0.04 |
| ENSG00000121350.16 | PYROXD1 | 383.19 | -0.44 | 0.11 | -4.12 | 0.00 | 0.01 |
| ENSG00000107560.12 | RAB11FIP2 | 964.35 | -0.34 | 0.09 | -3.67 | 0.00 | 0.01 |
| ENSG00000105514.8 | RAB3D | 9632.46 | 0.52 | 0.16 | 3.20 | 0.00 | 0.03 |
| ENSG00000167994.13 | RAB3IL1 | 257.90 | 1.15 | 0.37 | 3.12 | 0.00 | 0.04 |
| ENSG00000029725.17 | RABEP1 | 1061.88 | -0.34 | 0.08 | -4.25 | 0.00 | 0.00 |
| ENSG00000144840.10 | RABL3 | 336.96 | -0.45 | 0.11 | -3.98 | 0.00 | 0.01 |
| ENSG00000182185.19 | RAD51B | 194.69 | -0.32 | 0.10 | -3.06 | 0.00 | 0.04 |
| ENSG00000116191.19 | RALGPS2 | 1238.51 | -0.54 | 0.15 | -3.53 | 0.00 | 0.02 |
| ENSG00000137040.10 | RANBP6 | 431.34 | -0.41 | 0.13 | -3.18 | 0.00 | 0.03 |
| ENSG00000132359.15 | RAP1GAP2 | 5256.17 | 0.36 | 0.09 | 3.94 | 0.00 | 0.01 |
| ENSG00000138670.19 | RASGEF1B | 345.76 | -0.41 | 0.14 | -2.95 | 0.00 | 0.05 |
| ENSG00000171777.16 | RASGRP4 | 7213.20 | 0.49 | 0.17 | 2.94 | 0.00 | 0.05 |
| ENSG00000161847.14 | RAVER1 | 1770.57 | 0.26 | 0.08 | 3.16 | 0.00 | 0.03 |
| ENSG00000146587.18 | RBAK | 490.65 | -0.49 | 0.16 | -3.03 | 0.00 | 0.04 |
| ENSG00000139746.16 | RBM26 | 2483.01 | -0.23 | 0.06 | -3.70 | 0.00 | 0.01 |
| ENSG00000188739.15 | RBM34 | 240.25 | -0.36 | 0.11 | -3.34 | 0.00 | 0.02 |
| ENSG00000089682.17 | RBM41 | 515.58 | -0.43 | 0.10 | -4.24 | 0.00 | 0.00 |

| SCC DEG set | | | | | | | |
|---|---|---|---|---|---|---|---|
| Ensembl ID | Gene name | baseMean | Log FC | Log FC SE | Wald statistic value | P-value | Adjusted p-value |
| ENSG00000126254.12 | RBM42 | 798.91 | 0.25 | 0.08 | 2.97 | 0.00 | 0.05 |
| ENSG00000155636.15 | RBM45 | 177.78 | -0.43 | 0.14 | -3.12 | 0.00 | 0.04 |
| ENSG00000135945.10 | REV1 | 644.49 | -0.18 | 0.06 | -3.22 | 0.00 | 0.03 |
| ENSG00000035928.17 | RFC1 | 723.11 | -0.30 | 0.10 | -3.01 | 0.00 | 0.04 |
| ENSG00000132005.9 | RFX1 | 1134.52 | 0.38 | 0.11 | 3.37 | 0.00 | 0.02 |
| ENSG00000237441.10 | RGL2 | 3187.11 | 0.46 | 0.13 | 3.59 | 0.00 | 0.01 |
| ENSG00000185304.16 | RGPD2 | 478.34 | -2.28 | 0.62 | -3.66 | 0.00 | 0.01 |
| ENSG00000166532.16 | RIMKLB | 230.57 | -0.53 | 0.12 | -4.54 | 0.00 | 0.00 |
| ENSG00000039523.20 | RIPOR1 | 2149.82 | 0.37 | 0.11 | 3.34 | 0.00 | 0.02 |
| ENSG00000176623.12 | RMDN1 | 394.62 | -0.33 | 0.08 | -3.86 | 0.00 | 0.01 |
| ENSG00000115841.21 | RMDN2 | 97.48 | -0.51 | 0.14 | -3.78 | 0.00 | 0.01 |
| ENSG00000146083.12 | RNF44 | 6110.42 | 0.33 | 0.10 | 3.45 | 0.00 | 0.02 |
| ENSG00000023191.17 | RNH1 | 1657.83 | 0.23 | 0.07 | 3.36 | 0.00 | 0.02 |
| ENSG00000067836.13 | ROGDI | 737.44 | 0.72 | 0.20 | 3.60 | 0.00 | 0.01 |
| ENSG00000122484.9 | RPAP2 | 682.90 | -0.51 | 0.13 | -3.84 | 0.00 | 0.01 |
| ENSG00000197713.15 | RPE | 676.52 | -0.42 | 0.11 | -3.68 | 0.00 | 0.01 |
| ENSG00000103494.16 | RPGRIP1L | 58.02 | -0.55 | 0.17 | -3.14 | 0.00 | 0.04 |
| ENSG00000153574.9 | RPIA | 1138.56 | 0.76 | 0.26 | 2.94 | 0.00 | 0.05 |
| ENSG00000117676.14 | RPS6KA1 | 6814.50 | 0.43 | 0.12 | 3.46 | 0.00 | 0.02 |
| ENSG00000087302.9 | RTRAF | 563.68 | -0.24 | 0.08 | -3.15 | 0.00 | 0.03 |
| ENSG00000204130.14 | RUFY2 | 335.53 | -0.27 | 0.09 | -3.00 | 0.00 | 0.04 |
| ENSG00000018189.13 | RUFY3 | 726.00 | -0.28 | 0.09 | -2.98 | 0.00 | 0.05 |
| ENSG00000122481.17 | RWDD3 | 83.51 | -0.48 | 0.15 | -3.15 | 0.00 | 0.03 |
| ENSG00000186350.12 | RXRA | 12315.06 | 0.51 | 0.15 | 3.35 | 0.00 | 0.02 |
| ENSG00000163785.13 | RYK | 529.51 | -0.34 | 0.08 | -4.44 | 0.00 | 0.00 |
| ENSG00000126461.15 | SCAF1 | 1936.57 | 0.26 | 0.08 | 3.05 | 0.00 | 0.04 |
| ENSG00000114650.21 | SCAP | 2123.97 | 0.32 | 0.10 | 3.22 | 0.00 | 0.03 |
| ENSG00000140386.13 | SCAPER | 785.26 | -0.50 | 0.16 | -3.07 | 0.00 | 0.04 |
| ENSG00000047634.15 | SCML1 | 154.27 | -0.66 | 0.22 | -2.98 | 0.00 | 0.05 |
| ENSG00000142186.18 | SCYL1 | 1622.87 | 0.38 | 0.10 | 3.66 | 0.00 | 0.01 |
| ENSG00000078808.19 | SDF4 | 2100.38 | 0.17 | 0.05 | 3.21 | 0.00 | 0.03 |
| ENSG00000129657.16 | SEC14L1 | 19122.61 | 0.60 | 0.15 | 3.94 | 0.00 | 0.01 |
| ENSG00000133488.15 | SEC14L4 | 59.03 | 1.21 | 0.35 | 3.46 | 0.00 | 0.02 |
| ENSG00000148396.19 | SEC16A | 4574.57 | 0.38 | 0.10 | 3.96 | 0.00 | 0.01 |
| ENSG00000176986.16 | SEC24C | 2837.61 | 0.20 | 0.07 | 3.00 | 0.00 | 0.04 |
| ENSG00000138018.19 | SELENOI | 453.15 | -0.30 | 0.09 | -3.51 | 0.00 | 0.02 |
| ENSG00000196189.14 | SEMA4A | 6156.42 | 0.59 | 0.19 | 3.11 | 0.00 | 0.04 |
| ENSG00000187764.12 | SEMA4D | 18026.61 | 0.39 | 0.12 | 3.27 | 0.00 | 0.03 |
| ENSG00000138468.16 | SENP7 | 1415.30 | -0.37 | 0.13 | -2.92 | 0.00 | 0.05 |
| ENSG00000139718.12 | SETD1B | 7279.36 | 0.36 | 0.10 | 3.51 | 0.00 | 0.02 |
| ENSG00000099995.19 | SF3A1 | 5276.08 | 0.33 | 0.10 | 3.33 | 0.00 | 0.02 |
| ENSG00000095370.20 | SH2D3C | 4240.61 | 0.41 | 0.13 | 3.09 | 0.00 | 0.04 |
| ENSG00000141985.9 | SH3GL1 | 1620.69 | 0.48 | 0.13 | 3.72 | 0.00 | 0.01 |
| ENSG00000179526.17 | SHARPIN | 1713.66 | 0.68 | 0.23 | 2.96 | 0.00 | 0.05 |
| ENSG00000198892.7 | SHISA4 | 165.55 | 1.42 | 0.48 | 2.97 | 0.00 | 0.05 |
| ENSG00000122376.12 | SHLD2 | 350.08 | -0.44 | 0.14 | -3.09 | 0.00 | 0.04 |
| ENSG00000253251.3 | SHLD3 | 42.81 | -0.53 | 0.18 | -2.99 | 0.00 | 0.04 |
| ENSG00000146414.17 | SHPRH | 1379.28 | -0.42 | 0.11 | -3.63 | 0.00 | 0.01 |
| ENSG00000052723.12 | SIKE1 | 701.06 | -0.38 | 0.10 | -3.73 | 0.00 | 0.01 |
| ENSG00000139146.14 | SINHCAF | 751.33 | -0.25 | 0.08 | -3.08 | 0.00 | 0.04 |
| ENSG00000213445.10 | SIPA1 | 5976.35 | 0.40 | 0.12 | 3.30 | 0.00 | 0.03 |
| ENSG00000157933.10 | SKI | 3158.84 | 0.39 | 0.12 | 3.39 | 0.00 | 0.02 |
| ENSG00000124067.17 | SLC12A4 | 683.72 | 0.36 | 0.07 | 5.14 | 0.00 | 0.00 |
| ENSG00000110446.12 | SLC15A3 | 2513.28 | 0.41 | 0.11 | 3.77 | 0.00 | 0.01 |

| SCC DEG set | | | | | | | |
|---|---|---|---|---|---|---|---|
| Ensembl ID | Gene name | baseMean | Log FC | Log FC SE | Wald statistic value | P-value | Adjusted p-value |
| ENSG00000170190.16 | SLC16A5 | 671.96 | 0.42 | 0.13 | 3.36 | 0.00 | 0.02 |
| ENSG00000173638.19 | SLC19A1 | 4813.99 | 0.51 | 0.16 | 3.11 | 0.00 | 0.04 |
| ENSG00000114120.14 | SLC25A36 | 812.42 | -0.46 | 0.15 | -3.07 | 0.00 | 0.04 |
| ENSG00000164209.17 | SLC25A46 | 889.07 | -0.30 | 0.10 | -3.04 | 0.00 | 0.04 |
| ENSG00000127526.15 | SLC35E1 | 1265.30 | 0.22 | 0.06 | 3.57 | 0.00 | 0.02 |
| ENSG00000196950.14 | SLC39A10 | 700.29 | -0.48 | 0.14 | -3.33 | 0.00 | 0.02 |
| ENSG00000167703.15 | SLC43A2 | 7432.37 | 0.48 | 0.12 | 4.00 | 0.00 | 0.01 |
| ENSG00000129353.15 | SLC44A2 | 10605.34 | 0.35 | 0.10 | 3.35 | 0.00 | 0.02 |
| ENSG00000022567.10 | SLC45A4 | 5445.90 | 0.52 | 0.17 | 3.05 | 0.00 | 0.04 |
| ENSG00000153147.6 | SMARCA5 | 1874.92 | -0.25 | 0.09 | -2.97 | 0.00 | 0.05 |
| ENSG00000163104.18 | SMARCAD1 | 452.47 | -0.44 | 0.14 | -3.18 | 0.00 | 0.03 |
| ENSG00000073584.20 | SMARCE1 | 1557.81 | -0.31 | 0.06 | -4.86 | 0.00 | 0.00 |
| ENSG00000108055.11 | SMC3 | 1048.69 | -0.40 | 0.09 | -4.38 | 0.00 | 0.00 |
| ENSG00000198887.9 | SMC5 | 906.76 | -0.45 | 0.12 | -3.82 | 0.00 | 0.01 |
| ENSG00000163029.16 | SMC6 | 525.81 | -0.50 | 0.12 | -4.24 | 0.00 | 0.00 |
| ENSG00000256537.5 | SMIM10L1 | 284.93 | -0.30 | 0.07 | -4.32 | 0.00 | 0.00 |
| ENSG00000172062.17 | SMN1 | 334.18 | -0.33 | 0.11 | -3.10 | 0.00 | 0.04 |
| ENSG00000164975.16 | SNAPC3 | 846.09 | -0.34 | 0.11 | -2.97 | 0.00 | 0.05 |
| ENSG00000168566.13 | SNRNP48 | 499.02 | -0.39 | 0.09 | -4.43 | 0.00 | 0.00 |
| ENSG00000135317.14 | SNX14 | 2567.26 | -0.28 | 0.09 | -3.25 | 0.00 | 0.03 |
| ENSG00000137642.13 | SORL1 | 72760.84 | 0.60 | 0.15 | 3.90 | 0.00 | 0.01 |
| ENSG00000163611.11 | SPICE1 | 306.97 | -0.44 | 0.13 | -3.41 | 0.00 | 0.02 |
| ENSG00000166145.15 | SPINT1 | 389.17 | 0.57 | 0.17 | 3.23 | 0.00 | 0.03 |
| ENSG00000183018.9 | SPNS2 | 52.85 | 0.61 | 0.20 | 3.04 | 0.00 | 0.04 |
| ENSG00000153914.16 | SREK1 | 1867.92 | -0.34 | 0.07 | -5.06 | 0.00 | 0.00 |
| ENSG00000153006.16 | SREK1IP1 | 502.20 | -0.32 | 0.09 | -3.53 | 0.00 | 0.02 |
| ENSG00000167978.17 | SRRM2 | 24959.33 | 0.31 | 0.10 | 3.06 | 0.00 | 0.04 |
| ENSG00000188529.15 | SRSF10 | 1886.66 | -0.29 | 0.09 | -3.12 | 0.00 | 0.04 |
| ENSG00000116754.14 | SRSF11 | 2598.36 | -0.36 | 0.08 | -4.24 | 0.00 | 0.00 |
| ENSG00000141298.19 | SSH2 | 25166.88 | 0.38 | 0.13 | 2.98 | 0.00 | 0.05 |
| ENSG00000123096.12 | SSPN | 98.91 | -0.65 | 0.22 | -2.92 | 0.00 | 0.05 |
| ENSG00000035720.8 | STAP1 | 56.72 | -1.05 | 0.33 | -3.20 | 0.00 | 0.03 |
| ENSG00000166888.12 | STAT6 | 14366.40 | 0.36 | 0.10 | 3.64 | 0.00 | 0.01 |
| ENSG00000072786.13 | STK10 | 15338.00 | 0.31 | 0.10 | 3.22 | 0.00 | 0.03 |
| ENSG00000144589.22 | STK11IP | 530.60 | 0.29 | 0.09 | 3.06 | 0.00 | 0.04 |
| ENSG00000170310.15 | STX8 | 229.69 | -0.41 | 0.12 | -3.31 | 0.00 | 0.03 |
| ENSG00000198252.12 | STYX | 1077.00 | -0.25 | 0.08 | -2.97 | 0.00 | 0.05 |
| ENSG00000213648.11 | SULT1A4 | 30.01 | 2.60 | 0.53 | 4.92 | 0.00 | 0.00 |
| ENSG00000119760.16 | SUPT7L | 704.24 | -0.19 | 0.06 | -2.94 | 0.00 | 0.05 |
| ENSG00000100647.8 | SUSD6 | 9656.77 | 0.50 | 0.14 | 3.69 | 0.00 | 0.01 |
| ENSG00000198168.9 | SVIP | 296.92 | -0.41 | 0.11 | -3.85 | 0.00 | 0.01 |
| ENSG00000196074.13 | SYCP2 | 118.64 | -0.52 | 0.17 | -3.09 | 0.00 | 0.04 |
| ENSG00000135316.19 | SYNCRIP | 2404.91 | -0.38 | 0.11 | -3.36 | 0.00 | 0.02 |
| ENSG00000143498.18 | TAF1A | 85.76 | -0.53 | 0.14 | -3.74 | 0.00 | 0.01 |
| ENSG00000064313.13 | TAF2 | 1214.18 | -0.18 | 0.06 | -2.92 | 0.00 | 0.05 |
| ENSG00000177156.11 | TALDO1 | 10152.30 | 0.47 | 0.16 | 2.95 | 0.00 | 0.05 |
| ENSG00000183597.16 | TANGO2 | 3539.80 | 0.51 | 0.17 | 2.98 | 0.00 | 0.05 |
| ENSG00000149930.18 | TAOK2 | 1662.25 | 0.23 | 0.06 | 3.64 | 0.00 | 0.01 |
| ENSG00000231925.13 | TAPBP | 22579.12 | 0.41 | 0.14 | 2.98 | 0.00 | 0.05 |
| ENSG00000185418.16 | TARS3 | 228.60 | -0.54 | 0.14 | -3.81 | 0.00 | 0.01 |
| ENSG00000212124.2 | TAS2R19 | 59.52 | -0.61 | 0.14 | -4.37 | 0.00 | 0.00 |
| ENSG00000255837.2 | TAS2R20 | 93.63 | -0.71 | 0.21 | -3.36 | 0.00 | 0.02 |
| ENSG00000256188.3 | TAS2R30 | 152.71 | -0.49 | 0.10 | -4.84 | 0.00 | 0.00 |
| ENSG00000212126.3 | TAS2R50 | 21.14 | -0.86 | 0.25 | -3.40 | 0.00 | 0.02 |

| SCC DEG set | | | | | | | |
|---|---|---|---|---|---|---|---|
| Ensembl ID | Gene name | baseMean | Log FC | Log FC SE | Wald statistic value | P-value | Adjusted p-value |
| ENSG00000132405.19 | TBC1D14 | 5937.84 | 0.47 | 0.15 | 3.17 | 0.00 | 0.03 |
| ENSG00000274611.4 | TBC1D3 | 40.93 | 5.43 | 1.73 | 3.14 | 0.00 | 0.04 |
| ENSG00000146350.14 | TBC1D32 | 128.20 | -0.58 | 0.18 | -3.24 | 0.00 | 0.03 |
| ENSG00000274419.6 | TBC1D3D | 34.29 | 3.93 | 1.13 | 3.48 | 0.00 | 0.02 |
| ENSG00000101849.18 | TBL1X | 5389.95 | 0.36 | 0.11 | 3.20 | 0.00 | 0.03 |
| ENSG00000110719.10 | TCIRG1 | 8719.64 | 0.49 | 0.13 | 3.70 | 0.00 | 0.01 |
| ENSG00000083544.16 | TDRD3 | 369.61 | -0.47 | 0.12 | -3.84 | 0.00 | 0.01 |
| ENSG00000129566.13 | TEP1 | 3914.06 | 0.42 | 0.11 | 3.71 | 0.00 | 0.01 |
| ENSG00000167302.11 | TEPSIN | 1188.02 | 0.34 | 0.10 | 3.32 | 0.00 | 0.03 |
| ENSG00000147601.15 | TERF1 | 916.98 | -0.31 | 0.09 | -3.57 | 0.00 | 0.02 |
| ENSG00000088992.18 | TESC | 1317.40 | 1.03 | 0.27 | 3.78 | 0.00 | 0.01 |
| ENSG00000187605.16 | TET3 | 8427.87 | 0.42 | 0.14 | 3.07 | 0.00 | 0.04 |
| ENSG00000136891.14 | TEX10 | 334.74 | -0.32 | 0.10 | -3.08 | 0.00 | 0.04 |
| ENSG00000144043.12 | TEX261 | 1357.94 | 0.20 | 0.07 | 2.97 | 0.00 | 0.05 |
| ENSG00000029639.11 | TFB1M | 158.62 | -0.44 | 0.13 | -3.36 | 0.00 | 0.02 |
| ENSG00000152291.15 | TGOLN2 | 15527.44 | 0.29 | 0.10 | 2.98 | 0.00 | 0.05 |
| ENSG00000115970.19 | THADA | 1446.36 | -0.24 | 0.07 | -3.42 | 0.00 | 0.02 |
| ENSG00000174796.12 | THAP6 | 466.32 | -0.33 | 0.11 | -2.97 | 0.00 | 0.05 |
| ENSG00000168152.13 | THAP9 | 45.90 | -0.65 | 0.22 | -3.01 | 0.00 | 0.04 |
| ENSG00000169231.13 | THBS3 | 291.16 | 0.44 | 0.12 | 3.55 | 0.00 | 0.02 |
| ENSG00000196407.12 | THEM5 | 45.37 | 1.86 | 0.55 | 3.36 | 0.00 | 0.02 |
| ENSG00000130775.16 | THEMIS2 | 11830.73 | 0.45 | 0.14 | 3.23 | 0.00 | 0.03 |
| ENSG00000079134.13 | THOC1 | 363.82 | -0.46 | 0.13 | -3.43 | 0.00 | 0.02 |
| ENSG00000125676.20 | THOC2 | 1432.72 | -0.27 | 0.08 | -3.45 | 0.00 | 0.02 |
| ENSG00000066654.14 | THUMPD1 | 1032.61 | -0.33 | 0.09 | -3.67 | 0.00 | 0.01 |
| ENSG00000116001.17 | TIA1 | 1546.32 | -0.27 | 0.09 | -2.98 | 0.00 | 0.05 |
| ENSG00000140332.18 | TLE3 | 15648.87 | 0.50 | 0.15 | 3.34 | 0.00 | 0.02 |
| ENSG00000137076.21 | TLN1 | 38533.64 | 0.61 | 0.17 | 3.68 | 0.00 | 0.01 |
| ENSG00000172765.18 | TMCC1 | 1561.63 | 0.48 | 0.15 | 3.11 | 0.00 | 0.04 |
| ENSG00000106460.19 | TMEM106B | 1208.72 | -0.46 | 0.11 | -4.09 | 0.00 | 0.01 |
| ENSG00000166575.17 | TMEM135 | 331.04 | -0.39 | 0.12 | -3.34 | 0.00 | 0.02 |
| ENSG00000164180.14 | TMEM161B | 658.07 | -0.36 | 0.10 | -3.68 | 0.00 | 0.01 |
| ENSG00000182087.14 | TMEM259 | 3190.26 | 0.23 | 0.07 | 3.16 | 0.00 | 0.03 |
| ENSG00000137842.7 | TMEM62 | 264.06 | -0.27 | 0.09 | -2.93 | 0.00 | 0.05 |
| ENSG00000167874.7 | TMEM88 | 152.69 | 0.67 | 0.21 | 3.19 | 0.00 | 0.03 |
| ENSG00000188167.9 | TMPPE | 1031.31 | 0.36 | 0.10 | 3.67 | 0.00 | 0.01 |
| ENSG00000178297.14 | TMPRSS9 | 126.66 | 1.80 | 0.46 | 3.90 | 0.00 | 0.01 |
| ENSG00000139324.12 | TMTC3 | 227.99 | -0.40 | 0.13 | -3.04 | 0.00 | 0.04 |
| ENSG00000185215.11 | TNFAIP2 | 20425.99 | 0.41 | 0.14 | 2.99 | 0.00 | 0.05 |
| ENSG00000173535.15 | TNFRSF10C | 17972.00 | 0.58 | 0.18 | 3.26 | 0.00 | 0.03 |
| ENSG00000157873.18 | TNFRSF14 | 4869.97 | 0.38 | 0.10 | 3.83 | 0.00 | 0.01 |
| ENSG00000049249.9 | TNFRSF9 | 388.51 | 0.62 | 0.19 | 3.21 | 0.00 | 0.03 |
| ENSG00000061938.21 | TNK2 | 4725.68 | 0.36 | 0.08 | 4.41 | 0.00 | 0.00 |
| ENSG00000182095.15 | TNRC18 | 9847.51 | 0.43 | 0.12 | 3.67 | 0.00 | 0.01 |
| ENSG00000198718.13 | TOGARAM1 | 401.55 | -0.32 | 0.11 | -2.98 | 0.00 | 0.05 |
| ENSG00000078902.16 | TOLLIP | 2367.53 | 0.41 | 0.14 | 2.93 | 0.00 | 0.05 |
| ENSG00000177302.15 | TOP3A | 1588.47 | 0.33 | 0.08 | 4.00 | 0.00 | 0.01 |
| ENSG00000078804.13 | TP53INP2 | 483.46 | 0.63 | 0.19 | 3.30 | 0.00 | 0.03 |
| ENSG00000076554.16 | TPD52 | 335.21 | -0.70 | 0.23 | -3.02 | 0.00 | 0.04 |
| ENSG00000134900.12 | TPP2 | 1763.77 | -0.30 | 0.10 | -2.96 | 0.00 | 0.05 |
| ENSG00000146826.17 | TRAPPC14 | 1668.34 | 0.39 | 0.13 | 2.94 | 0.00 | 0.05 |
| ENSG00000146063.20 | TRIM41 | 1025.87 | 0.18 | 0.06 | 3.20 | 0.00 | 0.03 |
| ENSG00000183718.6 | TRIM52 | 1089.57 | -0.30 | 0.09 | -3.37 | 0.00 | 0.02 |
| ENSG00000066651.20 | TRMT11 | 117.26 | -0.50 | 0.12 | -4.04 | 0.00 | 0.01 |

| SCC DEG set | | | | | | | |
|---|---|---|---|---|---|---|---|
| Ensembl ID | Gene name | baseMean | Log FC | Log FC SE | Wald statistic value | P-value | Adjusted p-value |
| ENSG00000122435.11 | TRMT13 | 275.72 | -0.59 | 0.14 | -4.06 | 0.00 | 0.01 |
| ENSG00000072756.17 | TRNT1 | 399.98 | -0.31 | 0.08 | -3.62 | 0.00 | 0.01 |
| ENSG00000165832.6 | TRUB1 | 171.51 | -0.38 | 0.11 | -3.47 | 0.00 | 0.02 |
| ENSG00000103197.18 | TSC2 | 2005.41 | 0.42 | 0.09 | 4.73 | 0.00 | 0.00 |
| ENSG00000168785.8 | TSPAN5 | 1878.83 | 0.89 | 0.30 | 2.94 | 0.00 | 0.05 |
| ENSG00000143643.13 | TTC13 | 654.48 | -0.29 | 0.08 | -3.65 | 0.00 | 0.01 |
| ENSG00000163728.11 | TTC14 | 2128.32 | -0.44 | 0.12 | -3.78 | 0.00 | 0.01 |
| ENSG00000052841.15 | TTC17 | 2374.94 | -0.14 | 0.05 | -2.93 | 0.00 | 0.05 |
| ENSG00000123607.16 | TTC21B | 1413.78 | -0.33 | 0.09 | -3.56 | 0.00 | 0.02 |
| ENSG00000155158.20 | TTC39B | 739.21 | -0.40 | 0.13 | -3.07 | 0.00 | 0.04 |
| ENSG00000165533.19 | TTC8 | 25.62 | -0.53 | 0.18 | -2.92 | 0.00 | 0.05 |
| ENSG00000214021.17 | TTLL3 | 2464.69 | 0.30 | 0.10 | 3.07 | 0.00 | 0.04 |
| ENSG00000167552.15 | TUBA1A | 6169.09 | 0.51 | 0.17 | 2.98 | 0.00 | 0.05 |
| ENSG00000074935.14 | TUBE1 | 186.50 | -0.51 | 0.17 | -3.04 | 0.00 | 0.04 |
| ENSG00000105397.14 | TYK2 | 6013.28 | 0.42 | 0.09 | 4.43 | 0.00 | 0.00 |
| ENSG00000162971.11 | TYW5 | 402.29 | -0.45 | 0.12 | -3.60 | 0.00 | 0.01 |
| ENSG00000063244.13 | U2AF2 | 2967.22 | 0.24 | 0.07 | 3.29 | 0.00 | 0.03 |
| ENSG00000163714.18 | U2SURP | 1897.57 | -0.44 | 0.09 | -5.02 | 0.00 | 0.00 |
| ENSG00000130985.17 | UBA1 | 5022.74 | 0.44 | 0.12 | 3.61 | 0.00 | 0.01 |
| ENSG00000081307.15 | UBA5 | 321.46 | -0.40 | 0.13 | -3.22 | 0.00 | 0.03 |
| ENSG00000169139.12 | UBE2V2 | 330.37 | -0.36 | 0.12 | -3.00 | 0.00 | 0.04 |
| ENSG00000114062.22 | UBE3A | 2057.52 | -0.32 | 0.09 | -3.64 | 0.00 | 0.01 |
| ENSG00000127481.15 | UBR4 | 8701.52 | 0.35 | 0.10 | 3.42 | 0.00 | 0.02 |
| ENSG00000116750.14 | UCHL5 | 495.50 | -0.47 | 0.09 | -4.94 | 0.00 | 0.00 |
| ENSG00000014123.10 | UFL1 | 538.04 | -0.38 | 0.12 | -3.09 | 0.00 | 0.04 |
| ENSG00000120686.12 | UFM1 | 584.05 | -0.38 | 0.11 | -3.47 | 0.00 | 0.02 |
| ENSG00000109775.11 | UFSP2 | 125.34 | -0.41 | 0.13 | -3.21 | 0.00 | 0.03 |
| ENSG00000136731.13 | UGGT1 | 3003.28 | 0.33 | 0.09 | 3.79 | 0.00 | 0.01 |
| ENSG00000147854.17 | UHRF2 | 1466.31 | -0.33 | 0.09 | -3.59 | 0.00 | 0.01 |
| ENSG00000177169.10 | ULK1 | 3768.28 | 0.44 | 0.15 | 2.96 | 0.00 | 0.05 |
| ENSG00000109103.12 | UNC119 | 2189.37 | 0.38 | 0.10 | 3.72 | 0.00 | 0.01 |
| ENSG00000092929.12 | UNC13D | 6514.90 | 0.42 | 0.12 | 3.46 | 0.00 | 0.02 |
| ENSG00000140553.18 | UNC45A | 733.65 | 0.27 | 0.08 | 3.43 | 0.00 | 0.02 |
| ENSG00000005007.13 | UPF1 | 4071.71 | 0.38 | 0.11 | 3.48 | 0.00 | 0.02 |
| ENSG00000125351.13 | UPF3B | 122.50 | -0.45 | 0.14 | -3.11 | 0.00 | 0.04 |
| ENSG00000156256.15 | USP16 | 607.02 | -0.41 | 0.10 | -4.07 | 0.00 | 0.01 |
| ENSG00000172046.20 | USP19 | 1958.14 | 0.33 | 0.10 | 3.17 | 0.00 | 0.03 |
| ENSG00000124422.12 | USP22 | 3745.63 | 0.26 | 0.09 | 3.09 | 0.00 | 0.04 |
| ENSG00000170242.19 | USP47 | 1163.73 | -0.39 | 0.08 | -5.13 | 0.00 | 0.00 |
| ENSG00000145390.12 | USP53 | 477.98 | -0.86 | 0.24 | -3.57 | 0.00 | 0.02 |
| ENSG00000147679.12 | UTP23 | 362.62 | -0.31 | 0.10 | -3.15 | 0.00 | 0.03 |
| ENSG00000117533.15 | VAMP4 | 592.82 | -0.30 | 0.09 | -3.12 | 0.00 | 0.04 |
| ENSG00000151650.8 | VENTX | 217.64 | 0.59 | 0.17 | 3.59 | 0.00 | 0.01 |
| ENSG00000197969.14 | VPS13A | 1703.00 | -0.38 | 0.12 | -3.21 | 0.00 | 0.03 |
| ENSG00000167987.11 | VPS37C | 847.88 | 0.56 | 0.10 | 5.60 | 0.00 | 0.00 |
| ENSG00000128815.20 | WDFY4 | 5810.38 | 0.41 | 0.14 | 3.04 | 0.00 | 0.04 |
| ENSG00000157796.18 | WDR19 | 211.97 | -0.34 | 0.11 | -2.93 | 0.00 | 0.05 |
| ENSG00000118965.16 | WDR35 | 166.90 | -0.37 | 0.10 | -3.76 | 0.00 | 0.01 |
| ENSG00000091157.15 | WDR7 | 1461.59 | -0.17 | 0.06 | -2.98 | 0.00 | 0.05 |
| ENSG00000082068.9 | WDR70 | 456.70 | -0.33 | 0.08 | -4.45 | 0.00 | 0.00 |
| ENSG00000140006.12 | WDR89 | 141.29 | -0.56 | 0.19 | -2.94 | 0.00 | 0.05 |
| ENSG00000196151.11 | WDSUB1 | 125.81 | -0.29 | 0.08 | -3.60 | 0.00 | 0.01 |
| ENSG00000165392.11 | WRN | 336.51 | -0.26 | 0.09 | -2.96 | 0.00 | 0.05 |
| ENSG00000123124.14 | WWP1 | 1039.46 | -0.32 | 0.10 | -3.03 | 0.00 | 0.04 |

| SCC DEG set | | | | | | | |
|---|---|---|---|---|---|---|---|
| Ensembl ID | Gene name | baseMean | Log FC | Log FC SE | Wald statistic value | P-value | Adjusted p-value |
| ENSG00000158156.9 | XKR8 | 1811.01 | 0.41 | 0.11 | 3.84 | 0.00 | 0.01 |
| ENSG00000136936.11 | XPA | 98.53 | -0.48 | 0.16 | -2.98 | 0.00 | 0.05 |
| ENSG00000073050.12 | XRCC1 | 499.58 | 0.24 | 0.06 | 3.87 | 0.00 | 0.01 |
| ENSG00000176105.14 | YES1 | 128.71 | -0.65 | 0.15 | -4.31 | 0.00 | 0.00 |
| ENSG00000236287.8 | ZBED5 | 848.41 | -0.40 | 0.13 | -3.01 | 0.00 | 0.04 |
| ENSG00000126804.14 | ZBTB1 | 1282.41 | -0.39 | 0.11 | -3.64 | 0.00 | 0.01 |
| ENSG00000171448.9 | ZBTB26 | 285.61 | -0.32 | 0.10 | -3.15 | 0.00 | 0.03 |
| ENSG00000177888.8 | ZBTB41 | 447.55 | -0.52 | 0.17 | -3.00 | 0.00 | 0.04 |
| ENSG00000147905.18 | ZCCHC7 | 422.33 | -0.33 | 0.11 | -3.13 | 0.00 | 0.04 |
| ENSG00000139168.9 | ZCRB1 | 253.73 | -0.27 | 0.08 | -3.22 | 0.00 | 0.03 |
| ENSG00000175893.12 | ZDHHC21 | 601.56 | -0.65 | 0.14 | -4.66 | 0.00 | 0.00 |
| ENSG00000142065.14 | ZFP14 | 378.54 | -0.48 | 0.15 | -3.19 | 0.00 | 0.03 |
| ENSG00000180787.6 | ZFP3 | 301.16 | -0.52 | 0.17 | -3.10 | 0.00 | 0.04 |
| ENSG00000100711.14 | ZFYVE21 | 326.68 | 0.22 | 0.07 | 3.10 | 0.00 | 0.04 |
| ENSG00000138658.16 | ZGRF1 | 157.18 | -0.41 | 0.14 | -3.01 | 0.00 | 0.04 |
| ENSG00000189298.14 | ZKSCAN3 | 253.33 | -0.32 | 0.10 | -3.13 | 0.00 | 0.04 |
| ENSG00000187626.9 | ZKSCAN4 | 275.26 | -0.28 | 0.08 | -3.35 | 0.00 | 0.02 |
| ENSG00000166432.16 | ZMAT1 | 580.75 | -0.51 | 0.16 | -3.16 | 0.00 | 0.03 |
| ENSG00000108175.19 | ZMIZ1 | 4719.95 | 0.37 | 0.12 | 2.97 | 0.00 | 0.05 |
| ENSG00000197056.11 | ZMYM1 | 271.58 | -0.35 | 0.11 | -3.27 | 0.00 | 0.03 |
| ENSG00000121741.17 | ZMYM2 | 3207.29 | -0.26 | 0.07 | -3.89 | 0.00 | 0.01 |
| ENSG00000132950.19 | ZMYM5 | 458.12 | -0.32 | 0.10 | -3.18 | 0.00 | 0.03 |
| ENSG00000213762.12 | ZNF134 | 425.96 | -0.40 | 0.13 | -3.22 | 0.00 | 0.03 |
| ENSG00000196646.12 | ZNF136 | 491.14 | -0.31 | 0.09 | -3.35 | 0.00 | 0.02 |
| ENSG00000197008.10 | ZNF138 | 169.10 | -0.75 | 0.21 | -3.63 | 0.00 | 0.01 |
| ENSG00000105708.9 | ZNF14 | 246.47 | -0.59 | 0.13 | -4.62 | 0.00 | 0.00 |
| ENSG00000196387.10 | ZNF140 | 294.46 | -0.43 | 0.08 | -5.59 | 0.00 | 0.00 |
| ENSG00000131127.14 | ZNF141 | 904.16 | -0.47 | 0.15 | -3.11 | 0.00 | 0.04 |
| ENSG00000167635.12 | ZNF146 | 662.06 | -0.45 | 0.14 | -3.21 | 0.00 | 0.03 |
| ENSG00000186272.13 | ZNF17 | 198.64 | -0.44 | 0.12 | -3.62 | 0.00 | 0.01 |
| ENSG00000167384.11 | ZNF180 | 231.33 | -0.54 | 0.13 | -4.18 | 0.00 | 0.00 |
| ENSG00000197841.15 | ZNF181 | 126.20 | -0.61 | 0.20 | -3.06 | 0.00 | 0.04 |
| ENSG00000147118.12 | ZNF182 | 289.35 | -0.32 | 0.11 | -2.99 | 0.00 | 0.04 |
| ENSG00000096654.16 | ZNF184 | 151.88 | -0.52 | 0.16 | -3.32 | 0.00 | 0.03 |
| ENSG00000136870.11 | ZNF189 | 481.76 | -0.35 | 0.12 | -2.98 | 0.00 | 0.05 |
| ENSG00000005801.18 | ZNF195 | 657.61 | -0.45 | 0.12 | -3.93 | 0.00 | 0.01 |
| ENSG00000186448.15 | ZNF197 | 368.12 | -0.57 | 0.09 | -6.69 | 0.00 | 0.00 |
| ENSG00000160321.16 | ZNF208 | 47.02 | -1.15 | 0.34 | -3.43 | 0.00 | 0.02 |
| ENSG00000121417.14 | ZNF211 | 347.62 | -0.33 | 0.10 | -3.19 | 0.00 | 0.03 |
| ENSG00000159905.15 | ZNF221 | 25.26 | -0.63 | 0.20 | -3.18 | 0.00 | 0.03 |
| ENSG00000256294.8 | ZNF225 | 186.41 | -0.45 | 0.12 | -3.80 | 0.00 | 0.01 |
| ENSG00000167380.17 | ZNF226 | 498.72 | -0.56 | 0.13 | -4.35 | 0.00 | 0.00 |
| ENSG00000131115.16 | ZNF227 | 349.96 | -0.40 | 0.12 | -3.35 | 0.00 | 0.02 |
| ENSG00000159917.17 | ZNF235 | 249.67 | -0.46 | 0.12 | -3.69 | 0.00 | 0.01 |
| ENSG00000175395.16 | ZNF25 | 253.05 | -0.36 | 0.11 | -3.43 | 0.00 | 0.02 |
| ENSG00000256771.5 | ZNF253 | 223.83 | -0.64 | 0.17 | -3.70 | 0.00 | 0.01 |
| ENSG00000197134.13 | ZNF257 | 50.60 | -0.79 | 0.26 | -3.01 | 0.00 | 0.04 |
| ENSG00000254004.7 | ZNF260 | 256.03 | -0.59 | 0.17 | -3.56 | 0.00 | 0.02 |
| ENSG00000090612.22 | ZNF268 | 332.37 | -0.51 | 0.12 | -4.05 | 0.00 | 0.01 |
| ENSG00000198039.12 | ZNF273 | 161.06 | -0.65 | 0.17 | -3.75 | 0.00 | 0.01 |
| ENSG00000137871.21 | ZNF280D | 708.00 | -0.47 | 0.15 | -3.07 | 0.00 | 0.04 |
| ENSG00000167637.18 | ZNF283 | 174.88 | -0.46 | 0.11 | -4.20 | 0.00 | 0.00 |
| ENSG00000186026.7 | ZNF284 | 117.52 | -0.42 | 0.13 | -3.19 | 0.00 | 0.03 |
| ENSG00000089335.22 | ZNF302 | 277.83 | -0.67 | 0.18 | -3.62 | 0.00 | 0.01 |

| SCC DEG set | | | | | | | |
|---|---|---|---|---|---|---|---|
| Ensembl ID | Gene name | baseMean | Log FC | Log FC SE | Wald statistic value | P-value | Adjusted p-value |
| ENSG00000205903.7 | ZNF316 | 1735.39 | 0.31 | 0.10 | 3.16 | 0.00 | 0.03 |
| ENSG00000162664.17 | ZNF326 | 613.16 | -0.33 | 0.10 | -3.41 | 0.00 | 0.02 |
| ENSG00000196693.15 | ZNF33B | 657.04 | -0.56 | 0.15 | -3.60 | 0.00 | 0.01 |
| ENSG00000197937.13 | ZNF347 | 270.44 | -0.71 | 0.19 | -3.72 | 0.00 | 0.01 |
| ENSG00000169981.11 | ZNF35 | 81.49 | -0.53 | 0.14 | -3.82 | 0.00 | 0.01 |
| ENSG00000178338.11 | ZNF354B | 158.89 | -0.49 | 0.16 | -3.04 | 0.00 | 0.04 |
| ENSG00000177932.7 | ZNF354C | 178.14 | -0.65 | 0.20 | -3.26 | 0.00 | 0.03 |
| ENSG00000178175.12 | ZNF366 | 157.55 | 0.68 | 0.22 | 3.03 | 0.00 | 0.04 |
| ENSG00000075407.19 | ZNF37A | 668.19 | -0.62 | 0.12 | -5.05 | 0.00 | 0.00 |
| ENSG00000105136.22 | ZNF419 | 224.56 | -0.49 | 0.16 | -3.08 | 0.00 | 0.04 |
| ENSG00000130818.12 | ZNF426 | 376.83 | -0.30 | 0.10 | -3.04 | 0.00 | 0.04 |
| ENSG00000256087.7 | ZNF432 | 167.36 | -0.47 | 0.15 | -3.26 | 0.00 | 0.03 |
| ENSG00000197857.15 | ZNF44 | 461.73 | -0.50 | 0.13 | -3.80 | 0.00 | 0.01 |
| ENSG00000197044.11 | ZNF441 | 212.17 | -0.75 | 0.18 | -4.08 | 0.00 | 0.01 |
| ENSG00000180855.16 | ZNF443 | 84.98 | -0.71 | 0.23 | -3.11 | 0.00 | 0.04 |
| ENSG00000124459.12 | ZNF45 | 292.81 | -0.41 | 0.12 | -3.33 | 0.00 | 0.02 |
| ENSG00000198464.14 | ZNF480 | 231.05 | -0.70 | 0.17 | -4.17 | 0.00 | 0.00 |
| ENSG00000127081.14 | ZNF484 | 549.31 | -0.44 | 0.13 | -3.53 | 0.00 | 0.02 |
| ENSG00000198298.13 | ZNF485 | 43.05 | -0.56 | 0.19 | -2.92 | 0.00 | 0.05 |
| ENSG00000196653.12 | ZNF502 | 38.23 | -0.81 | 0.28 | -2.95 | 0.00 | 0.05 |
| ENSG00000081665.14 | ZNF506 | 1633.04 | -0.33 | 0.10 | -3.25 | 0.00 | 0.03 |
| ENSG00000168813.17 | ZNF507 | 983.81 | -0.36 | 0.10 | -3.76 | 0.00 | 0.01 |
| ENSG00000081386.13 | ZNF510 | 462.53 | -0.55 | 0.13 | -4.23 | 0.00 | 0.00 |
| ENSG00000178163.8 | ZNF518B | 1120.68 | -0.28 | 0.08 | -3.45 | 0.00 | 0.02 |
| ENSG00000171817.17 | ZNF540 | 81.44 | -0.76 | 0.21 | -3.62 | 0.00 | 0.01 |
| ENSG00000152433.15 | ZNF547 | 36.27 | -0.56 | 0.17 | -3.18 | 0.00 | 0.03 |
| ENSG00000130544.12 | ZNF557 | 601.78 | -0.24 | 0.08 | -3.23 | 0.00 | 0.03 |
| ENSG00000188321.14 | ZNF559 | 367.65 | -0.62 | 0.17 | -3.63 | 0.00 | 0.01 |
| ENSG00000171469.11 | ZNF561 | 531.57 | -0.29 | 0.09 | -3.40 | 0.00 | 0.02 |
| ENSG00000198453.14 | ZNF568 | 134.85 | -0.54 | 0.18 | -2.97 | 0.00 | 0.05 |
| ENSG00000180938.6 | ZNF572 | 38.05 | -0.58 | 0.19 | -3.01 | 0.00 | 0.04 |
| ENSG00000018869.18 | ZNF582 | 51.61 | -0.58 | 0.15 | -3.85 | 0.00 | 0.01 |
| ENSG00000198440.9 | ZNF583 | 102.22 | -0.76 | 0.19 | -4.03 | 0.00 | 0.01 |
| ENSG00000166716.10 | ZNF592 | 5374.31 | 0.36 | 0.11 | 3.28 | 0.00 | 0.03 |
| ENSG00000180626.10 | ZNF594 | 244.27 | -0.75 | 0.18 | -4.18 | 0.00 | 0.00 |
| ENSG00000167981.7 | ZNF597 | 187.52 | -0.34 | 0.10 | -3.35 | 0.00 | 0.02 |
| ENSG00000196458.11 | ZNF605 | 297.19 | -0.60 | 0.19 | -3.09 | 0.00 | 0.04 |
| ENSG00000142556.19 | ZNF614 | 166.02 | -0.50 | 0.17 | -2.99 | 0.00 | 0.04 |
| ENSG00000197619.15 | ZNF615 | 207.71 | -0.46 | 0.14 | -3.36 | 0.00 | 0.02 |
| ENSG00000204611.7 | ZNF616 | 187.79 | -0.40 | 0.13 | -3.01 | 0.00 | 0.04 |
| ENSG00000197566.10 | ZNF624 | 137.25 | -0.68 | 0.15 | -4.62 | 0.00 | 0.00 |
| ENSG00000198551.10 | ZNF627 | 149.59 | -0.48 | 0.12 | -3.94 | 0.00 | 0.01 |
| ENSG00000122482.22 | ZNF644 | 1200.66 | -0.49 | 0.13 | -3.69 | 0.00 | 0.01 |
| ENSG00000144792.9 | ZNF660 | 34.71 | -0.95 | 0.20 | -4.69 | 0.00 | 0.00 |
| ENSG00000198046.13 | ZNF667 | 23.18 | -1.26 | 0.40 | -3.17 | 0.00 | 0.03 |
| ENSG00000197372.10 | ZNF675 | 212.55 | -0.65 | 0.19 | -3.40 | 0.00 | 0.02 |
| ENSG00000181450.18 | ZNF678 | 296.13 | -0.61 | 0.16 | -3.79 | 0.00 | 0.01 |
| ENSG00000173041.12 | ZNF680 | 179.61 | -0.78 | 0.17 | -4.48 | 0.00 | 0.00 |
| ENSG00000197124.12 | ZNF682 | 42.12 | -0.70 | 0.24 | -2.95 | 0.00 | 0.05 |
| ENSG00000171163.16 | ZNF692 | 407.35 | 0.34 | 0.10 | 3.59 | 0.00 | 0.01 |
| ENSG00000196110.8 | ZNF699 | 426.67 | -0.32 | 0.08 | -3.85 | 0.00 | 0.01 |
| ENSG00000196757.8 | ZNF700 | 863.09 | -0.37 | 0.12 | -3.20 | 0.00 | 0.03 |
| ENSG00000140548.10 | ZNF710 | 2698.78 | 0.44 | 0.10 | 4.37 | 0.00 | 0.00 |
| ENSG00000160352.16 | ZNF714 | 329.51 | -0.41 | 0.13 | -3.08 | 0.00 | 0.04 |

| SCC DEG set | | | | | | | |
|---|---|---|---|---|---|---|---|
| Ensembl ID | Gene name | baseMean | Log FC | Log FC SE | Wald statistic value | P-value | Adjusted p-value |
| ENSG00000197302.11 | ZNF720 | 405.86 | -0.49 | 0.12 | -4.10 | 0.00 | 0.01 |
| ENSG00000182903.16 | ZNF721 | 523.77 | -0.67 | 0.18 | -3.76 | 0.00 | 0.01 |
| ENSG00000237440.9 | ZNF737 | 692.57 | -0.62 | 0.15 | -4.05 | 0.00 | 0.01 |
| ENSG00000172687.14 | ZNF738 | 176.01 | -0.53 | 0.17 | -3.06 | 0.00 | 0.04 |
| ENSG00000162086.16 | ZNF75A | 512.52 | -0.42 | 0.11 | -3.89 | 0.00 | 0.01 |
| ENSG00000160336.17 | ZNF761 | 192.97 | -0.51 | 0.14 | -3.56 | 0.00 | 0.02 |
| ENSG00000196214.11 | ZNF766 | 399.93 | -0.35 | 0.10 | -3.52 | 0.00 | 0.02 |
| ENSG00000175691.9 | ZNF77 | 52.05 | -0.53 | 0.14 | -3.78 | 0.00 | 0.01 |
| ENSG00000128000.17 | ZNF780B | 912.41 | -0.47 | 0.14 | -3.26 | 0.00 | 0.03 |
| ENSG00000196597.13 | ZNF782 | 309.62 | -0.49 | 0.16 | -3.02 | 0.00 | 0.04 |
| ENSG00000197863.9 | ZNF790 | 69.51 | -0.70 | 0.19 | -3.79 | 0.00 | 0.01 |
| ENSG00000173875.14 | ZNF791 | 908.88 | -0.28 | 0.08 | -3.65 | 0.00 | 0.01 |
| ENSG00000196466.10 | ZNF799 | 159.50 | -0.43 | 0.13 | -3.36 | 0.00 | 0.02 |
| ENSG00000048405.11 | ZNF800 | 1494.83 | -0.28 | 0.09 | -3.17 | 0.00 | 0.03 |
| ENSG00000180257.14 | ZNF816 | 141.27 | -0.58 | 0.17 | -3.47 | 0.00 | 0.02 |
| ENSG00000198040.11 | ZNF84 | 453.99 | -0.59 | 0.16 | -3.72 | 0.00 | 0.01 |
| ENSG00000197608.12 | ZNF841 | 491.07 | -0.36 | 0.12 | -3.11 | 0.00 | 0.04 |
| ENSG00000196605.7 | ZNF846 | 110.93 | -0.49 | 0.13 | -3.69 | 0.00 | 0.01 |
| ENSG00000167232.14 | ZNF91 | 1267.32 | -0.71 | 0.18 | -3.95 | 0.00 | 0.01 |
| ENSG00000146757.14 | ZNF92 | 194.48 | -0.65 | 0.20 | -3.30 | 0.00 | 0.03 |
| ENSG00000188372.15 | ZP3 | 31.39 | 0.89 | 0.25 | 3.52 | 0.00 | 0.02 |
| ENSG00000132485.14 | ZRANB2 | 1560.40 | -0.41 | 0.12 | -3.46 | 0.00 | 0.02 |
| ENSG00000121988.18 | ZRANB3 | 64.43 | -0.53 | 0.18 | -2.97 | 0.00 | 0.05 |
| ENSG00000158691.15 | ZSCAN12 | 335.25 | -0.63 | 0.17 | -3.67 | 0.00 | 0.01 |
| ENSG00000196812.6 | ZSCAN16 | 123.53 | -0.42 | 0.12 | -3.51 | 0.00 | 0.02 |
| ENSG00000197062.12 | ZSCAN26 | 248.59 | -0.40 | 0.09 | -4.60 | 0.00 | 0.00 |
| ENSG00000186814.14 | ZSCAN30 | 339.85 | -0.41 | 0.12 | -3.49 | 0.00 | 0.02 |
| ENSG00000214655.11 | ZSWIM8 | 3120.74 | 0.42 | 0.11 | 3.92 | 0.00 | 0.01 |
| ENSG00000074755.15 | ZZEF1 | 6286.52 | 0.22 | 0.07 | 3.10 | 0.00 | 0.04 |
| ENSG00000036549.13 | ZZZ3 | 691.35 | -0.43 | 0.13 | -3.40 | 0.00 | 0.02 |

| DEG set Name | Reactome Pathway | Reactome Set Size | InSet Intersection Size | FDR (adj p-value) | Gene InSet Intersection |
|---|---|---|---|---|---|
| SCC DEG set_Down | [R-HSA-74160] Gene expression (Transcription) | 1498 | 115 | 0.00E+00 | ANAPC10; ANAPC4; ATR; BDP1; BMI1; CAMK2D; CENPJ; CHM; CNOT7; E2F5; GTF3C3; INTS13; KRBOX5; MAGOHB; MNAT1; MRE11; MSH2; MTERF1; MTF2; NR2C1; PSIP1; PSMD14; PSMD5; RPAP2; SMARCA5; SMARCE1; SNAPC3; SRSF11; TAF1A; TAF2; THOC1; THOC2; UPF3B; WRN; WWP1; YES1; ZFP14; ZKSCAN3; ZKSCAN4; ZNF136; ZNF138; ZNF14; ZNF140; ZNF141; ZNF17; ZNF180; ZNF184; ZNF189; ZNF195; ZNF197; ZNF208; ZNF211; ZNF221; ZNF225; ZNF226; ZNF227; ZNF235; ZNF25; ZNF253; ZNF257; ZNF268; ZNF273; ZNF302; ZNF33B; ZNF347; ZNF354B; ZNF354C; ZNF37A; ZNF419; ZNF426; ZNF432; ZNF441; ZNF443; ZNF45; ZNF480; ZNF484; ZNF485; ZNF506; ZNF510; ZNF540; ZNF547; ZNF557; ZNF559; ZNF561; ZNF568; ZNF582; ZNF583; ZNF597; ZNF605; ZNF614; ZNF615; ZNF616; ZNF624; ZNF627; ZNF660; ZNF667; ZNF675; ZNF678; ZNF680; ZNF682; ZNF699; ZNF700; ZNF714; ZNF721; ZNF737; ZNF738; ZNF75A; ZNF761; ZNF77; ZNF782; ZNF790; ZNF791; ZNF799; ZNF92; ZZZ3 |
| SCC DEG set_Down | [R-HSA-73857] RNA Polymerase II Transcription | 1334 | 107 | 0.00E+00 | ANAPC10; ANAPC4; ATR; BMI1; CAMK2D; CENPJ; CHM; CNOT7; E2F5; INTS13; KRBOX5; MAGOHB; MNAT1; MRE11; MSH2; NR2C1; PSMD14; PSMD5; RPAP2; SMARCE1; SNAPC3; SRSF11; TAF2; THOC1; THOC2; UPF3B; WRN; WWP1; YES1; ZFP14; ZKSCAN3; ZKSCAN4; ZNF136; ZNF138; ZNF14; ZNF140; ZNF141; ZNF17; ZNF180; ZNF184; ZNF189; ZNF195; ZNF197; ZNF208; ZNF211; ZNF221; ZNF225; ZNF226; ZNF227; ZNF235; ZNF25; ZNF253; ZNF257; ZNF268; ZNF273; ZNF302; ZNF33B; ZNF347; ZNF354B; ZNF354C; ZNF37A; ZNF419; ZNF426; ZNF432; ZNF441; ZNF443; ZNF45; ZNF480; ZNF484; ZNF485; ZNF506; ZNF510; ZNF540; ZNF547; ZNF557; ZNF559; ZNF561; ZNF568; ZNF582; ZNF583; ZNF597; ZNF605; ZNF614; ZNF615; ZNF616; ZNF624; ZNF627; ZNF660; ZNF667; ZNF675; ZNF678; ZNF680; ZNF682; ZNF699; ZNF700; ZNF714; ZNF721; ZNF737; ZNF738; ZNF75A; ZNF761; ZNF77; ZNF782; ZNF790; ZNF791; ZNF799; ZNF92 |
| SCC DEG set_Down | [R-HSA-212436] Generic Transcription Pathway | 1212 | 99 | 0.00E+00 | ANAPC10; ANAPC4; ATR; BMI1; CAMK2D; CENPJ; CHM; CNOT7; E2F5; KRBOX5; MNAT1; MRE11; MSH2; NR2C1; PSMD14; PSMD5; SMARCE1; TAF2; WRN; WWP1; YES1; ZFP14; ZKSCAN3; ZKSCAN4; ZNF136; ZNF138; ZNF14; ZNF140; ZNF141; ZNF17; ZNF180; ZNF184; ZNF189; ZNF195; ZNF197; ZNF208; ZNF211; ZNF221; ZNF225; ZNF226; ZNF227; ZNF235; ZNF25; ZNF253; ZNF257; ZNF268; ZNF273; ZNF302; ZNF33B; ZNF347; ZNF354B; ZNF354C; ZNF37A; ZNF419; ZNF426; ZNF432; ZNF441; ZNF443; ZNF45; ZNF480; ZNF484; ZNF485; ZNF506; ZNF510; ZNF540; ZNF547; ZNF557; ZNF559; ZNF561; ZNF568; ZNF582; ZNF583; ZNF597; ZNF605; ZNF614; ZNF615; ZNF616; ZNF624; ZNF627; ZNF660; ZNF667; ZNF675; ZNF678; ZNF680; ZNF682; ZNF699; ZNF700; ZNF714; ZNF721; ZNF737; ZNF738; ZNF75A; ZNF761; ZNF77; ZNF782; ZNF790; ZNF791; ZNF799; ZNF92 |
| SCC DEG set_Up | [R-HSA-168256] Immune System | 2096 | 77 | 1.11E-06 | ADA2; ADAM8; ADGRE3; ADGRE5; AGER; ALDOA; ANPEP; AP2A2; ARSA; C5AR2; CBL; CD300LB; COTL1; CREBBP; CTSE; FBXL18; FSCN1; GSN; HSPA1A; HSPA1B; ICAM3; ICAM4; IGF2R; ITGAL; ITGAX; JAML; KRT1; LILRB2; MAN2B1; MAVS; MIB2; MRC2; MSN; MYH9; MYO1C; MYO9B; NBEAL2; NCF4; NLRP1; NLRP3; NUP50; ORAI2; PAK1; PDXK; PIK3CD; PITPNA; PLD2; PRDX6; PRG2; PSME3; PSMF1; PSTPIP1; PTAFR; PTEN; PTPN23; RAB3D; RAP1GAP2; RASGRP4; RPS6KA1; SEC24C; SIPA1; SLC44A2; STAT6; STK10; STK11IP; TALDO1; TAPBP; TCIRG1; TNFRSF14; TNFRSF9; TOLLIP; TRIM41; TUBA1A; TYK2; UBA1; UBR4; UNC13D |
| SCC DEG set_Up | [R-HSA-162582] Signal Transduction | 2533 | 76 | 2.67E-03 | ADGRE3; ADGRE5; AGO2; AP2A2; ARAP1; ARAP3; ARHGAP1; ARHGAP25; ARHGAP27; ARHGAP4; ARHGEF1; ARHGEF2; C5AR2; CBL; CDC42EP2; CREBBP; CSNK1G2; DAGLB; DGKZ; E2F4; GGA3; GNA11; GNAI2; GRK2; HDAC10; HGS; ITGA5; KDM4B; KMT2D; LTBP2; MAML3; MAP3K11; MEF2D; MIB2; MRTFA; MYH9; MYO9B; NCF4; NCOR2; NOTCH1; NOTCH2; PAK1; PFN1; PIK3CD; PIK3R5; PIP5K1C; PKN1; PLD2; PLXND1; PPP1CA; PREX1; PSME3; PSMF1; PTAFR; PTEN; PXN; RASGRP4; RGL2; RPS6KA1; RXRA; SH2D2A; SH3GL1; SHARPIN; SKI; SPINT1; STAT6; STK10; TBL1X; TCIRG1; THBS3; TLE3; TLN1; TSC2; TUBA1A; TYK2; ULK1 |
| SCC DEG set_Up | [R-HSA-168249] Innate Immune System | 1116 | 51 | 1.11E-06 | ADA2; ADAM8; ADGRE3; ADGRE5; AGER; ALDOA; ANPEP; AP2A2; ARSA; C5AR2; CD300LB; COTL1; CREBBP; GSN; HSPA1A; HSPA1B; ICAM3; IGF2R; ITGAL; ITGAX; KRT1; LILRB2; MAN2B1; MAVS; MYH9; MYO1C; MYO9B; NBEAL2; NCF4; NLRP1; NLRP3; PAK1; PDXK; PLD2; PRDX6; PRG2; PSME3; PSMF1; PSTPIP1; PTAFR; RAB3D; RASGRP4; RPS6KA1; SLC44A2; STAT6; STK10; STK11IP; TCIRG1; TOLLIP; UBR4; UNC13D |
| SCC DEG set_Up | [R-HSA-1266738] Developmental Biology | 1237 | 38 | 4.20E-02 | AGO2; AP2A2; CREBBP; ITGA5; KDM6B; KMT2D; KRT1; KRT23; KRT77; MAML3; MBP; MED12; MED16; MED25; MED9; MEF2D; MSN; MYH9; MYO9B; NCOR2; NOTCH1; PAK1; PFN1; PIK3CD; PIP5K1C; PITPNA; PLXND1; PSME3; PSMF1; RPS6KA1; RXRA; SEMA4A; SEMA4D; TBL1X; TET3; TLN1; TUBA1A; ZSWIM8 |
| SGRQ DEG set_Down | [R-HSA-168256] Immune System | 2096 | 37 | 5.85E-06 | AIM2; C2; CCR1; CD274; CEACAM1; EIF4G3; FYB1; GBP1; GBP4; GBP5; GBP6; GPR84; GYG1; ICAM1; IFI16; IFI35; IFIH1; IL10; IL27; JAK2; MAP2K6; MCEMP1; MYO10; NECTIN2; PDCD1LG2; PIK3AP1; PTPN12; RIGI; SOS1; SP100; TIFA; TLR5; TREML4; TRIM22; TRIM69; UBR1; VAMP3 |
| SGRQ DEG set_Down | [R-HSA-1280215] Cytokine Signaling in Immune system | 769 | 18 | 3.63E-04 | CCR1; EIF4G3; GBP1; GBP4; GBP5; GBP6; ICAM1; IFI35; IL10; IL27; JAK2; MAP2K6; PTPN12; RIGI; SOS1; SP100; TIFA; TRIM22 |
| SGRQ DEG set_Down | [R-HSA-913531] Interferon Signaling | 267 | 12 | 4.01E-05 | EIF4G3; GBP1; GBP4; GBP5; GBP6; ICAM1; IFI35; JAK2; MAP2K6; RIGI; SP100; TRIM22 |
| SGRQ DEG set_Up | [R-HSA-71291] Metabolism of amino acids and derivatives | 371 | 5 | 1.18E-02 | RPL12; RPL13; RPL13A; RPL3; RPS6 |
| SGRQ DEG set_Up | [R-HSA-72766] Translation | 294 | 5 | 4.36E-03 | RPL12; RPL13; RPL13A; RPL3; RPS6 |
| SGRQ DEG set_Up | [R-HSA-376176] Signaling by ROBO receptors | 217 | 5 | 1.13E-03 | RPL12; RPL13; RPL13A; RPL3; RPS6 |
| SGRQ DEG set_Up | [R-HSA-72312] rRNA processing | 203 | 5 | 8.62E-04 | RPL12; RPL13; RPL13A; RPL3; RPS6 |
| SGRQ DEG set_Up | [R-HSA-8868773] rRNA processing in the nucleus and cytosol | 193 | 5 | 7.11E-04 | RPL12; RPL13; RPL13A; RPL3; RPS6 |
| SGRQ DEG set_Up | [R-HSA-6791226] Major pathway of rRNA processing in the nucleolus and cytosol | 183 | 5 | 5.79E-04 | RPL12; RPL13; RPL13A; RPL3; RPS6 |
| SGRQ DEG set_Up | [R-HSA-9010553] Regulation of expression of SLITs and ROBOs | 171 | 5 | 4.41E-04 | RPL12; RPL13; RPL13A; RPL3; RPS6 |
| SGRQ DEG set_Up | [R-HSA-9711097] Cellular response to starvation | 157 | 5 | 3.09E-04 | RPL12; RPL13; RPL13A; RPL3; RPS6 |
| SGRQ DEG set_Up | [R-HSA-168255] Influenza Infection | 154 | 5 | 2.97E-04 | RPL12; RPL13; RPL13A; RPL3; RPS6 |
| SGRQ DEG set_Up | [R-HSA-168273] Influenza Viral RNA Transcription and Replication | 134 | 5 | 1.61E-04 | RPL12; RPL13; RPL13A; RPL3; RPS6 |
| SGRQ DEG set_Up | [R-HSA-72613] Eukaryotic Translation Initiation | 120 | 5 | 1.00E-04 | RPL12; RPL13; RPL13A; RPL3; RPS6 |
| SGRQ DEG set_Up | [R-HSA-72737] Cap-dependent Translation Initiation | 120 | 5 | 1.00E-04 | RPL12; RPL13; RPL13A; RPL3; RPS6 |
| SGRQ DEG set_Up | [R-HSA-2408522] Selenoamino acid metabolism | 117 | 5 | 1.00E-04 | RPL12; RPL13; RPL13A; RPL3; RPS6 |
| SGRQ DEG set_Up | [R-HSA-927802] Nonsense-Mediated Decay (NMD) | 116 | 5 | 1.00E-04 | RPL12; RPL13; RPL13A; RPL3; RPS6 |
| SGRQ DEG set_Up | [R-HSA-975957] Nonsense Mediated Decay (NMD) enhanced by the Exon Junction Complex (EJC) | 116 | 5 | 1.00E-04 | RPL12; RPL13; RPL13A; RPL3; RPS6 |
| SGRQ DEG set_Up | [R-HSA-1799339] SRP-dependent cotranslational protein targeting to membrane | 113 | 5 | 1.00E-04 | RPL12; RPL13; RPL13A; RPL3; RPS6 |
| SGRQ DEG set_Up | [R-HSA-72706] GTP hydrolysis and joining of the 60S ribosomal subunit | 113 | 5 | 1.00E-04 | RPL12; RPL13; RPL13A; RPL3; RPS6 |
| SGRQ DEG set_Up | [R-HSA-156827] L13a-mediated translational silencing of Ceruloplasmin expression | 112 | 5 | 1.00E-04 | RPL12; RPL13; RPL13A; RPL3; RPS6 |
| SGRQ DEG set_Up | [R-HSA-425393] Transport of inorganic cations/anions and amino acids/oligopeptides | 105 | 3 | 1.22E-02 | SLC4A3; SLC6A19; SLC9A3 |
| SGRQ DEG set_Up | [R-HSA-72689] Formation of a pool of free 40S subunits | 102 | 5 | 9.05E-05 | RPL12; RPL13; RPL13A; RPL3; RPS6 |
| SGRQ DEG set_Up | [R-HSA-9633012] Response of EIF2AK4 (GCN2) to amino acid deficiency | 102 | 5 | 9.05E-05 | RPL12; RPL13; RPL13A; RPL3; RPS6 |
| SGRQ DEG set_Up | [R-HSA-975956] Nonsense Mediated Decay (NMD) independent of the Exon Junction Complex (EJC) | 96 | 5 | 8.95E-05 | RPL12; RPL13; RPL13A; RPL3; RPS6 |
| SGRQ DEG set_Up | [R-HSA-156842] Eukaryotic Translation Elongation | 95 | 5 | 8.95E-05 | RPL12; RPL13; RPL13A; RPL3; RPS6 |
| SGRQ DEG set_Up | [R-HSA-2408557] Selenocysteine synthesis | 94 | 5 | 8.95E-05 | RPL12; RPL13; RPL13A; RPL3; RPS6 |
| SGRQ DEG set_Up | [R-HSA-72764] Eukaryotic Translation Termination | 94 | 5 | 8.95E-05 | RPL12; RPL13; RPL13A; RPL3; RPS6 |
| SGRQ DEG set_Up | [R-HSA-156902] Peptide chain elongation | 90 | 5 | 8.95E-05 | RPL12; RPL13; RPL13A; RPL3; RPS6 |
| SGRQ DEG set_Up | [R-HSA-192823] Viral mRNA Translation | 90 | 5 | 8.95E-05 | RPL12; RPL13; RPL13A; RPL3; RPS6 |
| X-ray DEG set_Down | [R-HSA-168256] Immune System | 2096 | 28 | 2.60E-03 | CD274; CLEC2B; CUL2; ERP44; GBP4; GBP5; HERC5; IFI27; IFI6; IFIH1; IFIT1; IFIT3; IFIT5; JAK2; MX1; OAS1; OAS3; PPM1B; PTPN2; RIGI; RSAD2; SERPING1; SOS1; TBK1; TRIM22; TRIM69; UBR1; XAF1 |
| X-ray DEG set_Down | [R-HSA-1280215] Cytokine Signaling in Immune system | 769 | 20 | 5.06E-06 | GBP4; GBP5; HERC5; IFI27; IFI6; IFIT1; IFIT3; IFIT5; JAK2; MX1; OAS1; OAS3; PPM1B; PTPN2; RIGI; RSAD2; SOS1; TBK1; TRIM22; XAF1 |
| X-ray DEG set_Down | [R-HSA-913531] Interferon Signaling | 267 | 18 | 1.63E-11 | GBP4; GBP5; HERC5; IFI27; IFI6; IFIT1; IFIT3; IFIT5; JAK2; MX1; OAS1; OAS3; PPM1B; PTPN2; RIGI; RSAD2; TRIM22; XAF1 |

| X-ray DEG set | | | | | | | |
|---|---|---|---|---|---|---|---|
| Ensembl ID | Gene name | baseMean | Log FC | Log FC SE | Wald statistic value | P-value | Adjusted p-value |
| ENSG00000152766.6 | ANKRD22 | 749.36 | -2.06 | 0.52 | -3.96 | 0.00 | 0.02 |
| ENSG00000196975.16 | ANXA4 | 698.47 | -0.37 | 0.10 | -3.57 | 0.00 | 0.03 |
| ENSG00000047365.13 | ARAP2 | 3056.64 | -0.71 | 0.16 | -4.55 | 0.00 | 0.01 |
| ENSG00000032219.19 | ARID4A | 1928.46 | -0.40 | 0.12 | -3.36 | 0.00 | 0.05 |
| ENSG00000165997.5 | ARL5B | 724.43 | -0.46 | 0.13 | -3.43 | 0.00 | 0.04 |
| ENSG00000114439.19 | BBX | 2353.55 | -0.33 | 0.09 | -3.80 | 0.00 | 0.02 |
| ENSG00000173681.17 | BCLAF3 | 833.36 | -0.52 | 0.12 | -4.24 | 0.00 | 0.01 |
| ENSG00000117475.14 | BLZF1 | 418.11 | -0.51 | 0.14 | -3.72 | 0.00 | 0.02 |
| ENSG00000166164.16 | BRD7 | 1027.92 | -0.31 | 0.09 | -3.63 | 0.00 | 0.03 |
| ENSG00000072736.19 | NFATC3 | 4644.62 | 0.33 | 0.08 | 3.97 | 0.00 | 0.02 |
| ENSG00000106392.11 | C1GALT1 | 1964.47 | -0.61 | 0.17 | -3.52 | 0.00 | 0.04 |
| ENSG00000113583.8 | C5orf15 | 754.86 | -0.52 | 0.15 | -3.43 | 0.00 | 0.04 |
| ENSG00000147996.17 | CBWD5 | 1299.21 | -0.39 | 0.10 | -3.80 | 0.00 | 0.02 |
| ENSG00000134480.16 | CCNH | 1851.89 | -0.40 | 0.10 | -3.95 | 0.00 | 0.02 |
| ENSG00000082258.13 | CCNT2 | 2439.77 | -0.40 | 0.12 | -3.41 | 0.00 | 0.04 |
| ENSG00000120217.14 | CD274 | 2167.85 | -1.71 | 0.50 | -3.45 | 0.00 | 0.04 |
| ENSG00000134371.14 | CDC73 | 2035.93 | -0.44 | 0.12 | -3.56 | 0.00 | 0.03 |
| ENSG00000174799.11 | CEP135 | 614.85 | -0.60 | 0.14 | -4.42 | 0.00 | 0.01 |
| ENSG00000135315.12 | CEP162 | 308.45 | -0.80 | 0.19 | -4.21 | 0.00 | 0.01 |
| ENSG00000153922.11 | CHD1 | 3599.49 | -0.57 | 0.15 | -3.77 | 0.00 | 0.02 |
| ENSG00000110852.5 | CLEC2B | 958.92 | -0.80 | 0.23 | -3.44 | 0.00 | 0.04 |
| ENSG00000113240.14 | CLK4 | 1896.26 | -0.50 | 0.15 | -3.36 | 0.00 | 0.05 |
| ENSG00000134326.12 | CMPK2 | 1724.51 | -1.62 | 0.47 | -3.48 | 0.00 | 0.04 |
| ENSG00000108094.17 | CUL2 | 812.22 | -0.28 | 0.08 | -3.57 | 0.00 | 0.03 |
| ENSG00000108654.16 | DDX5 | 14805.97 | -0.29 | 0.08 | -3.62 | 0.00 | 0.03 |
| ENSG00000107201.11 | DDX58 | 7070.06 | -1.03 | 0.28 | -3.65 | 0.00 | 0.03 |
| ENSG00000106992.19 | AK1 | 437.67 | 1.00 | 0.27 | 3.73 | 0.00 | 0.02 |
| ENSG00000137628.18 | DDX60 | 4103.30 | -1.43 | 0.36 | -3.92 | 0.00 | 0.02 |
| ENSG00000213047.13 | DENND1B | 1057.41 | -0.54 | 0.14 | -3.98 | 0.00 | 0.02 |
| ENSG00000147202.19 | DIAPH2 | 1104.61 | -0.46 | 0.12 | -3.81 | 0.00 | 0.02 |
| ENSG00000104093.14 | DMXL2 | 8110.65 | -0.62 | 0.14 | -4.34 | 0.00 | 0.01 |
| ENSG00000132294.15 | EFR3A | 2312.33 | -0.40 | 0.11 | -3.74 | 0.00 | 0.02 |
| ENSG00000104412.8 | EMC2 | 410.53 | -0.63 | 0.16 | -4.01 | 0.00 | 0.02 |
| ENSG00000133106.15 | EPSTI1 | 3506.91 | -1.42 | 0.40 | -3.51 | 0.00 | 0.04 |
| ENSG00000023318.9 | ERP44 | 2298.25 | -0.51 | 0.14 | -3.57 | 0.00 | 0.03 |
| ENSG00000090989.18 | EXOC1 | 1247.88 | -0.49 | 0.14 | -3.59 | 0.00 | 0.03 |
| ENSG00000166801.17 | FAM111A | 3387.33 | -0.52 | 0.13 | -3.93 | 0.00 | 0.02 |
| ENSG00000113391.19 | FAM172A | 807.11 | -0.36 | 0.10 | -3.65 | 0.00 | 0.03 |
| ENSG00000219626.9 | FAM228B | 326.57 | -0.50 | 0.12 | -4.01 | 0.00 | 0.02 |
| ENSG00000143756.12 | FBXO28 | 952.55 | -0.30 | 0.08 | -3.74 | 0.00 | 0.02 |
| ENSG00000171055.15 | FEZ2 | 783.27 | -0.42 | 0.11 | -3.69 | 0.00 | 0.03 |
| ENSG00000112367.12 | FIG4 | 2590.29 | -0.50 | 0.14 | -3.48 | 0.00 | 0.04 |
| ENSG00000114650.21 | SCAP | 2080.17 | 0.37 | 0.11 | 3.53 | 0.00 | 0.04 |
| ENSG00000102081.16 | FMR1 | 2433.48 | -0.45 | 0.12 | -3.82 | 0.00 | 0.02 |
| ENSG00000168522.13 | FNTA | 1773.67 | -0.37 | 0.09 | -4.08 | 0.00 | 0.02 |
| ENSG00000162654.9 | GBP4 | 4790.51 | -0.93 | 0.27 | -3.41 | 0.00 | 0.04 |
| ENSG00000154451.15 | GBP5 | 35542.51 | -1.33 | 0.40 | -3.36 | 0.00 | 0.05 |
| ENSG00000062194.16 | GPBP1 | 2648.95 | -0.51 | 0.14 | -3.72 | 0.00 | 0.02 |
| ENSG00000138646.9 | HERC5 | 1635.54 | -1.53 | 0.40 | -3.81 | 0.00 | 0.02 |
| ENSG00000165949.13 | IFI27 | 551.76 | -3.54 | 1.03 | -3.43 | 0.00 | 0.04 |
| ENSG00000137965.11 | IFI44 | 1798.02 | -2.41 | 0.50 | -4.80 | 0.00 | 0.00 |
| ENSG00000137959.17 | IFI44L | 7440.32 | -2.91 | 0.65 | -4.50 | 0.00 | 0.01 |
| ENSG00000126709.16 | IFI6 | 2959.68 | -1.52 | 0.40 | -3.79 | 0.00 | 0.02 |
| ENSG00000115267.9 | IFIH1 | 2409.91 | -1.13 | 0.29 | -3.96 | 0.00 | 0.02 |
| ENSG00000185745.10 | IFIT1 | 2607.18 | -1.63 | 0.48 | -3.40 | 0.00 | 0.04 |
| ENSG00000119917.15 | IFIT3 | 9443.81 | -1.57 | 0.41 | -3.87 | 0.00 | 0.02 |
| ENSG00000152778.10 | IFIT5 | 1919.70 | -1.13 | 0.32 | -3.54 | 0.00 | 0.04 |
| ENSG00000096968.14 | JAK2 | 5541.44 | -0.86 | 0.23 | -3.79 | 0.00 | 0.02 |
| ENSG00000151657.12 | KIN | 575.53 | -0.40 | 0.10 | -3.92 | 0.00 | 0.02 |
| ENSG00000002549.13 | LAP3 | 2399.82 | -1.43 | 0.40 | -3.58 | 0.00 | 0.03 |
| ENSG00000198160.15 | MIER1 | 3214.68 | -0.51 | 0.15 | -3.46 | 0.00 | 0.04 |

| X-ray DEG set | | | | | | | |
|---|---|---|---|---|---|---|---|
| Ensembl ID | Gene name | baseMean | Log FC | Log FC SE | Wald statistic value | P-value | Adjusted p-value |
| ENSG00000158411.11 | MITD1 | 687.71 | -0.47 | 0.13 | -3.71 | 0.00 | 0.02 |
| ENSG00000157601.15 | MX1 | 8414.18 | -1.53 | 0.45 | -3.44 | 0.00 | 0.04 |
| ENSG00000162601.11 | MYSM1 | 1993.54 | -0.33 | 0.10 | -3.41 | 0.00 | 0.04 |
| ENSG00000244754.9 | N4BP2L2 | 12707.55 | -0.44 | 0.11 | -3.86 | 0.00 | 0.02 |
| ENSG00000132780.17 | NASP | 932.95 | -0.44 | 0.12 | -3.83 | 0.00 | 0.02 |
| ENSG00000111912.20 | NCOA7 | 1132.50 | -0.40 | 0.11 | -3.52 | 0.00 | 0.04 |
| ENSG00000151092.18 | NGLY1 | 1111.33 | -0.28 | 0.07 | -3.97 | 0.00 | 0.02 |
| ENSG00000089127.15 | OAS1 | 5755.18 | -1.80 | 0.45 | -3.96 | 0.00 | 0.02 |
| ENSG00000111331.14 | OAS3 | 10931.35 | -1.95 | 0.49 | -4.00 | 0.00 | 0.02 |
| ENSG00000164219.10 | PGGT1B | 1762.84 | -0.49 | 0.12 | -4.02 | 0.00 | 0.02 |
| ENSG00000136147.18 | PHF11 | 1207.45 | -0.60 | 0.16 | -3.74 | 0.00 | 0.02 |
| ENSG00000146247.14 | PHIP | 5016.76 | -0.46 | 0.13 | -3.57 | 0.00 | 0.03 |
| ENSG00000102893.16 | PHKB | 2051.37 | -0.30 | 0.09 | -3.44 | 0.00 | 0.04 |
| ENSG00000132424.17 | PNISR | 5264.71 | -0.41 | 0.09 | -4.40 | 0.00 | 0.01 |
| ENSG00000101751.11 | POLI | 530.30 | -0.40 | 0.11 | -3.61 | 0.00 | 0.03 |
| ENSG00000122008.15 | POLK | 949.20 | -0.51 | 0.13 | -3.87 | 0.00 | 0.02 |
| ENSG00000138032.21 | PPM1B | 1930.90 | -0.42 | 0.11 | -3.66 | 0.00 | 0.03 |
| ENSG00000142546.14 | NOSIP | 1095.17 | 0.32 | 0.09 | 3.44 | 0.00 | 0.04 |
| ENSG00000058272.19 | PPP1R12A | 5974.89 | -0.48 | 0.13 | -3.62 | 0.00 | 0.03 |
| ENSG00000132356.12 | PRKAA1 | 2271.64 | -0.22 | 0.06 | -3.55 | 0.00 | 0.03 |
| ENSG00000175354.20 | PTPN2 | 1245.15 | -0.59 | 0.17 | -3.52 | 0.00 | 0.04 |
| ENSG00000166024.14 | R3HCC1L | 712.70 | -0.47 | 0.14 | -3.39 | 0.00 | 0.04 |
| ENSG00000206418.5 | RAB12 | 378.67 | -0.55 | 0.16 | -3.50 | 0.00 | 0.04 |
| ENSG00000119707.14 | RBM25 | 4138.43 | -0.33 | 0.06 | -5.11 | 0.00 | 0.00 |
| ENSG00000149823.9 | VPS51 | 1433.81 | 0.35 | 0.10 | 3.68 | 0.00 | 0.03 |
| ENSG00000134444.15 | RELCH | 2105.00 | -0.30 | 0.08 | -3.69 | 0.00 | 0.03 |
| ENSG00000185304.16 | RGPD2 | 519.39 | -2.46 | 0.66 | -3.73 | 0.00 | 0.02 |
| ENSG00000134318.15 | ROCK2 | 1291.21 | -0.56 | 0.13 | -4.35 | 0.00 | 0.01 |
| ENSG00000134321.13 | RSAD2 | 4589.55 | -2.35 | 0.61 | -3.83 | 0.00 | 0.02 |
| ENSG00000111011.18 | RSRC2 | 1442.29 | -0.40 | 0.10 | -3.99 | 0.00 | 0.02 |
| ENSG00000136514.3 | RTP4 | 537.22 | -1.66 | 0.39 | -4.22 | 0.00 | 0.01 |
| ENSG00000177409.12 | SAMD9L | 10945.52 | -1.25 | 0.29 | -4.28 | 0.00 | 0.01 |
| ENSG00000140386.13 | SCAPER | 788.08 | -0.69 | 0.17 | -4.02 | 0.00 | 0.02 |
| ENSG00000092108.22 | SCFD1 | 2007.47 | -0.39 | 0.11 | -3.41 | 0.00 | 0.04 |
| ENSG00000054282.16 | SDCCAG8 | 410.24 | -0.46 | 0.13 | -3.65 | 0.00 | 0.03 |
| ENSG00000108387.16 | SEPTIN4 | 464.91 | -2.79 | 0.70 | -3.96 | 0.00 | 0.02 |
| ENSG00000122545.21 | SEPTIN7 | 3533.11 | -0.34 | 0.10 | -3.39 | 0.00 | 0.04 |
| ENSG00000149131.17 | SERPING1 | 3225.83 | -1.69 | 0.49 | -3.47 | 0.00 | 0.04 |
| ENSG00000136603.14 | SKIL | 1411.24 | -0.57 | 0.16 | -3.46 | 0.00 | 0.04 |
| ENSG00000115904.15 | SOS1 | 3996.04 | -0.38 | 0.07 | -5.20 | 0.00 | 0.00 |
| ENSG00000079263.19 | SP140 | 1420.34 | -0.67 | 0.20 | -3.39 | 0.00 | 0.04 |
| ENSG00000196141.14 | SPATS2L | 378.10 | -1.92 | 0.46 | -4.20 | 0.00 | 0.01 |
| ENSG00000040341.18 | STAU2 | 873.16 | -0.34 | 0.10 | -3.48 | 0.00 | 0.04 |
| ENSG00000183735.11 | TBK1 | 1408.65 | -0.53 | 0.15 | -3.62 | 0.00 | 0.03 |
| ENSG00000148737.17 | TCF7L2 | 1027.63 | -0.61 | 0.15 | -3.95 | 0.00 | 0.02 |
| ENSG00000196116.8 | TDRD7 | 1044.10 | -0.76 | 0.22 | -3.48 | 0.00 | 0.04 |
| ENSG00000112773.16 | TENT5A | 1547.24 | -0.47 | 0.14 | -3.41 | 0.00 | 0.04 |
| ENSG00000105967.16 | TFEC | 1394.45 | -0.83 | 0.24 | -3.49 | 0.00 | 0.04 |
| ENSG00000169718.18 | DUS1L | 818.65 | 0.30 | 0.08 | 3.73 | 0.00 | 0.02 |
| ENSG00000114354.15 | TFG | 1571.02 | -0.46 | 0.11 | -4.09 | 0.00 | 0.02 |
| ENSG00000116001.17 | TIA1 | 1555.69 | -0.35 | 0.08 | -4.18 | 0.00 | 0.01 |
| ENSG00000078237.7 | TIGAR | 584.03 | -0.42 | 0.11 | -3.74 | 0.00 | 0.02 |
| ENSG00000164548.12 | TRA2A | 1819.53 | -0.41 | 0.12 | -3.40 | 0.00 | 0.04 |
| ENSG00000132274.16 | TRIM22 | 15160.28 | -0.95 | 0.27 | -3.46 | 0.00 | 0.04 |
| ENSG00000185880.13 | TRIM69 | 1903.63 | -0.76 | 0.20 | -3.76 | 0.00 | 0.02 |
| ENSG00000163728.11 | TTC14 | 2152.43 | -0.50 | 0.13 | -3.75 | 0.00 | 0.02 |
| ENSG00000159459.12 | UBR1 | 1484.56 | -0.39 | 0.10 | -3.80 | 0.00 | 0.02 |
| ENSG00000151461.20 | UPF2 | 1833.30 | -0.35 | 0.10 | -3.37 | 0.00 | 0.05 |
| ENSG00000155313.16 | USP25 | 2906.76 | -0.52 | 0.14 | -3.75 | 0.00 | 0.02 |
| ENSG00000138592.14 | USP8 | 2643.61 | -0.35 | 0.10 | -3.48 | 0.00 | 0.04 |
| ENSG00000168899.5 | VAMP5 | 365.40 | -1.20 | 0.33 | -3.65 | 0.00 | 0.03 |

| X-ray DEG set | | | | | | | |
|---|---|---|---|---|---|---|---|
| Ensembl ID | Gene name | baseMean | Log FC | Log FC SE | Wald statistic value | P-value | Adjusted p-value |
| ENSG00000085449.15 | WDFY1 | 2561.09 | -0.79 | 0.23 | -3.46 | 0.00 | 0.04 |
| ENSG00000132530.17 | XAF1 | 6074.95 | -1.54 | 0.36 | -4.24 | 0.00 | 0.01 |
| ENSG00000186908.15 | ZDHHC17 | 1747.57 | -0.55 | 0.15 | -3.71 | 0.00 | 0.02 |
| ENSG00000133858.17 | ZFC3H1 | 3519.82 | -0.34 | 0.10 | -3.46 | 0.00 | 0.04 |
| ENSG00000010244.19 | ZNF207 | 9781.28 | -0.31 | 0.08 | -3.84 | 0.00 | 0.02 |
| ENSG00000177853.15 | ZNF518A | 1629.41 | -0.55 | 0.16 | -3.44 | 0.00 | 0.04 |
| ENSG00000075292.19 | ZNF638 | 3429.56 | -0.39 | 0.10 | -3.94 | 0.00 | 0.02 |
| ENSG00000283378.1 | CNTNAP3C | 389.74 | 0.97 | 0.28 | 3.47 | 0.00 | 0.04 |

| SGRQ DEG set | | | | | | | |
|---|---|---|---|---|---|---|---|
| Gene ID | Gene name | baseMean | Log FC | Log FC SE | Wald statistic value | P-value | Adjusted p-value |
| ENSG00000108846.16 | ABCC3 | 719.10 | -0.75 | 0.21 | -3.49 | 0.00 | 0.05 |
| ENSG00000168615.13 | ADAM9 | 1019.38 | -0.57 | 0.16 | -3.61 | 0.00 | 0.04 |
| ENSG00000163568.15 | AIM2 | 583.81 | -1.25 | 0.35 | -3.60 | 0.00 | 0.04 |
| ENSG00000078053.17 | AMPH | 11.55 | -1.64 | 0.47 | -3.51 | 0.00 | 0.05 |
| ENSG00000011201.12 | ANOS1 | 34.18 | -2.30 | 0.48 | -4.77 | 0.00 | 0.00 |
| ENSG00000138772.13 | ANXA3 | 1503.05 | -1.43 | 0.40 | -3.59 | 0.00 | 0.04 |
| ENSG00000196975.16 | ANXA4 | 698.47 | -0.34 | 0.09 | -3.56 | 0.00 | 0.04 |
| ENSG00000100342.21 | APOL1 | 1888.98 | -1.01 | 0.29 | -3.53 | 0.00 | 0.05 |
| ENSG00000221963.6 | APOL6 | 17213.54 | -0.95 | 0.26 | -3.67 | 0.00 | 0.04 |
| ENSG00000146376.11 | ARHGAP18 | 749.84 | -0.59 | 0.17 | -3.55 | 0.00 | 0.04 |
| ENSG00000102401.20 | ARMCX3 | 501.54 | -0.31 | 0.09 | -3.50 | 0.00 | 0.05 |
| ENSG00000054793.14 | ATP9A | 410.14 | -0.98 | 0.28 | -3.56 | 0.00 | 0.04 |
| ENSG00000007516.14 | BAIAP3 | 147.60 | 1.12 | 0.29 | 3.83 | 0.00 | 0.03 |
| ENSG00000168062.10 | BATF2 | 1596.69 | -1.55 | 0.41 | -3.78 | 0.00 | 0.03 |
| ENSG00000198604.11 | BAZ1A | 5286.55 | -0.70 | 0.20 | -3.47 | 0.00 | 0.05 |
| ENSG00000166278.15 | C2 | 244.38 | -1.41 | 0.32 | -4.44 | 0.00 | 0.01 |
| ENSG00000182472.9 | CAPN12 | 86.14 | 1.12 | 0.27 | 4.10 | 0.00 | 0.02 |
| ENSG00000153113.24 | CAST | 5492.14 | -0.38 | 0.09 | -4.14 | 0.00 | 0.02 |
| ENSG00000269720.2 | CCDC194 | 33.84 | -1.32 | 0.35 | -3.82 | 0.00 | 0.03 |
| ENSG00000163823.4 | CCR1 | 5022.28 | -0.83 | 0.23 | -3.52 | 0.00 | 0.05 |
| ENSG00000121797.10 | CCRL2 | 171.74 | -1.04 | 0.29 | -3.57 | 0.00 | 0.04 |
| ENSG00000120217.14 | CD274 | 2167.85 | -1.74 | 0.42 | -4.11 | 0.00 | 0.02 |
| ENSG00000010278.15 | CD9 | 520.06 | -0.91 | 0.24 | -3.74 | 0.00 | 0.03 |
| ENSG00000163814.8 | CDCP1 | 90.61 | -1.57 | 0.44 | -3.55 | 0.00 | 0.04 |
| ENSG00000079385.23 | CEACAM1 | 2528.38 | -1.31 | 0.31 | -4.29 | 0.00 | 0.02 |
| ENSG00000170835.17 | CEL | 17.27 | -0.85 | 0.25 | -3.43 | 0.00 | 0.05 |
| ENSG00000143126.8 | CELSR2 | 131.83 | 0.64 | 0.18 | 3.58 | 0.00 | 0.04 |
| ENSG00000178404.10 | CEP295NL | 519.27 | -0.86 | 0.20 | -4.23 | 0.00 | 0.02 |
| ENSG00000231256.8 | CFAP97D1 | 12.27 | -2.03 | 0.51 | -3.98 | 0.00 | 0.02 |
| ENSG00000064886.14 | CHI3L2 | 98.23 | 0.83 | 0.20 | 4.12 | 0.00 | 0.02 |
| ENSG00000083937.10 | CHMP2B | 1137.85 | -0.60 | 0.17 | -3.45 | 0.00 | 0.05 |
| ENSG00000120903.13 | CHRNA2 | 24.43 | -1.60 | 0.43 | -3.69 | 0.00 | 0.04 |
| ENSG00000169105.8 | CHST14 | 183.28 | 0.30 | 0.09 | 3.49 | 0.00 | 0.05 |
| ENSG00000164237.9 | CMBL | 134.78 | -2.58 | 0.68 | -3.77 | 0.00 | 0.03 |
| ENSG00000130176.8 | CNN1 | 21.08 | -2.38 | 0.55 | -4.30 | 0.00 | 0.02 |
| ENSG00000044459.15 | CNTLN | 218.35 | -0.43 | 0.12 | -3.48 | 0.00 | 0.05 |
| ENSG00000283378.1 | CNTNAP3C | 389.74 | 0.91 | 0.26 | 3.49 | 0.00 | 0.05 |
| ENSG00000082438.18 | COBLL1 | 387.44 | 0.74 | 0.21 | 3.59 | 0.00 | 0.04 |
| ENSG00000188215.10 | DCUN1D3 | 530.95 | -0.56 | 0.16 | -3.47 | 0.00 | 0.05 |
| ENSG00000165490.13 | DDIAS | 211.75 | -1.13 | 0.28 | -4.02 | 0.00 | 0.02 |
| ENSG00000107201.11 | DDX58 | 7070.06 | -0.90 | 0.26 | -3.45 | 0.00 | 0.05 |
| ENSG00000181381.15 | DDX60L | 12494.93 | -0.96 | 0.24 | -4.06 | 0.00 | 0.02 |
| ENSG00000147202.19 | DIAPH2 | 1104.61 | -0.41 | 0.11 | -3.68 | 0.00 | 0.04 |
| ENSG00000162946.23 | DISC1 | 2992.55 | -0.81 | 0.19 | -4.21 | 0.00 | 0.02 |
| ENSG00000171962.18 | DRC3 | 6.52 | 1.73 | 0.46 | 3.79 | 0.00 | 0.03 |
| ENSG00000111817.19 | DSE | 2169.25 | -0.71 | 0.20 | -3.50 | 0.00 | 0.05 |
| ENSG00000169718.18 | DUS1L | 818.65 | 0.28 | 0.08 | 3.77 | 0.00 | 0.03 |
| ENSG00000132294.15 | EFR3A | 2312.33 | -0.36 | 0.10 | -3.72 | 0.00 | 0.03 |
| ENSG00000075151.24 | EIF4G3 | 3501.05 | -0.58 | 0.14 | -4.12 | 0.00 | 0.02 |
| ENSG00000168913.7 | ENHO | 9.29 | 1.88 | 0.36 | 5.16 | 0.00 | 0.00 |
| ENSG00000285708.1 | ENSG0000028571 | 114.58 | -0.83 | 0.24 | -3.43 | 0.00 | 0.05 |

| SGRQ DEG set | | | | | | | |
|---|---|---|---|---|---|---|---|
| Gene ID | Gene name | baseMean | Log FC | Log FC SE | Wald statistic value | P-value | Adjusted p-value |
| ENSG00000288681.1 | ENSG0000028868 | 29.23 | -6.10 | 1.68 | -3.63 | 0.00 | 0.04 |
| ENSG00000085832.17 | EPS15 | 3581.05 | -0.30 | 0.09 | -3.52 | 0.00 | 0.05 |
| ENSG00000104626.15 | ERI1 | 663.21 | -0.92 | 0.22 | -4.16 | 0.00 | 0.02 |
| ENSG00000166801.17 | FAM111A | 3387.33 | -0.44 | 0.13 | -3.46 | 0.00 | 0.05 |
| ENSG00000113391.19 | FAM172A | 807.11 | -0.33 | 0.09 | -3.60 | 0.00 | 0.04 |
| ENSG00000078098.15 | FAP | 8.35 | -2.15 | 0.42 | -5.11 | 0.00 | 0.00 |
| ENSG00000140092.15 | FBLN5 | 48.24 | 1.01 | 0.29 | 3.46 | 0.00 | 0.05 |
| ENSG00000143756.12 | FBXO28 | 952.55 | -0.27 | 0.08 | -3.47 | 0.00 | 0.05 |
| ENSG00000112367.12 | FIG4 | 2590.29 | -0.49 | 0.13 | -3.93 | 0.00 | 0.02 |
| ENSG00000261308.4 | FIGNL2 | 40.93 | 0.82 | 0.18 | 4.47 | 0.00 | 0.01 |
| ENSG00000119686.10 | FLVCR2 | 561.99 | -0.97 | 0.27 | -3.66 | 0.00 | 0.04 |
| ENSG00000186765.12 | FSCN2 | 11.61 | 1.18 | 0.29 | 4.05 | 0.00 | 0.02 |
| ENSG00000082074.19 | FYB1 | 34291.87 | -0.57 | 0.16 | -3.50 | 0.00 | 0.05 |
| ENSG00000099860.9 | GADD45B | 1745.08 | -0.78 | 0.22 | -3.58 | 0.00 | 0.04 |
| ENSG00000054983.17 | GALC | 1226.98 | -0.41 | 0.11 | -3.79 | 0.00 | 0.03 |
| ENSG00000143891.17 | GALM | 507.14 | -1.25 | 0.35 | -3.55 | 0.00 | 0.04 |
| ENSG00000141013.17 | GAS8 | 84.03 | -0.73 | 0.18 | -4.14 | 0.00 | 0.02 |
| ENSG00000117228.11 | GBP1 | 11410.44 | -1.24 | 0.35 | -3.50 | 0.00 | 0.05 |
| ENSG00000162654.9 | GBP4 | 4790.51 | -0.96 | 0.23 | -4.17 | 0.00 | 0.02 |
| ENSG00000154451.15 | GBP5 | 35542.51 | -1.30 | 0.35 | -3.75 | 0.00 | 0.03 |
| ENSG00000183347.15 | GBP6 | 290.04 | -1.74 | 0.43 | -4.01 | 0.00 | 0.02 |
| ENSG00000131979.20 | GCH1 | 1306.08 | -0.93 | 0.25 | -3.74 | 0.00 | 0.03 |
| ENSG00000066455.13 | GOLGA5 | 638.99 | -0.30 | 0.08 | -3.85 | 0.00 | 0.03 |
| ENSG00000139572.4 | GPR84 | 153.58 | -1.95 | 0.47 | -4.12 | 0.00 | 0.02 |
| ENSG00000198785.7 | GRIN3A | 50.11 | -1.42 | 0.37 | -3.80 | 0.00 | 0.03 |
| ENSG00000163754.18 | GYG1 | 1832.58 | -1.12 | 0.32 | -3.46 | 0.00 | 0.05 |
| ENSG00000187475.6 | H1-6 | 54.60 | -1.04 | 0.25 | -4.13 | 0.00 | 0.02 |
| ENSG00000103253.19 | HAGHL | 40.69 | 1.01 | 0.22 | 4.57 | 0.00 | 0.01 |
| ENSG00000244734.4 | HBB | 2730.16 | 1.38 | 0.40 | 3.49 | 0.00 | 0.05 |
| ENSG00000090339.9 | ICAM1 | 2178.09 | -0.64 | 0.18 | -3.52 | 0.00 | 0.05 |
| ENSG00000163565.20 | IFI16 | 10121.14 | -0.76 | 0.19 | -4.02 | 0.00 | 0.02 |
| ENSG00000068079.8 | IFI35 | 1232.53 | -1.05 | 0.26 | -4.08 | 0.00 | 0.02 |
| ENSG00000115267.9 | IFIH1 | 2409.91 | -0.98 | 0.27 | -3.68 | 0.00 | 0.04 |
| ENSG00000282122.1 | IGHV7-4-1 | 6.82 | -5.48 | 1.14 | -4.80 | 0.00 | 0.00 |
| ENSG00000136634.7 | IL10 | 18.70 | -1.57 | 0.36 | -4.33 | 0.00 | 0.02 |
| ENSG00000197272.2 | IL27 | 34.80 | -1.55 | 0.40 | -3.92 | 0.00 | 0.02 |
| ENSG00000148384.14 | INPP5E | 267.37 | 0.47 | 0.14 | 3.44 | 0.00 | 0.05 |
| ENSG00000117595.12 | IRF6 | 15.64 | 1.27 | 0.37 | 3.49 | 0.00 | 0.05 |
| ENSG00000096968.14 | JAK2 | 5541.44 | -0.73 | 0.21 | -3.44 | 0.00 | 0.05 |
| ENSG00000176076.7 | KCNE5 | 51.54 | -1.07 | 0.30 | -3.61 | 0.00 | 0.04 |
| ENSG00000102554.14 | KLF5 | 287.55 | -0.76 | 0.20 | -3.81 | 0.00 | 0.03 |
| ENSG00000185271.9 | KLHL33 | 13.07 | 1.40 | 0.35 | 3.98 | 0.00 | 0.02 |
| ENSG00000189182.10 | KRT77 | 24.30 | 9.36 | 2.58 | 3.63 | 0.00 | 0.04 |
| ENSG00000078081.8 | LAMP3 | 106.99 | -1.58 | 0.40 | -3.91 | 0.00 | 0.02 |
| ENSG00000002549.13 | LAP3 | 2399.82 | -1.44 | 0.34 | -4.22 | 0.00 | 0.02 |
| ENSG00000116977.19 | LGALS8 | 3086.67 | -0.47 | 0.12 | -3.98 | 0.00 | 0.02 |
| ENSG00000102897.10 | LYRM1 | 553.83 | -0.64 | 0.18 | -3.49 | 0.00 | 0.05 |
| ENSG00000108984.15 | MAP2K6 | 2172.90 | -0.76 | 0.21 | -3.58 | 0.00 | 0.04 |
| ENSG00000168175.15 | MAPK1IP1L | 2329.39 | -0.21 | 0.06 | -3.74 | 0.00 | 0.03 |
| ENSG00000183019.8 | MCEMP1 | 770.73 | -1.62 | 0.44 | -3.68 | 0.00 | 0.04 |
| ENSG00000158411.11 | MITD1 | 687.71 | -0.41 | 0.12 | -3.50 | 0.00 | 0.05 |

| SGRQ DEG set | | | | | | | |
|---|---|---|---|---|---|---|---|
| Gene ID | Gene name | baseMean | Log FC | Log FC SE | Wald statistic value | P-value | Adjusted p-value |
| ENSG00000184350.11 | MRGPRE | 8.59 | -3.39 | 0.98 | -3.46 | 0.00 | 0.05 |
| ENSG00000110079.19 | MS4A4A | 213.04 | -0.99 | 0.26 | -3.86 | 0.00 | 0.03 |
| ENSG00000101335.10 | MYL9 | 1852.83 | -1.45 | 0.41 | -3.54 | 0.00 | 0.05 |
| ENSG00000145555.15 | MYO10 | 124.65 | -1.55 | 0.36 | -4.26 | 0.00 | 0.02 |
| ENSG00000036448.10 | MYOM2 | 154.37 | 2.65 | 0.65 | 4.09 | 0.00 | 0.02 |
| ENSG00000145911.6 | N4BP3 | 79.88 | 1.11 | 0.27 | 4.06 | 0.00 | 0.02 |
| ENSG00000130202.10 | NECTIN2 | 522.20 | -1.62 | 0.45 | -3.62 | 0.00 | 0.04 |
| ENSG00000072736.19 | NFATC3 | 4644.62 | 0.29 | 0.08 | 3.55 | 0.00 | 0.04 |
| ENSG00000086288.12 | NME8 | 74.98 | -0.81 | 0.18 | -4.61 | 0.00 | 0.01 |
| ENSG00000188000.6 | OR7D2 | 4.88 | 4.50 | 1.12 | 4.01 | 0.00 | 0.02 |
| ENSG00000197646.8 | PDCD1LG2 | 201.03 | -2.32 | 0.59 | -3.93 | 0.00 | 0.02 |
| ENSG00000163110.15 | PDLIM5 | 1587.10 | -0.33 | 0.09 | -3.56 | 0.00 | 0.04 |
| ENSG00000102893.16 | PHKB | 2051.37 | -0.31 | 0.07 | -4.23 | 0.00 | 0.02 |
| ENSG00000155629.15 | PIK3AP1 | 11008.59 | -0.90 | 0.24 | -3.77 | 0.00 | 0.03 |
| ENSG00000188313.13 | PLSCR1 | 3809.84 | -1.21 | 0.34 | -3.60 | 0.00 | 0.04 |
| ENSG00000143224.18 | PPOX | 226.62 | 0.62 | 0.16 | 3.91 | 0.00 | 0.02 |
| ENSG00000138738.11 | PRDM5 | 256.21 | -1.01 | 0.28 | -3.60 | 0.00 | 0.04 |
| ENSG00000183248.12 | PRR36 | 4.15 | 2.13 | 0.62 | 3.46 | 0.00 | 0.05 |
| ENSG00000106772.19 | PRUNE2 | 75.64 | -2.14 | 0.39 | -5.42 | 0.00 | 0.00 |
| ENSG00000152229.18 | PSTPIP2 | 3277.85 | -1.17 | 0.28 | -4.11 | 0.00 | 0.02 |
| ENSG00000127947.16 | PTPN12 | 3934.41 | -0.52 | 0.15 | -3.49 | 0.00 | 0.05 |
| ENSG00000110060.9 | PUS3 | 95.40 | -0.58 | 0.16 | -3.63 | 0.00 | 0.04 |
| ENSG00000165661.17 | QSOX2 | 402.36 | 0.43 | 0.11 | 3.99 | 0.00 | 0.02 |
| ENSG00000139832.5 | RAB20 | 381.37 | -0.97 | 0.19 | -5.07 | 0.00 | 0.00 |
| ENSG00000075391.17 | RASAL2 | 28.06 | 0.72 | 0.19 | 3.73 | 0.00 | 0.03 |
| ENSG00000099849.15 | RASSF7 | 196.84 | 0.50 | 0.14 | 3.50 | 0.00 | 0.05 |
| ENSG00000072042.13 | RDH11 | 855.45 | -0.30 | 0.08 | -3.62 | 0.00 | 0.04 |
| ENSG00000099999.15 | RNF215 | 99.36 | 0.38 | 0.11 | 3.50 | 0.00 | 0.05 |
| ENSG00000154134.15 | ROBO3 | 73.12 | 1.12 | 0.31 | 3.63 | 0.00 | 0.04 |
| ENSG00000197958.13 | RPL12 | 2576.47 | 0.47 | 0.12 | 4.05 | 0.00 | 0.02 |
| ENSG00000167526.14 | RPL13 | 8488.27 | 0.50 | 0.13 | 3.77 | 0.00 | 0.03 |
| ENSG00000142541.18 | RPL13A | 7915.85 | 0.59 | 0.17 | 3.44 | 0.00 | 0.05 |
| ENSG00000100316.16 | RPL3 | 8744.45 | 0.66 | 0.18 | 3.61 | 0.00 | 0.04 |
| ENSG00000137154.13 | RPS6 | 2812.29 | 0.63 | 0.18 | 3.43 | 0.00 | 0.05 |
| ENSG00000081019.14 | RSBN1 | 1955.42 | -0.51 | 0.15 | -3.49 | 0.00 | 0.05 |
| ENSG00000108309.14 | RUNDC3A | 1383.89 | 1.84 | 0.53 | 3.47 | 0.00 | 0.05 |
| ENSG00000177409.12 | SAMD9L | 10945.52 | -0.99 | 0.28 | -3.52 | 0.00 | 0.05 |
| ENSG00000284194.3 | SCO2 | 659.50 | -1.00 | 0.28 | -3.52 | 0.00 | 0.05 |
| ENSG00000162512.16 | SDC3 | 332.16 | -0.86 | 0.24 | -3.62 | 0.00 | 0.04 |
| ENSG00000143434.17 | SEMA6C | 46.46 | 0.72 | 0.17 | 4.13 | 0.00 | 0.02 |
| ENSG00000184990.13 | SIVA1 | 354.01 | 0.42 | 0.10 | 4.02 | 0.00 | 0.02 |
| ENSG00000108932.13 | SLC16A6 | 823.43 | -0.47 | 0.12 | -3.99 | 0.00 | 0.02 |
| ENSG00000143036.17 | SLC44A3 | 8.83 | -1.53 | 0.43 | -3.51 | 0.00 | 0.05 |
| ENSG00000114923.17 | SLC4A3 | 11.10 | 1.26 | 0.37 | 3.45 | 0.00 | 0.05 |
| ENSG00000111181.13 | SLC6A12 | 164.83 | -0.74 | 0.19 | -3.89 | 0.00 | 0.02 |
| ENSG00000174358.16 | SLC6A19 | 146.19 | 3.41 | 0.94 | 3.62 | 0.00 | 0.04 |
| ENSG00000066230.12 | SLC9A3 | 79.56 | 0.77 | 0.22 | 3.44 | 0.00 | 0.05 |
| ENSG00000082014.17 | SMARCD3 | 1089.13 | -0.88 | 0.26 | -3.46 | 0.00 | 0.05 |
| ENSG00000115904.15 | SOS1 | 3996.04 | -0.28 | 0.08 | -3.46 | 0.00 | 0.05 |
| ENSG00000067066.17 | SP100 | 9275.37 | -0.59 | 0.17 | -3.45 | 0.00 | 0.05 |
| ENSG00000186583.12 | SPATC1 | 44.07 | -1.58 | 0.36 | -4.35 | 0.00 | 0.02 |

| SGRQ DEG set | | | | | | | |
|---|---|---|---|---|---|---|---|
| Gene ID | Gene name | baseMean | Log FC | Log FC SE | Wald statistic value | P-value | Adjusted p-value |
| ENSG00000196141.14 | SPATS2L | 378.10 | -1.60 | 0.42 | -3.79 | 0.00 | 0.03 |
| ENSG00000198917.13 | SPOUT1 | 402.19 | 0.32 | 0.09 | 3.70 | 0.00 | 0.04 |
| ENSG00000005206.17 | SPPL2B | 661.33 | 0.38 | 0.09 | 4.07 | 0.00 | 0.02 |
| ENSG00000100883.13 | SRP54 | 1040.48 | -0.26 | 0.07 | -3.48 | 0.00 | 0.05 |
| ENSG00000104375.17 | STK3 | 555.21 | -0.90 | 0.22 | -4.11 | 0.00 | 0.02 |
| ENSG00000204634.13 | TBC1D8 | 1065.16 | -0.90 | 0.22 | -4.17 | 0.00 | 0.02 |
| ENSG00000148737.17 | TCF7L2 | 1027.63 | -0.53 | 0.14 | -3.67 | 0.00 | 0.04 |
| ENSG00000185339.9 | TCN2 | 246.19 | -0.99 | 0.26 | -3.79 | 0.00 | 0.03 |
| ENSG00000204852.17 | TCTN1 | 207.43 | 0.72 | 0.18 | 3.96 | 0.00 | 0.02 |
| ENSG00000156414.20 | TDRD9 | 372.24 | -1.53 | 0.37 | -4.15 | 0.00 | 0.02 |
| ENSG00000198723.11 | TEX45 | 6.27 | 2.19 | 0.57 | 3.88 | 0.00 | 0.03 |
| ENSG00000114354.15 | TFG | 1571.02 | -0.39 | 0.11 | -3.48 | 0.00 | 0.05 |
| ENSG00000145365.11 | TIFA | 1060.11 | -1.24 | 0.33 | -3.77 | 0.00 | 0.03 |
| ENSG00000078237.7 | TIGAR | 584.03 | -0.38 | 0.10 | -3.70 | 0.00 | 0.04 |
| ENSG00000187554.14 | TLR5 | 2839.51 | -1.10 | 0.29 | -3.76 | 0.00 | 0.03 |
| ENSG00000146072.6 | TNFRSF21 | 39.33 | 0.86 | 0.21 | 4.10 | 0.00 | 0.02 |
| ENSG00000143549.21 | TPM3 | 13460.28 | -0.36 | 0.10 | -3.57 | 0.00 | 0.04 |
| ENSG00000188056.12 | TREML4 | 65.44 | -6.87 | 1.37 | -5.01 | 0.00 | 0.00 |
| ENSG00000132274.16 | TRIM22 | 15160.28 | -0.86 | 0.25 | -3.43 | 0.00 | 0.05 |
| ENSG00000185880.13 | TRIM69 | 1903.63 | -0.66 | 0.19 | -3.53 | 0.00 | 0.05 |
| ENSG00000159459.12 | UBR1 | 1484.56 | -0.35 | 0.09 | -3.69 | 0.00 | 0.04 |
| ENSG00000049245.13 | VAMP3 | 2038.43 | -0.66 | 0.18 | -3.74 | 0.00 | 0.03 |
| ENSG00000168899.5 | VAMP5 | 365.40 | -1.10 | 0.30 | -3.72 | 0.00 | 0.03 |
| ENSG00000149823.9 | VPS51 | 1433.81 | 0.33 | 0.09 | 3.71 | 0.00 | 0.03 |
| ENSG00000176871.9 | WSB2 | 1503.67 | -0.69 | 0.18 | -3.87 | 0.00 | 0.03 |
| ENSG00000114127.11 | XRN1 | 8154.76 | -0.70 | 0.20 | -3.58 | 0.00 | 0.04 |
| ENSG00000183621.15 | ZNF438 | 1557.73 | -0.87 | 0.25 | -3.52 | 0.00 | 0.05 |

| Gene name | Log FC SGRQ outcome M6 | Log FC X-ray outome M6 | Reactome - Immune System | Reactome - Interferon gamma signaling |
|---|---|---|---|---|
| GBP5 | -1.30 | -1.33 | ✓ | ✓ |
| GBP4 | -0.96 | -0.93 | ✓ | ✓ |
| TRIM22 | -0.86 | -0.95 | ✓ | ✓ |
| JAK2 | -0.73 | -0.86 | ✓ | ✓ |
| CD274 | -1.74 | -1.71 | ✓ | ✗ |
| IFIH1 | -0.98 | -1.13 | ✓ | ✗ |
| DDX58 | -0.90 | -1.03 | ✓ | ✗ |
| TRIM69 | -0.66 | -0.76 | ✓ | ✗ |
| UBR1 | -0.35 | -0.39 | ✓ | ✗ |
| SOS1 | -0.28 | -0.38 | ✓ | ✗ |
| NFATC3 | 0.29 | 0.33 | ✓ | ✗ |
| SPATS2L | -1.60 | -1.92 | ✗ | ✗ |
| LAP3 | -1.44 | -1.43 | ✗ | ✗ |
| VAMP5 | -1.10 | -1.20 | ✗ | ✗ |
| SAMD9L | -0.99 | -1.25 | ✗ | ✗ |
| TCF7L2 | -0.53 | -0.61 | ✗ | ✗ |
| FIG4 | -0.49 | -0.50 | ✗ | ✗ |
| FAM111A | -0.44 | -0.52 | ✗ | ✗ |
| MITD1 | -0.41 | -0.47 | ✗ | ✗ |
| DIAPH2 | -0.41 | -0.46 | ✗ | ✗ |
| TFG | -0.39 | -0.46 | ✗ | ✗ |
| TIGAR | -0.38 | -0.42 | ✗ | ✗ |
| EFR3A | -0.36 | -0.40 | ✗ | ✗ |
| ANXA4 | -0.34 | -0.37 | ✗ | ✗ |
| FAM172A | -0.33 | -0.36 | ✗ | ✗ |
| PHKB | -0.31 | -0.30 | ✗ | ✗ |
| FBXO28 | -0.27 | -0.30 | ✗ | ✗ |
| DUS1L | 0.28 | 0.30 | ✗ | ✗ |
| VPS51 | 0.33 | 0.35 | ✗ | ✗ |
| CNTNAP3C | 0.91 | 0.97 | ✗ | ✗ |

| Biomarker | Heycken_FAIL_22 | Thompson_FAIL_13 | Long_RES_10 | Tabone_RES_25 | Tabone_RES_27 | Thompson_RES_5 | Zimmer_RES_3 | SGRQ DEG set | X-ray DEG set | SCC DEG set | Sum -Public signatures | Sum- Public sign + DEG Sets |
|---|---|---|---|---|---|---|---|---|---|---|---|---|
| CD274 | ✓ | | ✓ | ✓ | ✓ | | | ✓ | ✓ | | 4 | 6 |
| GBP5 | ✓ | | | | ✓ | | ✓ | ✓ | ✓ | | 3 | 5 |
| CASP5 | | | | ✓ | ✓ | | | | | | 2 | 2 |
| DUSP3 | | | | ✓ | | | ✓ | | | | 2 | 2 |
| FAM20A | ✓ | | | | ✓ | | | | | | 2 | 2 |
| FCGR1A | | | ✓ | | ✓ | | | | | | 2 | 2 |
| IFITM1 | ✓ | | | ✓ | | | | | | | 2 | 2 |
| MAP7D3 | | ✓ | | | | ✓ | | | | | 2 | 2 |
| PDCD1LG2 | | | | ✓ | ✓ | | | ✓ | | | 2 | 3 |
| SEPT4 | | | | ✓ | ✓ | | | | | | 2 | 2 |
| STT3A | | ✓ | | | | ✓ | | | | | 2 | 2 |
| TIFA | | | | ✓ | ✓ | | | ✓ | | | 2 | 3 |
| UCP2 | | ✓ | | | | ✓ | | | | | 2 | 2 |
| VAMP5 | ✓ | | | ✓ | | | | ✓ | ✓ | | 2 | 4 |
| A_33_P3281041 | ✓ | | | | | | | | | | 1 | 1 |
| ANXA3 | | | | ✓ | | | | ✓ | | | 1 | 2 |
| APOL1 | | | | ✓ | | | | ✓ | | | 1 | 2 |
| APOL6 | | | | ✓ | | | | ✓ | | | 1 | 2 |
| ATP1A1 | | ✓ | | | | | | | | | 1 | 1 |
| BAK1 | | | | | ✓ | | | | | | 1 | 1 |
| BATF2 | ✓ | | | | | | | ✓ | | | 1 | 2 |
| C1QB | | | | | ✓ | | | | | | 1 | 1 |
| C1QC | | | | | ✓ | | | | | | 1 | 1 |
| C2 | ✓ | | | | | | | ✓ | | | 1 | 2 |
| CDCP1 | | | | | ✓ | | | ✓ | | | 1 | 2 |
| CXorf40B | | ✓ | | | | | | | | | 1 | 1 |
| DARS2 | | ✓ | | | | | | | | | 1 | 1 |
| EFCAB2 | | | | ✓ | | | | | | | 1 | 1 |
| EGF | | | ✓ | | | | | | | | 1 | 1 |
| EMC1 | | ✓ | | | | | | | | | 1 | 1 |
| EXOC2 | | ✓ | | | | | | | | | 1 | 1 |
| FCER1G | | | | ✓ | | | | | | | 1 | 1 |
| FCGR1B | | | | | ✓ | | | | | | 1 | 1 |
| FCGR1CP | | | | | ✓ | | | | | | 1 | 1 |
| GBP1 | | | ✓ | | | | | ✓ | | | 1 | 2 |
| GBP1P1 | | | | | ✓ | | | | | | 1 | 1 |
| GBP2 | | | ✓ | | | | | | | | 1 | 1 |
| GBP6 | | | | | ✓ | | | ✓ | | | 1 | 2 |
| GK | ✓ | | | | | | | | | | 1 | 1 |
| GPR84 | | | | ✓ | | | | ✓ | | | 1 | 2 |
| GRAMD1C | | | | | ✓ | | | | | | 1 | 1 |
| GRIN3A | | | | | ✓ | | | ✓ | | | 1 | 2 |
| GYG1 | ✓ | | | | | | | ✓ | | | 1 | 2 |
| HIST1H1B | ✓ | | | | | | | | | | 1 | 1 |
| ICAM1 | | | | | ✓ | | | ✓ | | | 1 | 2 |
| IFIT2 | ✓ | | | | | | | | | | 1 | 1 |
| IGF2BP3 | | | | | ✓ | | | | | | 1 | 1 |
| IL15 | | | ✓ | | | | | | | | 1 | 1 |
| IL27 | ✓ | | | | | | | ✓ | | | 1 | 2 |
| IRF1 | | | | ✓ | | | | | | | 1 | 1 |
| KCNJ2-AS1 | ✓ | | | | | | | | | | 1 | 1 |
| KIAA0100 | | ✓ | | | | | | | | ✓ | 1 | 2 |
| KIF1B | | | ✓ | | | | | | | | 1 | 1 |
| KLF2 | | | | | | | ✓ | | | | 1 | 1 |
| KREMEN1 | ✓ | | | | | | | | | | 1 | 1 |
| LIMK1 | | | | | ✓ | | | | | | 1 | 1 |
| LPCAT2 | ✓ | | | | | | | | | | 1 | 1 |
| NOD2 | | | ✓ | | | | | | | | 1 | 1 |
| NRN1 | | | | | ✓ | | | | | | 1 | 1 |
| P2RY14 | | | | | ✓ | | | | | | 1 | 1 |
| PDE4D | ✓ | | | | | | | | | | 1 | 1 |
| PML | | | | ✓ | | | | | | | 1 | 1 |
| POLB | | | | ✓ | | | | | | | 1 | 1 |
| PRTN3 | | | | | ✓ | | | | | | 1 | 1 |
| PSMD1 | | ✓ | | | | | | | | | 1 | 1 |
| PSME2 | | | | ✓ | | | | | | | 1 | 1 |
| RARRES3 | | | | ✓ | | | | | | | 1 | 1 |
| RP11-295G20.2 | | | | | | ✓ | | | | | 1 | 1 |
| RP11-468E2.5 | | | | ✓ | | | | | | | 1 | 1 |

| Biomarker | Heycken_FAIL_22 | Thompson_FAIL_13 | Long_RES_10 | Tabone_RES_25 | Tabone_RES_27 | Thompson_RES_5 | Zimmer_RES_3 | SGRQ DEG set | X-ray DEG set | SCC DEG set | Sum -Public signatures | Sum- Public sign + DEG Sets |
|---|---|---|---|---|---|---|---|---|---|---|---|---|
| RP11-476010.1 | | | | ✓ | | | | | | | 1 | 1 |
| RPAP3 | ✓ | | | | | | | | | | 1 | 1 |
| SCARF1 | | | | | ✓ | | | | | | 1 | 1 |
| SDC3 | | | | | ✓ | | | ✓ | | | 1 | 2 |
| SEC31A | | ✓ | | | | | | | | | 1 | 1 |
| SERPING | ✓ | | | | | | | | | | 1 | 1 |
| SMARCD3 | | | | | | ✓ | | ✓ | | | 1 | 2 |
| SND1 | | ✓ | | | | | | | | | 1 | 1 |
| SOCS1 | | | | ✓ | | | | | | | 1 | 1 |
| SOCS3 | | | | | ✓ | | | | | | 1 | 1 |
| SORT1 | | | | ✓ | | | | | | | 1 | 1 |
| STAT1 | ✓ | | | | | | | | | | 1 | 1 |
| TIMM10 | | | | ✓ | | | | | | | 1 | 1 |
| TLR1 | | | ✓ | | | | | | | | 1 | 1 |
| TLR5 | | | ✓ | | | | | ✓ | | | 1 | 2 |
| TNFRSF21 | ✓ | | | | | | | ✓ | | | 1 | 2 |
| TRIM27 | ✓ | | | | | | | | | | 1 | 1 |
| UBE2L6 | | | | ✓ | | | | | | | 1 | 1 |
| VWA8 | | ✓ | | | | | | | | | 1 | 1 |

| Variable Name (diagonal) | Variable Name (vertical) | Spearman's rank correlation | Adjusted p-value |
|---|---|---|---|
| IFNG | IL-10 | 0.16 | 0.24 |
| IFNG | IL-12 | 0.00 | 1.00 |
| IFNG | IL-17A | -0.15 | 0.28 |
| IFNG | IL-1B | 0.32 | 0.01 |
| IFNG | IL-2 | -0.44 | 0.00 |
| IFNG | IL-4 | 0.56 | 0.00 |
| IFNG | IL-6 | 0.05 | 0.73 |
| IFNG | IL-8 | 0.21 | 0.13 |
| IFNG | TNF | -0.19 | 0.16 |
| IFNG | TBS score | 0.03 | 0.85 |
| IFNG | SGRQ score | -0.02 | 0.89 |
| IFNG | X-ray score | 0.32 | 0.05 |
| IFNG | Hb | -0.20 | 0.14 |
| IFNG | ESR | 0.15 | 0.29 |
| IFNG | NLR | 0.42 | 0.00 |
| IFNG | MLR | 0.20 | 0.15 |
| IFNG | SII | 0.41 | 0.00 |
| IFNG | Heycken_FAIL_22 | 0.46 | 0.00 |
| IFNG | Long_RES_10 | 0.45 | 0.00 |
| IFNG | Tabone_RES_25 | 0.43 | 0.00 |
| IFNG | Tabone_RES_27 | 0.45 | 0.00 |
| IFNG | Thompson_FAIL_13 | -0.45 | 0.00 |
| IFNG | Thompson_RES_5 | 0.27 | 0.04 |
| IFNG | Zimmer_RES_3 | 0.44 | 0.00 |
| IFNG | SCC DEG set | -0.29 | 0.02 |
| IFNG | X-ray M6 DEG set | 0.17 | 0.23 |
| IFNG | SGRQ M6 DEG set | 0.44 | 0.00 |
| IL-10 | IL-12 | 0.83 | 0.00 |
| IL-10 | IL-17A | 0.84 | 0.00 |
| IL-10 | IL-1B | 0.87 | 0.00 |
| IL-10 | IL-2 | 0.65 | 0.00 |
| IL-10 | IL-4 | 0.48 | 0.00 |
| IL-10 | IL-6 | 0.36 | 0.01 |
| IL-10 | IL-8 | 0.29 | 0.02 |
| IL-10 | TNF | 0.60 | 0.00 |
| IL-10 | TBS score | 0.22 | 0.11 |
| IL-10 | SGRQ score | 0.18 | 0.18 |
| IL-10 | X-ray score | 0.05 | 0.79 |
| IL-10 | Hb | -0.13 | 0.36 |
| IL-10 | ESR | -0.04 | 0.79 |
| IL-10 | NLR | 0.12 | 0.39 |
| IL-10 | MLR | -0.16 | 0.25 |
| IL-10 | SII | 0.20 | 0.14 |
| IL-10 | Heycken_FAIL_22 | 0.31 | 0.02 |
| IL-10 | Long_RES_10 | 0.31 | 0.02 |
| IL-10 | Tabone_RES_25 | 0.29 | 0.03 |
| IL-10 | Tabone_RES_27 | 0.30 | 0.02 |
| IL-10 | Thompson_FAIL_13 | -0.19 | 0.16 |

| Variable Name (diagonal) | Variable Name (vertical) | Spearman's rank correlation | Adjusted p-value |
|---|---|---|---|
| IL-10 | Thompson_RES_5 | 0.34 | 0.01 |
| IL-10 | Zimmer_RES_3 | 0.30 | 0.02 |
| IL-10 | SCC DEG set | -0.04 | 0.81 |
| IL-10 | X-ray M6 DEG set | 0.18 | 0.19 |
| IL-10 | SGRQ M6 DEG set | 0.29 | 0.02 |
| IL-12 | IL-17A | 0.90 | 0.00 |
| IL-12 | IL-1B | 0.79 | 0.00 |
| IL-12 | IL-2 | 0.80 | 0.00 |
| IL-12 | IL-4 | 0.34 | 0.01 |
| IL-12 | IL-6 | 0.27 | 0.04 |
| IL-12 | IL-8 | 0.20 | 0.15 |
| IL-12 | TNF | 0.56 | 0.00 |
| IL-12 | TBS score | 0.01 | 0.94 |
| IL-12 | SGRQ score | 0.04 | 0.79 |
| IL-12 | X-ray score | -0.10 | 0.55 |
| IL-12 | Hb | 0.09 | 0.53 |
| IL-12 | ESR | -0.21 | 0.14 |
| IL-12 | NLR | 0.08 | 0.59 |
| IL-12 | MLR | -0.16 | 0.26 |
| IL-12 | SII | 0.14 | 0.34 |
| IL-12 | Heycken_FAIL_22 | 0.13 | 0.33 |
| IL-12 | Long_RES_10 | 0.14 | 0.32 |
| IL-12 | Tabone_RES_25 | 0.12 | 0.39 |
| IL-12 | Tabone_RES_27 | 0.15 | 0.27 |
| IL-12 | Thompson_FAIL_13 | 0.03 | 0.84 |
| IL-12 | Thompson_RES_5 | 0.26 | 0.05 |
| IL-12 | Zimmer_RES_3 | 0.18 | 0.18 |
| IL-12 | SCC DEG set | 0.10 | 0.46 |
| IL-12 | X-ray M6 DEG set | 0.12 | 0.40 |
| IL-12 | SGRQ M6 DEG set | 0.12 | 0.40 |
| IL-17A | IL-1B | 0.78 | 0.00 |
| IL-17A | IL-2 | 0.86 | 0.00 |
| IL-17A | IL-4 | 0.32 | 0.01 |
| IL-17A | IL-6 | 0.38 | 0.00 |
| IL-17A | IL-8 | 0.32 | 0.01 |
| IL-17A | TNF | 0.69 | 0.00 |
| IL-17A | TBS score | 0.13 | 0.37 |
| IL-17A | SGRQ score | 0.18 | 0.18 |
| IL-17A | X-ray score | -0.10 | 0.57 |
| IL-17A | Hb | -0.05 | 0.76 |
| IL-17A | ESR | -0.12 | 0.38 |
| IL-17A | NLR | 0.04 | 0.77 |
| IL-17A | MLR | -0.21 | 0.13 |
| IL-17A | SII | 0.09 | 0.52 |
| IL-17A | Heycken_FAIL_22 | 0.12 | 0.40 |
| IL-17A | Long_RES_10 | 0.14 | 0.30 |
| IL-17A | Tabone_RES_25 | 0.13 | 0.36 |
| IL-17A | Tabone_RES_27 | 0.14 | 0.30 |

| Variable Name (diagonal) | Variable Name (vertical) | Spearman's rank correlation | Adjusted p-value |
|---|---|---|---|
| IL-17A | Thompson_FAIL_13 | 0.00 | 0.98 |
| IL-17A | Thompson_RES_5 | 0.27 | 0.04 |
| IL-17A | Zimmer_RES_3 | 0.17 | 0.23 |
| IL-17A | SCC DEG set | 0.02 | 0.91 |
| IL-17A | X-ray M6 DEG set | 0.09 | 0.53 |
| IL-17A | SGRQ M6 DEG set | 0.13 | 0.34 |
| IL-1B | IL-2 | 0.57 | 0.00 |
| IL-1B | IL-4 | 0.65 | 0.00 |
| IL-1B | IL-6 | 0.46 | 0.00 |
| IL-1B | IL-8 | 0.41 | 0.00 |
| IL-1B | TNF | 0.60 | 0.00 |
| IL-1B | TBS score | 0.13 | 0.38 |
| IL-1B | SGRQ score | 0.09 | 0.51 |
| IL-1B | X-ray score | 0.00 | 0.99 |
| IL-1B | Hb | -0.18 | 0.18 |
| IL-1B | ESR | 0.00 | 1.00 |
| IL-1B | NLR | 0.22 | 0.11 |
| IL-1B | MLR | -0.04 | 0.80 |
| IL-1B | SII | 0.23 | 0.09 |
| IL-1B | Heycken_FAIL_22 | 0.30 | 0.02 |
| IL-1B | Long_RES_10 | 0.32 | 0.01 |
| IL-1B | Tabone_RES_25 | 0.29 | 0.02 |
| IL-1B | Tabone_RES_27 | 0.32 | 0.01 |
| IL-1B | Thompson_FAIL_13 | -0.20 | 0.13 |
| IL-1B | Thompson_RES_5 | 0.32 | 0.01 |
| IL-1B | Zimmer_RES_3 | 0.36 | 0.01 |
| IL-1B | SCC DEG set | -0.03 | 0.85 |
| IL-1B | X-ray M6 DEG set | 0.17 | 0.20 |
| IL-1B | SGRQ M6 DEG set | 0.27 | 0.04 |
| IL-2 | IL-4 | 0.09 | 0.52 |
| IL-2 | IL-6 | 0.42 | 0.00 |
| IL-2 | IL-8 | 0.22 | 0.12 |
| IL-2 | TNF | 0.69 | 0.00 |
| IL-2 | TBS score | -0.01 | 0.97 |
| IL-2 | SGRQ score | 0.02 | 0.90 |
| IL-2 | X-ray score | -0.25 | 0.14 |
| IL-2 | Hb | -0.02 | 0.89 |
| IL-2 | ESR | -0.11 | 0.44 |
| IL-2 | NLR | -0.04 | 0.81 |
| IL-2 | MLR | -0.33 | 0.01 |
| IL-2 | SII | -0.01 | 0.97 |
| IL-2 | Heycken_FAIL_22 | 0.06 | 0.71 |
| IL-2 | Long_RES_10 | 0.09 | 0.53 |
| IL-2 | Tabone_RES_25 | 0.10 | 0.48 |
| IL-2 | Tabone_RES_27 | 0.09 | 0.53 |
| IL-2 | Thompson_FAIL_13 | 0.09 | 0.52 |
| IL-2 | Thompson_RES_5 | 0.23 | 0.10 |
| IL-2 | Zimmer_RES_3 | 0.11 | 0.44 |

| Variable Name (diagonal) | Variable Name (vertical) | Spearman's rank correlation | Adjusted p-value |
|---|---|---|---|
| IL-2 | SCC DEG set | 0.03 | 0.84 |
| IL-2 | X-ray M6 DEG set | 0.08 | 0.58 |
| IL-2 | SGRQ M6 DEG set | 0.09 | 0.55 |
| IL-4 | IL-6 | 0.37 | 0.00 |
| IL-4 | IL-8 | 0.34 | 0.01 |
| IL-4 | TNF | 0.23 | 0.09 |
| IL-4 | TBS score | 0.06 | 0.69 |
| IL-4 | SGRQ score | -0.01 | 0.95 |
| IL-4 | X-ray score | 0.11 | 0.53 |
| IL-4 | Hb | -0.29 | 0.03 |
| IL-4 | ESR | 0.13 | 0.36 |
| IL-4 | NLR | 0.18 | 0.19 |
| IL-4 | MLR | -0.09 | 0.53 |
| IL-4 | SII | 0.16 | 0.25 |
| IL-4 | Heycken_FAIL_22 | 0.28 | 0.03 |
| IL-4 | Long_RES_10 | 0.30 | 0.02 |
| IL-4 | Tabone_RES_25 | 0.31 | 0.02 |
| IL-4 | Tabone_RES_27 | 0.28 | 0.03 |
| IL-4 | Thompson_FAIL_13 | -0.31 | 0.02 |
| IL-4 | Thompson_RES_5 | 0.22 | 0.10 |
| IL-4 | Zimmer_RES_3 | 0.33 | 0.01 |
| IL-4 | SCC DEG set | -0.15 | 0.29 |
| IL-4 | X-ray M6 DEG set | 0.07 | 0.60 |
| IL-4 | SGRQ M6 DEG set | 0.28 | 0.03 |
| IL-6 | IL-8 | 0.33 | 0.01 |
| IL-6 | TNF | 0.39 | 0.00 |
| IL-6 | TBS score | 0.05 | 0.76 |
| IL-6 | SGRQ score | 0.13 | 0.36 |
| IL-6 | X-ray score | 0.04 | 0.85 |
| IL-6 | Hb | -0.35 | 0.01 |
| IL-6 | ESR | 0.23 | 0.10 |
| IL-6 | NLR | 0.11 | 0.42 |
| IL-6 | MLR | -0.06 | 0.67 |
| IL-6 | SII | 0.22 | 0.10 |
| IL-6 | Heycken_FAIL_22 | 0.19 | 0.16 |
| IL-6 | Long_RES_10 | 0.26 | 0.05 |
| IL-6 | Tabone_RES_25 | 0.24 | 0.07 |
| IL-6 | Tabone_RES_27 | 0.23 | 0.10 |
| IL-6 | Thompson_FAIL_13 | -0.18 | 0.18 |
| IL-6 | Thompson_RES_5 | 0.19 | 0.17 |
| IL-6 | Zimmer_RES_3 | 0.33 | 0.01 |
| IL-6 | SCC DEG set | -0.23 | 0.09 |
| IL-6 | X-ray M6 DEG set | 0.11 | 0.44 |
| IL-6 | SGRQ M6 DEG set | 0.26 | 0.05 |
| IL-8 | TNF | 0.41 | 0.00 |
| IL-8 | TBS score | 0.45 | 0.00 |
| IL-8 | SGRQ score | 0.45 | 0.00 |
| IL-8 | X-ray score | 0.44 | 0.00 |

| Variable Name (diagonal) | Variable Name (vertical) | Spearman's rank correlation | Adjusted p-value |
|---|---|---|---|
| IL-8 | Hb | -0.21 | 0.12 |
| IL-8 | ESR | 0.36 | 0.01 |
| IL-8 | NLR | 0.55 | 0.00 |
| IL-8 | MLR | 0.26 | 0.05 |
| IL-8 | SII | 0.56 | 0.00 |
| IL-8 | Heycken_FAIL_22 | 0.42 | 0.00 |
| IL-8 | Long_RES_10 | 0.48 | 0.00 |
| IL-8 | Tabone_RES_25 | 0.44 | 0.00 |
| IL-8 | Tabone_RES_27 | 0.49 | 0.00 |
| IL-8 | Thompson_FAIL_13 | -0.34 | 0.01 |
| IL-8 | Thompson_RES_5 | 0.51 | 0.00 |
| IL-8 | Zimmer_RES_3 | 0.49 | 0.00 |
| IL-8 | SCC DEG set | -0.18 | 0.18 |
| IL-8 | X-ray M6 DEG set | 0.24 | 0.08 |
| IL-8 | SGRQ M6 DEG set | 0.43 | 0.00 |
| TNF | TBS score | 0.14 | 0.32 |
| TNF | SGRQ score | 0.21 | 0.13 |
| TNF | X-ray score | -0.10 | 0.58 |
| TNF | Hb | -0.17 | 0.23 |
| TNF | ESR | 0.05 | 0.77 |
| TNF | NLR | 0.10 | 0.46 |
| TNF | MLR | -0.11 | 0.41 |
| TNF | SII | 0.12 | 0.41 |
| TNF | Heycken_FAIL_22 | 0.22 | 0.11 |
| TNF | Long_RES_10 | 0.24 | 0.07 |
| TNF | Tabone_RES_25 | 0.25 | 0.07 |
| TNF | Tabone_RES_27 | 0.21 | 0.12 |
| TNF | Thompson_FAIL_13 | 0.01 | 0.97 |
| TNF | Thompson_RES_5 | 0.33 | 0.01 |
| TNF | Zimmer_RES_3 | 0.26 | 0.05 |
| TNF | SCC DEG set | 0.13 | 0.36 |
| TNF | X-ray M6 DEG set | 0.35 | 0.01 |
| TNF | SGRQ M6 DEG set | 0.23 | 0.09 |
| TBS score | SGRQ score | 0.79 | 0.00 |
| TBS score | X-ray score | 0.62 | 0.00 |
| TBS score | Hb | -0.32 | 0.02 |
| TBS score | ESR | 0.40 | 0.00 |
| TBS score | NLR | 0.42 | 0.00 |
| TBS score | MLR | 0.15 | 0.29 |
| TBS score | SII | 0.45 | 0.00 |
| TBS score | Heycken_FAIL_22 | 0.44 | 0.00 |
| TBS score | Long_RES_10 | 0.49 | 0.00 |
| TBS score | Tabone_RES_25 | 0.40 | 0.00 |
| TBS score | Tabone_RES_27 | 0.46 | 0.00 |
| TBS score | Thompson_FAIL_13 | -0.38 | 0.00 |
| TBS score | Thompson_RES_5 | 0.53 | 0.00 |
| TBS score | Zimmer_RES_3 | 0.42 | 0.00 |
| TBS score | SCC DEG set | 0.02 | 0.93 |

| Variable Name (diagonal) | Variable Name (vertical) | Spearman's rank correlation | Adjusted p-value |
|---|---|---|---|
| TBS score | X-ray M6 DEG set | 0.34 | 0.01 |
| TBS score | SGRQ M6 DEG set | 0.43 | 0.00 |
| SGRQ score | X-ray score | 0.65 | 0.00 |
| SGRQ score | Hb | -0.22 | 0.10 |
| SGRQ score | ESR | 0.46 | 0.00 |
| SGRQ score | NLR | 0.39 | 0.00 |
| SGRQ score | MLR | 0.23 | 0.09 |
| SGRQ score | SII | 0.49 | 0.00 |
| SGRQ score | Heycken_FAIL_22 | 0.41 | 0.00 |
| SGRQ score | Long_RES_10 | 0.48 | 0.00 |
| SGRQ score | Tabone_RES_25 | 0.41 | 0.00 |
| SGRQ score | Tabone_RES_27 | 0.48 | 0.00 |
| SGRQ score | Thompson_FAIL_13 | -0.25 | 0.06 |
| SGRQ score | Thompson_RES_5 | 0.54 | 0.00 |
| SGRQ score | Zimmer_RES_3 | 0.45 | 0.00 |
| SGRQ score | SCC DEG set | -0.12 | 0.38 |
| SGRQ score | X-ray M6 DEG set | 0.23 | 0.08 |
| SGRQ score | SGRQ M6 DEG set | 0.40 | 0.00 |
| X-ray score | Hb | -0.15 | 0.28 |
| X-ray score | ESR | 0.47 | 0.00 |
| X-ray score | NLR | 0.36 | 0.01 |
| X-ray score | MLR | 0.05 | 0.72 |
| X-ray score | SII | 0.49 | 0.00 |
| X-ray score | Heycken_FAIL_22 | 0.49 | 0.00 |
| X-ray score | Long_RES_10 | 0.49 | 0.00 |
| X-ray score | Tabone_RES_25 | 0.46 | 0.00 |
| X-ray score | Tabone_RES_27 | 0.52 | 0.00 |
| X-ray score | Thompson_FAIL_13 | -0.40 | 0.01 |
| X-ray score | Thompson_RES_5 | 0.44 | 0.00 |
| X-ray score | Zimmer_RES_3 | 0.48 | 0.00 |
| X-ray score | SCC DEG set | -0.13 | 0.44 |
| X-ray score | X-ray M6 DEG set | 0.29 | 0.08 |
| X-ray score | SGRQ M6 DEG set | 0.44 | 0.01 |
| Hb | ESR | -0.52 | 0.00 |
| Hb | NLR | -0.38 | 0.00 |
| Hb | MLR | -0.25 | 0.00 |
| Hb | SII | -0.32 | 0.00 |
| Hb | Heycken_FAIL_22 | -0.43 | 0.00 |
| Hb | Long_RES_10 | -0.46 | 0.00 |
| Hb | Tabone_RES_25 | -0.45 | 0.00 |
| Hb | Tabone_RES_27 | -0.40 | 0.00 |
| Hb | Thompson_FAIL_13 | 0.53 | 0.00 |
| Hb | Thompson_RES_5 | -0.18 | 0.18 |
| Hb | Zimmer_RES_3 | -0.38 | 0.00 |
| Hb | SCC DEG set | 0.11 | 0.41 |
| Hb | X-ray M6 DEG set | -0.32 | 0.01 |
| Hb | SGRQ M6 DEG set | -0.49 | 0.00 |
| ESR | NLR | 0.36 | 0.00 |

| Variable Name (diagonal) | Variable Name (vertical) | Spearman's rank correlation | Adjusted p-value |
|---|---|---|---|
| ESR | MLR | 0.13 | 0.16 |
| ESR | SII | 0.45 | 0.00 |
| ESR | Heycken_FAIL_22 | 0.53 | 0.00 |
| ESR | Long_RES_10 | 0.61 | 0.00 |
| ESR | Tabone_RES_25 | 0.57 | 0.00 |
| ESR | Tabone_RES_27 | 0.60 | 0.00 |
| ESR | Thompson_FAIL_13 | -0.36 | 0.01 |
| ESR | Thompson_RES_5 | 0.37 | 0.00 |
| ESR | Zimmer_RES_3 | 0.55 | 0.00 |
| ESR | SCC DEG set | -0.17 | 0.23 |
| ESR | X-ray M6 DEG set | 0.22 | 0.10 |
| ESR | SGRQ M6 DEG set | 0.53 | 0.00 |
| NLR | MLR | 0.44 | 0.00 |
| NLR | SII | 0.78 | 0.00 |
| NLR | Heycken_FAIL_22 | 0.61 | 0.00 |
| NLR | Long_RES_10 | 0.63 | 0.00 |
| NLR | Tabone_RES_25 | 0.59 | 0.00 |
| NLR | Tabone_RES_27 | 0.62 | 0.00 |
| NLR | Thompson_FAIL_13 | -0.54 | 0.00 |
| NLR | Thompson_RES_5 | 0.53 | 0.00 |
| NLR | Zimmer_RES_3 | 0.60 | 0.00 |
| NLR | SCC DEG set | -0.20 | 0.14 |
| NLR | X-ray M6 DEG set | 0.37 | 0.00 |
| NLR | SGRQ M6 DEG set | 0.61 | 0.00 |
| MLR | SII | 0.38 | 0.00 |
| MLR | Heycken_FAIL_22 | 0.20 | 0.14 |
| MLR | Long_RES_10 | 0.21 | 0.12 |
| MLR | Tabone_RES_25 | 0.16 | 0.23 |
| MLR | Tabone_RES_27 | 0.21 | 0.13 |
| MLR | Thompson_FAIL_13 | -0.13 | 0.34 |
| MLR | Thompson_RES_5 | 0.20 | 0.14 |
| MLR | Zimmer_RES_3 | 0.23 | 0.09 |
| MLR | SCC DEG set | 0.07 | 0.60 |
| MLR | X-ray M6 DEG set | 0.18 | 0.19 |
| MLR | SGRQ M6 DEG set | 0.17 | 0.20 |
| SII | Heycken_FAIL_22 | 0.68 | 0.00 |
| SII | Long_RES_10 | 0.71 | 0.00 |
| SII | Tabone_RES_25 | 0.66 | 0.00 |
| SII | Tabone_RES_27 | 0.71 | 0.00 |
| SII | Thompson_FAIL_13 | -0.57 | 0.00 |
| SII | Thompson_RES_5 | 0.59 | 0.00 |
| SII | Zimmer_RES_3 | 0.67 | 0.00 |
| SII | SCC DEG set | -0.16 | 0.24 |
| SII | X-ray M6 DEG set | 0.39 | 0.00 |
| SII | SGRQ M6 DEG set | 0.68 | 0.00 |
| Heycken_FAIL_22 | Long_RES_10 | 0.96 | 0.00 |
| Heycken_FAIL_22 | Tabone_RES_25 | 0.97 | 0.00 |
| Heycken_FAIL_22 | Tabone_RES_27 | 0.96 | 0.00 |

| Variable Name (diagonal) | Variable Name (vertical) | Spearman's rank correlation | Adjusted p-value |
|---|---|---|---|
| Heycken_FAIL_22 | Thompson_FAIL_13 | -0.55 | 0.00 |
| Heycken_FAIL_22 | Thompson_RES_5 | 0.69 | 0.00 |
| Heycken_FAIL_22 | Zimmer_RES_3 | 0.89 | 0.00 |
| Heycken_FAIL_22 | SCC DEG set | -0.24 | 0.07 |
| Heycken_FAIL_22 | X-ray M6 DEG set | 0.71 | 0.00 |
| Heycken_FAIL_22 | SGRQ M6 DEG set | 0.96 | 0.00 |
| Long_RES_10 | Tabone_RES_25 | 0.97 | 0.00 |
| Long_RES_10 | Tabone_RES_27 | 0.98 | 0.00 |
| Long_RES_10 | Thompson_FAIL_13 | -0.55 | 0.00 |
| Long_RES_10 | Thompson_RES_5 | 0.74 | 0.00 |
| Long_RES_10 | Zimmer_RES_3 | 0.91 | 0.00 |
| Long_RES_10 | SCC DEG set | -0.29 | 0.02 |
| Long_RES_10 | X-ray M6 DEG set | 0.62 | 0.00 |
| Long_RES_10 | SGRQ M6 DEG set | 0.96 | 0.00 |
| Tabone_RES_25 | Tabone_RES_27 | 0.97 | 0.00 |
| Tabone_RES_25 | Thompson_FAIL_13 | -0.57 | 0.00 |
| Tabone_RES_25 | Thompson_RES_5 | 0.69 | 0.00 |
| Tabone_RES_25 | Zimmer_RES_3 | 0.90 | 0.00 |
| Tabone_RES_25 | SCC DEG set | -0.33 | 0.01 |
| Tabone_RES_25 | X-ray M6 DEG set | 0.65 | 0.00 |
| Tabone_RES_25 | SGRQ M6 DEG set | 0.97 | 0.00 |
| Tabone_RES_27 | Thompson_FAIL_13 | -0.53 | 0.00 |
| Tabone_RES_27 | Thompson_RES_5 | 0.74 | 0.00 |
| Tabone_RES_27 | Zimmer_RES_3 | 0.91 | 0.00 |
| Tabone_RES_27 | SCC DEG set | -0.32 | 0.01 |
| Tabone_RES_27 | X-ray M6 DEG set | 0.60 | 0.00 |
| Tabone_RES_27 | SGRQ M6 DEG set | 0.93 | 0.00 |
| Thompson_FAIL_13 | Thompson_RES_5 | -0.24 | 0.07 |
| Thompson_FAIL_13 | Zimmer_RES_3 | -0.44 | 0.00 |
| Thompson_FAIL_13 | SCC DEG set | 0.40 | 0.00 |
| Thompson_FAIL_13 | X-ray M6 DEG set | -0.29 | 0.03 |
| Thompson_FAIL_13 | SGRQ M6 DEG set | -0.61 | 0.00 |
| Thompson_RES_5 | Zimmer_RES_3 | 0.74 | 0.00 |
| Thompson_RES_5 | SCC DEG set | -0.27 | 0.04 |
| Thompson_RES_5 | X-ray M6 DEG set | 0.36 | 0.00 |
| Thompson_RES_5 | SGRQ M6 DEG set | 0.70 | 0.00 |
| Zimmer_RES_3 | SCC DEG set | -0.27 | 0.04 |
| Zimmer_RES_3 | X-ray M6 DEG set | 0.60 | 0.00 |
| Zimmer_RES_3 | SGRQ M6 DEG set | 0.86 | 0.00 |
| SCC DEG set | X-ray M6 DEG set | 0.28 | 0.03 |
| SCC DEG set | SGRQ M6 DEG set | -0.35 | 0.01 |
| X-ray M6 DEG set | SGRQ M6 DEG set | 0.66 | 0.00 |
